# Is intimate partner violence decreasing? Analysis of population-level trends in 21 countries

**DOI:** 10.64898/2026.01.13.26344010

**Authors:** Milly Marston, Amber Peterman, Tia Palmero, Carolina Coll, Karen Devries

## Abstract

**Introduction:** Around the world, 193 countries have pledged to eliminate violence against women and girls by 2030, the most common form of which is intimate partner violence (IPV). To verify if progress is being made, we examined trends in prevalence of IPV against women and girls across 21 countries with three or more Demographic and Health Surveys. Additionally, we examine whether trends differ by type, frequency and severity of IPV; and whether changes are concentrated in specific sub-groups of women and girls.

**Methods:** We pooled individual-level data from within-country surveys conducted over time and harmonised outcomes to ensure comparability. We estimated weighted crude prevalence ratios and used log binomial regression models to calculate age adjusted risk ratios and predict annual age adjusted prevalence of 12-month physical and/or sexual IPV and severe IPV against women and girls aged 15-49, overall and by age group, education, wealth and place of residence. We used an ordered logit model to estimate the frequency of 12-month physical and/or sexual IPV.

**Results:** We found no consistent cross-country trends over time in IPV prevalence: five countries showed declines, 15 demonstrated mixed directions or no changes, and one a consistent increase. In most countries, levels of frequent or severe IPV changed in the same way as overall prevalence, but four countries with an overall decline showed an increase in the proportion of severe IPV. Changes were not systematically concentrated in younger, wealthier, or more educated women and girls.

**Conclusion:** Progress towards reducing IPV against women and girls cannot be assumed. Overall, less than a quarter of examined countries showed consistent declines over approximately a decade prior to the most recent data collection. Further efforts leveraging evidence-based and locally adapted, scalable interventions and policies are urgently needed to accelerate prevention and to respond to IPV against women and girls.

**Key messages:** *What is already known on this topic?:* Around the world, 193 countries have pledged to eliminate violence against women and girls by 2030. A recent paper has concluded that prevalence of IPV is declining in low- and middle-income countries, with an average annual rate of change of –0.2% (95% CI –0.4 to – 0.03) (https://www.thelancet.com/journals/langlo/article/PIIS2214-109X(23)00417-5/fulltext). However, this recent paper pooled multiple survey datasets from 21 countries, across only two time points, despite more surveys being available, which limits the ability to identify overall trends. In addition, the study does not ensure consistent items are included across surveys. This leaves open the possibility that the declines observed are due to methodological differences between surveys and/or are not consistent within countries.

*What this study adds:* We address many of the methodological concerns raised from the previous analysis, including the standardisation of outcomes, and the inclusion of countries which have at least three measurement points—thus more confidently identifying trends, typically over a decade or longer. This leads us to a different overall conclusion. We find that the global prevalence of physical and/or sexual IPV against women and girls is not systematically declining across countries. Overall, less than a quarter of examined countries showed consistent declines in physical and/or sexual IPV over time, namely in Colombia, Kenya, Philippines, Tanzania, Uganda. Further, we find that in Zambia, Kenya, Nigeria and Zimbabwe, as overall physical IPV prevalence reduces, the proportion of women and girls who experience severe physical IPV increases.

*How this study might affect research, practice or policy:* Our results suggest that further efforts leveraging evidence-based and scalable interventions and policies are urgently needed to accelerate prevention and response to IPV against women and girls. Second, careful monitoring is also needed to ensure gains are sustained equally across vulnerable groups. Finally, sustaining and strengthening national data collection architecture is critical to enable continued progress towards elimination of IPV against women and girls.

## Introduction

Intimate partner violence (IPV) against women and girls is a major global public health and human rights issue. Experience of IPV is associated with a wide range of adverse health outcomes, including incident major depressive disorder, anxiety disorders and self-harm, induced abortion, and HIV/AIDS(1). The first global prevalence modelling study estimated that in 2010, 30% of women aged 15 years and over had experienced physical and/or sexual violence from an intimate partner in their lifetime(2). In recognition of the huge societal costs of IPV, reducing the prevalence of IPV is a key indicator of achievement of Sustainable Development Goal (SDG) Target 5.2, the ‘elimination of all forms of violence against women and girls(3).

With five years remaining to achieve the 2030 SDGs, are we on track to hit our target for IPV? So far, we have no clear answer. The latest global estimates calculated in 2018 put lifetime prevalence at 27%(4), which is not statistically different from the global estimates from 2010(2, 4). However, the 2010 and 2018 global estimates are not designed to be directly comparable over time, as they pool data from recent estimates across countries (2, 4). Thus, estimates suffer from compositional changes in countries included (81 countries in 2010 to 161 countries in 2018), making these comparisons misleading. Further, these global estimates, cannot inform country-specific policy and programming efforts(2, 4). Other existing trend analyses compare different measures or geographies using two time points, making it unclear whether trends are true changes in prevalence, or might be masking fluctuations over time or changes due to methodological differences between surveys(5, 6).

In addition to overall trends in population prevalence at the national level, there are many other critical unanswered questions for policy and program implementation. For example, are potential reductions in overall prevalence driven by decreases in less severe acts of violence, leaving women who are experiencing severe violence or more frequent acts of violence behind? Further, do we know whether changes in IPV prevalence are occurring equitably across different population groups of women and girls? The answers to these questions are important to inform population-level and targeted prevention strategies to address IPV.

We update and extend previous efforts to answer the central question: is women’s and girls’ experience of past-year physical and/or sexual IPV decreasing, and are changes concentrated in specific subgroups? Similar to previous analyses, we use data from individual recode of the nationally representative Demographic and Health Surveys (DHS), which ask standardized IPV questions to women and girls of reproductive age (from 15 to 49 years). However, we address limitations in previous analyses by using repeated cross-sectional data from countries with three or more DHS. Taken together, surveys typically span over a decade of time, with sufficient data points to assess if initial trends between shorter time periods hold over the longer term. In addition, we ensure comparability and harmonization of questions on women’s and girls’ experience of physical and/or sexual IPV across survey rounds. The overall objective of the paper is to provide evidence towards collective progress towards Target 5.2, as well as if progress differs by type, frequency and severity of IPV, age group, and risk factors, including educational attainment, urban or rural residence, and household wealth.

## Methods

### Data

The DHS are multi-topic nationally representative cross-sectional household surveys led by national governments with technical assistance from IFC International(7). Since its inception in 1984, the DHS program has collected representative data on a wide range of demographic and health topics through more than 400 surveys in over 90 countries. The DHS serve as a source of data for monitoring 30 SDG indicators, including for child and maternal mortality, nutrition, family planning, food security, access to clean water, and IPV, among others(8).

The DHS started collecting data on IPV in 1990, drawing on experience from the WHO-led multi-country study on Women’s Health and Domestic Violence Against Women(9). The DHS collect behaviourally specific indicators for violent acts of violence perpetrated by an intimate partner, sampling typically one women or girl of reproductive age (15 to 49) per household. All interviews for the domestic violence module are conducted in adherence to ethical protocol and safety of participants in mind. For example, interviews are carried out by female interviewers in private spaces and who undergo specialised training on how to ask questions on sensitive topics. Further, both survey participants and interviewers are supported with referrals and debriefing to address potential adverse events(10).

In building the sample for this analysis, we consider the evolution of the DHS over time, specifically changes in Phase 4 (starting in 1997) and Phase 5 (starting in 2003)(11). Across phases, questions or question wording, as well as the coding of some variables in the domestic violence module changed. We carefully ensured consistency in items and samples to rule out changes due to methodological differences between surveys within a country. We used the original questionnaires from the DHS reports, the datasets, and finally the DHS indicator manual(7), to check: the number of questions asked, the consistency of the question wording, references to partners included in questions (i.e., current or past partners) and who questions were asked to (i.e., who was considered an ‘ever-partnered woman’). All countries with three or more surveys administering the domestic violence module which allowed consistent indicators over time were eligible for inclusion in the current study. We required at least three survey waves to avoid making conclusions about trends with just two time points, which may be misleading due to relatively short periods to assess trends(7). Table S1 gives more information on alignment of samples and indicators over time for each included country.

### Definition of ‘ever-partnered women’

The denominator for our main analyses included women and girls aged 15 to 49 who had ever been in a union. In most countries this was defined by the surveys as ever married or cohabited with a partner. Some surveys excluded widows from their samples entirely (Cameroon 2004, Malawi 2004, Mali 2006) or did not ask widows all questions (Cameroon 2011, Nigeria 2008, Uganda 2006 and 2011, Haiti 2005-06, India 2005-06). In addition, some countries asked domestic violence questions of slightly different age ranges. These deviations from the standard sample are described in table S3 by country and survey wave.

### Definition of intimate partner

All surveys asked women about experiences over the past 12 months from their ‘current or most recent husband or partner’, which correspond to the main indicators used in our analysis. From recode Phase 6 (starting in 2008) onwards, the majority of surveys also asked about violence over the same time frame from any partner, however they did not include individual violence items. Instead, questions encompassing any partner asked more generally whether any partner “ever hit, slap, kick or physically hurt respondent” and whether any partner “physically forced to have sex or to perform sexual acts.” This means that these indicators could not be standardised or combined with the majority of measures corresponding to the current or most recent partner. To see if excluding violence from previous partners in the recent 12-month period would change our estimates, we repeated the analysis including any recent violence from previous partners in a sensitivity analysis.

### Definitions of recent violence

Recent physical and/or sexual IPV was defined following the SDG target 5.2.1 as answering yes to any of the questions on experiencing acts of physical and sexual violence in the last 12 months. For physical violence these acts included being: a) pushed, shook or had something thrown by husband/partner, b) slapped by husband/partner, c) punched with fist or hit by something harmful by husband/partner, d) kicked or dragged by husband/partner, e) strangled or burnt by husband/partner, f) threatened with knife/gun or other weapon by husband/partner, g) arm twisted or hair pulled by husband/partner. For sexual violence these acts included being: a) physically forced into unwanted sex by husband/partner, b) forced into other unwanted sexual acts by husband/partner, c) physically forced to perform sexual acts respondent did not want to by husband/partner (See table S1 for items included for each country). We also created separate indicators for each type of IPV (physical, sexual) for disaggregated analyses.

Individuals with missing data for all items were excluded from the analysis. When an individual reported experiencing an IPV item but not whether it was recent IPV (i.e. whether it occurred in the last 12 months) they were still included, based on their remaining responses to the other items. To assess the impact of including individuals who were classified as not experiencing recent IPV, but had at least one positive response to an item, but did not report whether it was recent or not, we conducted a sensitivity analysis by reclassifying these individuals as having experienced recent IPV to determine whether it changed the overall results.

In addition to the main prevalence indicator, we also assessed trends in IPV frequency and severity. For frequency, following previous analyses, a score was created by summing the number of items of physical or sexual violence reported, with a score of 1 assigned to each item if an act occurred “sometimes” and 2 if it occurred “often”(12, 13). For this analysis, surveys using continuous variables to measure frequency (from Phase 4) were excluded to maintain comparability with later surveys (Phase 5 and later). This resulted in dropping of four countries: Cameroon, Dominican Republic, Malawi and Mali. Finally, we looked at changes in the prevalence of severe physical violence, modelled as a binary variable. Those who reported any of the physical violence acts (d) - (f) mentioned above were classified as experiencing severe violence

### Other variables

We modelled age in two ways: First in five-year age groups (15-19 years, and so on), and second as younger (aged 15-24 years) and older groups (aged 25-49 years). Wealth was modelled as a categorical variable, using household wealth quintiles based on country specific indices pre-computed in the DHS(14) categorised into three groups: lowest two quintiles (poorest/poor); middle quintile (middle); and highest two quintiles (richer/richest). Education was grouped into none, some primary and some secondary and higher, and place of residence was urban or rural.

### Data analysis

All analyses were conducted in Stata 18.0, accounting for the two□stage cluster sampling survey design, and using the domestic violence weights provided by DHS, which adjust for the probability of selection into the domestic violence module. Individual level data from each survey were pooled to calculate crude prevalence ratios. To examine whether the risk of experiencing recent physical and/or sexual IPV changed between surveys, we estimated a log binomial regression model for each country, adjusting for survey as a categorical variable and five-year age groups to account for demographic trends. The sub-sample for this analysis was ever-partnered women.

To assess changes in the frequency of physical and/or sexual IPV, we fit ordinal logistic regression models. The IPV score (representing ordered categories) was the dependent variable, adjusted for survey as a categorical variable and five-year age groups. The analysis sample to assess changes in frequency was women who had experienced recent physical and/or sexual IPV. To estimate changes in prevalence of severe IPV, we estimated a log binomial regression model for each country, adjusting for survey as a categorical variable and five-year age groups, to account for demographic trends. The denominator for this analysis was women who reported any physical IPV in the past 12 months.

Since the intersurvey intervals vary by country and are at different calendar times we also looked at predicted prevalence ratios by calendar year, were derived from log binomial regression models using the binary IPV outcome as the dependent variable and adjusted for five-year age groups. To account for time-varying changes, separate models were specified for each consecutive survey pair within a country. The midpoint year of each survey was treated as a continuous variable, allowing predictions of prevalence for each year between surveys. This assumed a constant rate of change during the intersurvey interval in the absence of additional data points. To compare the rate and direction of change in IPV over calendar time across countries we also estimated the marginal effects using the models described above.

To investigate whether trends in IPV varied by place of residence, education and wealth, we used log binomial regression models with an added interaction term between survey and variable of interest in separate models. All estimates were adjusted for five-year age groups. With these models we produced estimated stratum specific risk ratios and used a Wald test to assess whether there was evidence of a differential trend.

We conducted a range of additional sensitivity analyses to assess whether our results were robust to small variations in a variety of methodological choices mentioned above. These include: item wording of violent acts, limitation of violence questions to current (versus current and previous) partner, definitions of ‘ever-partnered’, missing data and robustness to potential data quality issues specific to Rwanda (Supplementary Materials page 9). Details are reported in Supplementary Materials pages 52-56.

## Results

There were 55 countries with 123 surveys conducted between 2000 and 2024 with available data from the domestic violence module (assessed December, 2024, Supplementary materials table S2). Of these, 21 countries had at least three surveys, comprising 74 surveys total. After assessing the comparability of questions across surveys, we included a final sample of 21 countries with 68 surveys taking place from 2002 to 2023. We organize results by WHO region, while noting that direct comparisons across countries is not warranted due to differences in survey years, indicators and samples. The analysis countries include twelve in the African Region (Cameroon, Kenya, Malawi, Mali, Mozambique, Nigeria, Rwanda, Senegal, Tanzania, Uganda, Zambia, Zimbabwe), four in the Americas (Colombia, Dominican Republic, Haiti, Peru), three in the South-East Asian Region (Cambodia, India, Nepal) and one in each of the Eastern Mediterranean Region (Jordan) and the Western Pacific Region (Philippines).

Characteristics of analytic samples, including the year of survey, age and partnership status of surveyed women, number of unions and sample sizes are described by country in tables S3 and S4. We present further descriptive statistics for our sub-group indicators by country in table S5, including five-year age groups, place of residence, education level and wealth. Figure 1 plots unadjusted prevalence by country and survey, disaggregated by physical only, physical and sexual, and sexual only, with 95% confidence intervals (CI) (see table S7 for further details). Unadjusted prevalence of recent physical and/or sexual IPV varied between a low of 4.1% (95% CI 3.6-4.7%) in the Philippines (2022) and a high of 44.8% (41.5-48.1%) in Uganda (2006). Other comparatively high prevalence countries for at least one round include Zambia, Tanzania, and Cameroon, while low prevalence countries include Cambodia, Senegal and Jordan.

**Figure 1:**
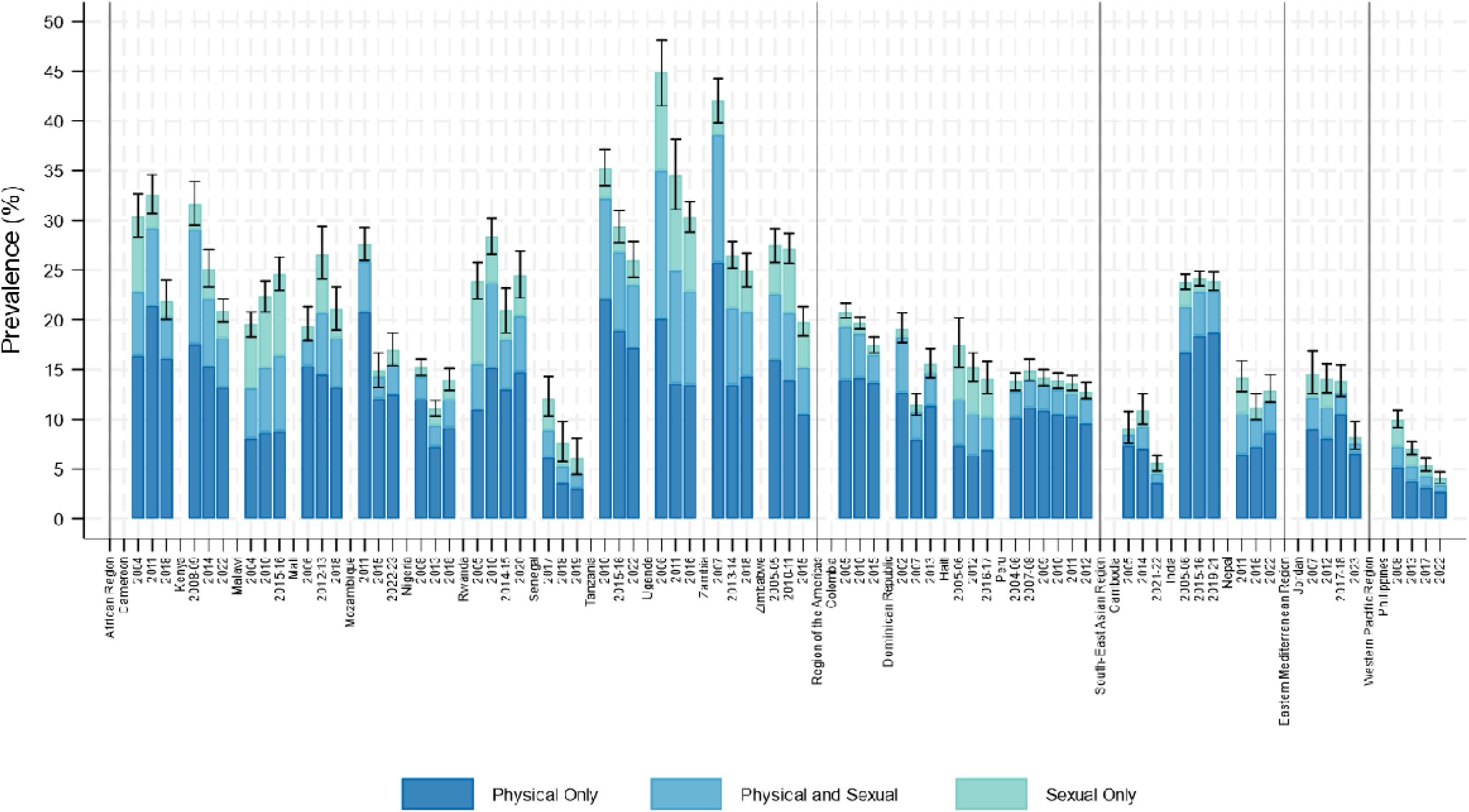
Prevalence of recent physical and/or sexual IPV by survey and country. Countries are ordered by WHO region and alphabetically within region. The prevalence bars and 95% confidence intervals are for any recent (12-month) physical and/or sexual IPV. Estimated prevalence for sexual IPV ranged from 1.7% (1.1-2.4%) in Jordan 2023 to 24.8% (22.3-27.4%) in Uganda 2006. Prevalence of physical IPV ranged from 4.4% (4.0-5.4%) in Cambodia 2021-22 to 34.9% (31.6-38.4%) in Uganda 2006.

### Is the prevalence of physical and/or sexual IPV decreasing?

Figure 2 shows graphical depiction of trends from country-specific log binomial regression models adjusting for age groups, where colours represent significance of trends (green for reductions, grey for no change and red for increases). We found no clear evidence of an overall trends across countries examined in prevalence of recent physical and/or sexual IPV. Instead, trajectories over time varied within countries. Overall, five countries showed evidence of a consistent and statistically significant downward trend in prevalence of physical and/or sexual IPV over survey periods (Colombia, Kenya, Philippines, Tanzania, Uganda). An additional seven countries showed a statistically significant decline in at least one period with no evidence of an increase in any other period (Cambodia, Cameroon, Jordan, Mozambique, Senegal, Zambia and Zimbabwe). There was no evidence suggesting statistically meaningful change in prevalence for three countries over any period (Haiti, India and Peru). Finally, there was a mixed pattern with evidence of statistically significant increases and decreases over different periods in five countries (Dominican Republic, Mali, Nepal, Nigeria and Rwanda). Finally, in one country, Malawi, there was evidence of a consistent and statistically significant increase in any recent physical and/or sexual IPV over the survey period. Table S8 presents marginal effects associated with these results to quantify the exact rate and direction of change, as well as level of statistical significance.

**Figure 2:**
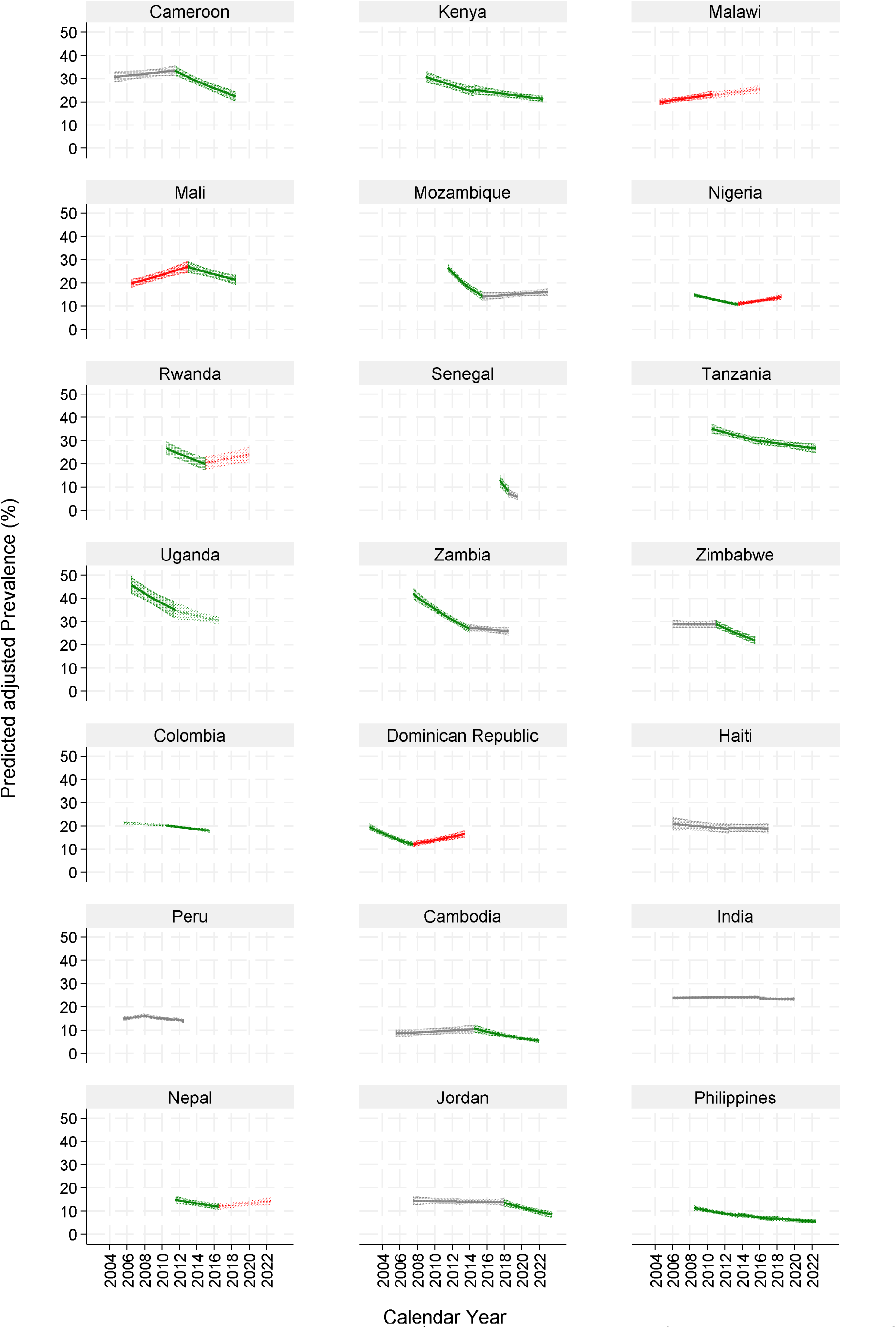
Predicted prevalence of any recent sexual and/or physical IPV, by country. Countries are ordered by WHO region and alphabetically within region. The prevalence and 95% confidence intervals are for any recent (12-month) physical and/or sexual IPV, from log binomial regression models adjusted for five-year age groups. Green is where there is strong evidence of a decline in the period (p<0.01); lighter green is weak evidence of a decline (p<0.05); red is strong evidence of an increase in the period (p<0.01); lighter red is weak evidence of an increase (p<0.05); grey there is no evidence of a change in prevalence over the period.

Figures S2 and S3 replicate results for changes in prevalence of recent physical IPV and sexual IPV separately. Comparing overall trends to those disaggregated by type, we found no clear overall patterns suggesting that changes in overall IPV prevalence are driven primarily by changes in physical IPV or sexual IPV. In seven countries (Haiti, Peru, Kenya, the Philippines, Jordan, Senegal, and Uganda) prevalence of disaggregated measures of physical IPV and sexual IPV changed in the same direction as overall prevalence. In the remaining 14 countries, the proportion of IPV that was physical IPV or sexual IPV varied either in direction or significance levels compared to overall trends in IPV. For example, In Nigeria the first period showed a decrease in physical IPV followed by an increase in the second period, however sexual IPV increase over both periods. In India, the overall trends in recent physical and/or sexual IPV showed no evidence of change, however in the first period there was evidence of an increase in recent physical IPV but a decrease in recent sexual IPV.

### Are there changes in frequency or severity of IPV for women experiencing IPV?

Figure 3 compares odds ratio estimates for changes in prevalence of recent physical and/or sexual IPV (in blue) to those for frequency of the same (in orange), where available. Comparison of the two shows how the composition of frequency may be changing within the overall trends in prevalence. In most countries, among women experiencing IPV, the frequency of IPV did not change significantly. However, in Uganda, where there was a significant decrease in overall prevalence of IPV, there was also a significant decrease in frequency of IPV for those experiencing it, suggesting additional *improvements* in quality of life and freedom from everyday violence for women and girls experiencing IPV. Conversely, in Zambia, while there was evidence of a significant decrease in recent IPV, this was paired with a significant increase in frequency of IPV (although trends were only weakly significant). This suggests that in Zambia, even though a smaller proportion of women and girls are experiencing IPV, quality of life and freedom from everyday violence could be *decreasing* among those women and girls.

**Figure 3:**
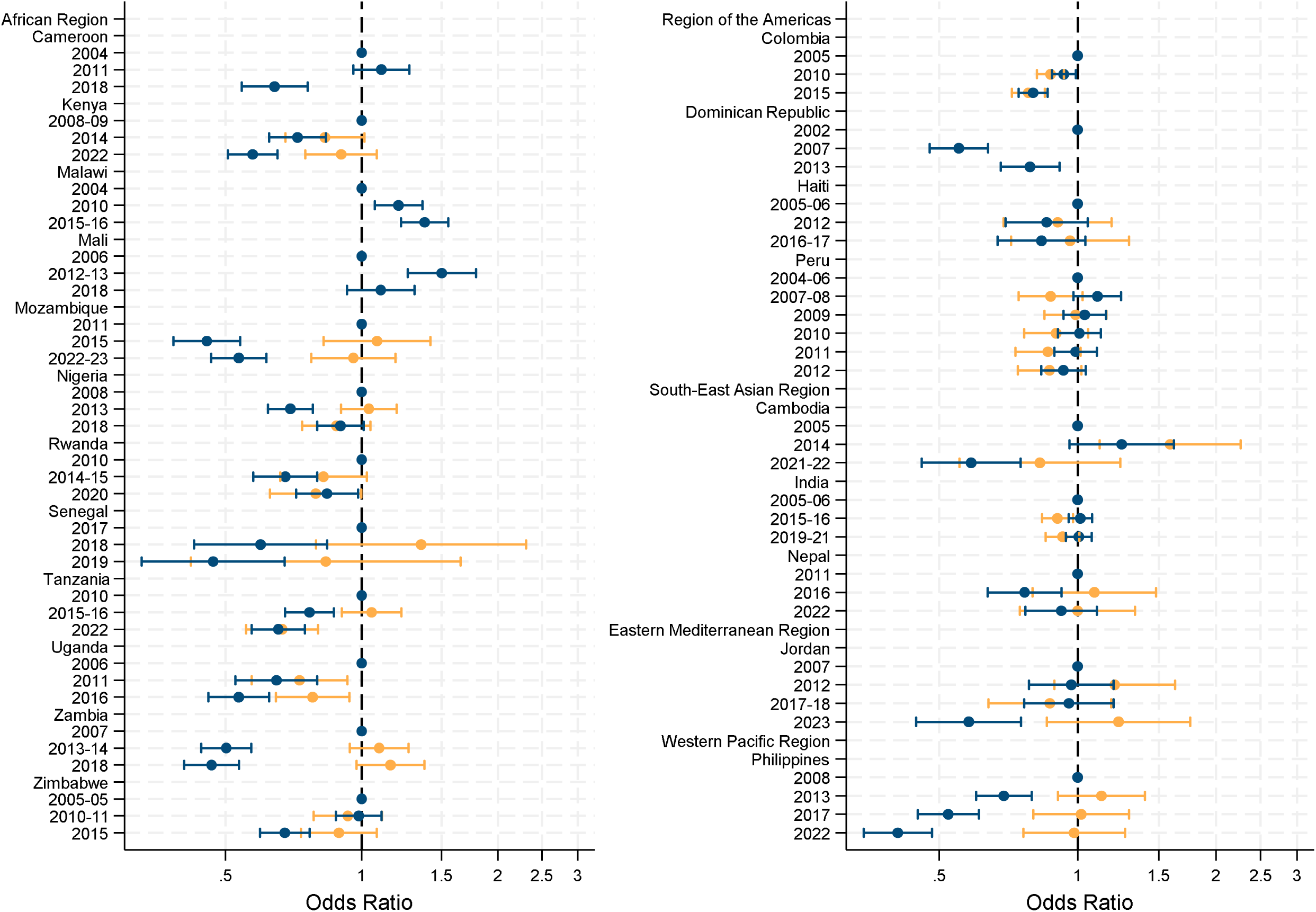
Odds ratios of any recent physical and/or sexual IPV trends (blue) and change in frequency (orange), by country. Odds ratios and 95% confidence intervals are for prevalence of any recent (12-month) physical and/or sexual IPV (blue) and frequency of any recent (12-month) physical and/or sexual IPV (orange), among those who experienced recent IPV, compared to baseline surveys from logistic and ordinal logistic regression models, respectively, adjusted for five-year age groups. Frequency of IPV Cameroon 2004, Dominican Republic 2002, Malawi 2004, Mali 2006 are omitted as frequency was collected differently in the baseline round so therefore may not be comparable. In each figure, *a* blue dot with confidence intervals crossing an odds ratio =1 indicates that there is no evidence of a change in the proportion of recent physical and/or sexual IPV compared to the baseline survey. Blue dots with confidence intervals strictly greater than 1 indicate that, there is an increase in the proportion of those who experience recent IPV. Conversely, a blue dot and confidence intervals strictly less than 1 indicates that there is a decrease proportion of women experiencing recent IPV. Similarly, in each figure, an orange dot with confidence intervals crossing an odds ratio =1 indicates that for those who experience recent physical and/or sexual IPV there is no evidence in a change in the odds of experiencing more frequent recent physical and/or sexual IPV. Therefore, if overall prevalence of recent physical and/or sexual IPV decreases, a non-significant odds ratio means that among those women who experience recent IPV, their experience of more frequent IPV remains the same. Orange dots with confidence intervals strictly greater than 1 indicate that as overall IPV prevalence changes, there is an increase in the proportion of those who experience recent IPV experiencing more frequent IPV. Conversely, an orange dot and confidence intervals strictly less than 1 indicates that as overall IPV prevalence changes, among women who experience recent IPV, there is a decrease proportion of women experiencing more frequent IPV.

A different picture emerges in Figure 4, which shows a similar comparison of risk ratio estimates for changes in prevalence of recent physical IPV (in blue) and severe recent physical IPV (in orange). For most countries, among women and girls who experienced physical IPV, there was no evidence of a change in the proportion of women and girls experiencing severe physical IPV. However, in four countries (Zambia, Kenya, Nigeria and Zimbabwe), there was evidence that alongside a significant reduction in recent physical IPV, there was a corresponding increase in severe physical IPV. Conversely, in Rwanda and Colombia, we observe both a significant reduction in recent physical IPV and the severity of recent physical IPV. These results indicate the importance of going beyond prevalence to examine characteristics of abuse women and girls face over time.

**Figure 4:**
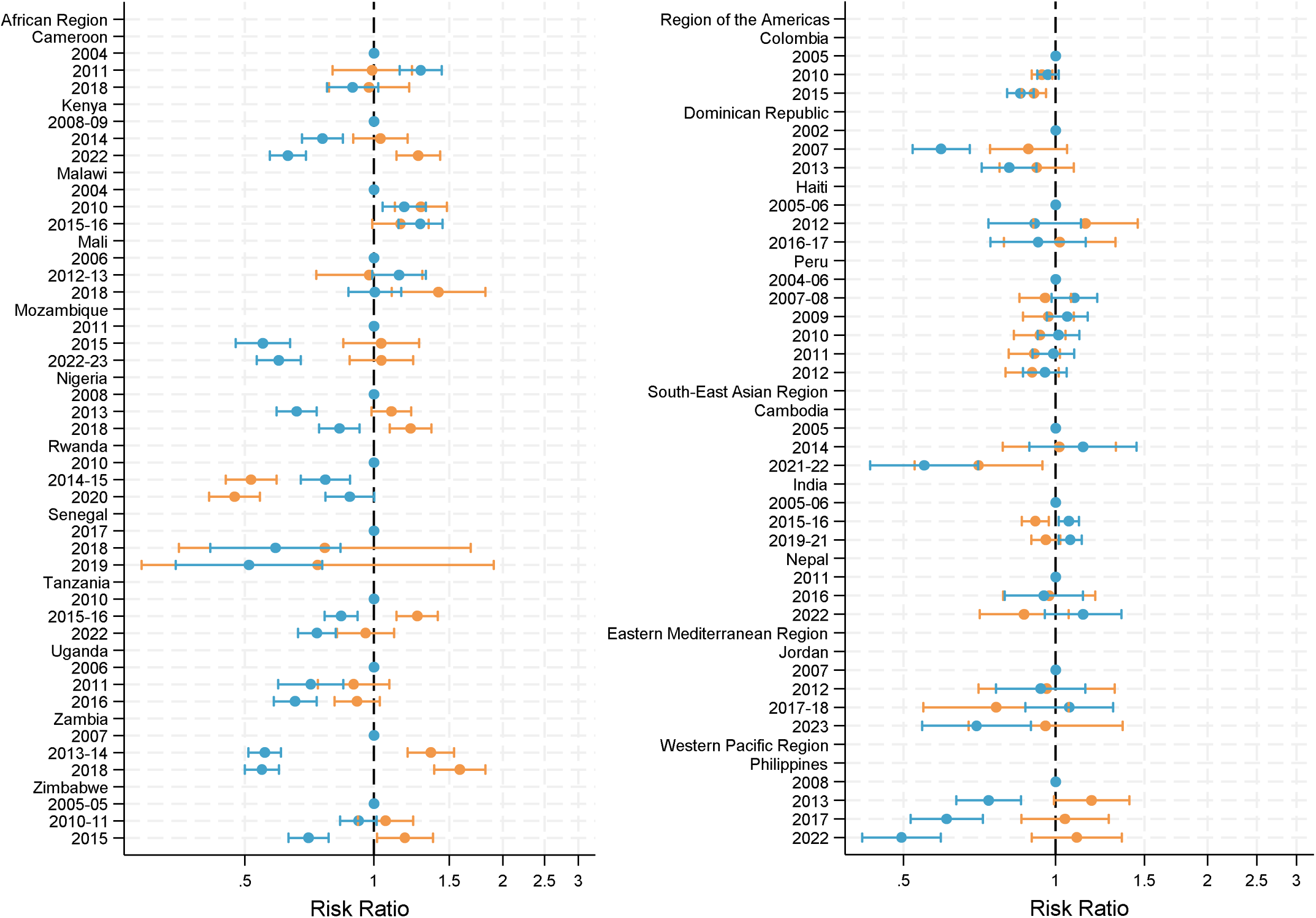
Risk ratios of trends in recent physical IPV (blue) and recent severe physical IPV (orange), by country. Risk ratios and 95% confidence intervals are for the prevalence of any recent (12-month) physical IPV (blue) and any severe recent (12-month) physical IPV (orange), among those who experienced recent physical IPV, compared to baseline surveys from log binomial regression models adjusted for five-year age groups. In each figure, blue dots with confidence intervals crossing an odds ratio =1 indicates that there is no evidence of a change in the proportion of recent physical IPV compared to the baseline survey. Alternatively, blue dots with confidence intervals strictly greater than 1 indicate that, there is an increase in the proportion of those who experience recent physical IPV. Conversely, a blue dot and confidence intervals strictly less than 1 indicates that there is a decrease proportion of women experiencing recent physical IPV. In each figure, an orange dot with confidence intervals crossing a risk ratio=1 indicates that for those who experience recent physical IPV there is no change in the risk of experiencing severe recent physical IPV. Therefore, if overall prevalence of recent physical IPV decreases, a non-significant risk ratio means that among those women who experience recent IPV, their experience of severe IPV remains the same. Orange dots and confidence intervals strictly greater than 1 indicate that as overall physical IPV prevalence changes, there is an increase in the proportion of those who experience severe physical IPV. Conversely, an orange dot and confidence intervals strictly less than 1 indicates that as overall physical IPV prevalence changes, among women who experience recent physical IPV, there is a decrease proportion of women experiencing severe IPV.

### Are changes concentrated in younger, more educated, more urban and richer women?

Overall, we do not find clear and consistent evidence of changes being concentrated in any subgroups of women across countries, indicating that where changes in IPV prevalence are occurring, these are broadly happening in an equitable way. We do, however, find country-specific differences. First, overall, trends in recent physical and/or sexual IPV were similar among younger age groups (15–24-year-olds) and older age groups (25–49-year-olds) (figure. S4 table S9). However, in India there was evidence of a reduction in recent physical and/or sexual IPV among 15–24-year-olds, but not among 25–49-year-olds (Wald test F=3.06 p<0.0001). Second, there was also no clear consistent pattern overall in terms of women’s education levels (figure S5 table S10). However, again in India, the increase in IPV between surveys occurred among women with secondary and higher education, as compared to those with lower education levels. Conversely, in Zambia there is a larger decrease in IPV across surveys among those with secondary and higher education, as compared to those with lower education. Third, women living in urban areas showed similar reductions in IPV as compared to rural women (figure S6 Table S11). Finally, we did find some suggestion of differences in trends by household wealth, however with no clear picture across countries (figure S7, table S12). In Cambodia, Cameroon, Kenya and Zambia IPV declined at higher rates in the richest two quintiles compared to the poorest two quintiles between surveys. However, in Nigeria there was an increasing trend of IPV that was larger in the poorest two quintiles as compared to the richest two quintiles.

### Sensitivity analyses

Our results were robust to sensitivity analyses for dealing with missings for the recall period of IPV (table S13), reference partner (figure S8), or varying item wording for different behavioural acts (figure S9). There are minor exceptions to robustness of results. For example, only a small proportion of the final domestic violence sample we used had missing data for all items between 0.00 and 0.58%. All surveys except four had missings under 1% for recall period of acts, including: Cameroon 2004 (2.19%), Jordan 2007 (1.49%), Zambia 2007 (1.63%) and Zimbabwe 2005-06 (2.79%). If we assume the most extreme case, that all women and girls with missing recall experienced recent IPV, only Zimbabwe trends would change (switching to significant declines over both survey periods). Figure S8 shows almost identical trends across countries while varying the reference to any most recent partner versus current partner (in the last 12 months). Finally, figure S9 shows results from a robustness check excluding two physical IPV items which changed in wording over time in four countries: Cambodia, Cameroon, Dominican Republic and Malawi. While overall prevalence changes due to the removal of items with wording changes, overall trends remain identical.

## Discussion

We analyse 68 surveys representing 21 countries taking place from 2002 to 2023 to assess if IPV is decreasing over time. We do not find overall evidence suggesting IPV prevalence is decreasing, instead, our findings suggest that progress towards SDG target 5.2 is highly uneven across countries. Of the countries we examined, only four countries had evidence of consistent declines over time, while one had a consistent increase. In most countries the frequency and severity of IPV changed in the same way as the overall prevalence of physical and/or sexual IPV. This suggests that where IPV prevalence has declined, the proportion of women and girls who continue to experience severe physical IPV has also declined. However, in four countries (Kenya, Tanzania, Zambia and Zimbabwe), data suggest that declines in IPV prevalence may be driven by reductions in less severe physical acts of violence. The implications of this finding are that among women and girls still experiencing IPV, IPV is more severe.

We found no clear pattern of increase or decreasing trend by age group, educational attainment, urban/rural residence, or household wealth. This suggests that when prevalence changes, it generally does so in an equitable way. However, it also suggests that younger cohorts of women and girls, where we might expect changes to originate, are not showing substantial differences over time as compared to older cohorts. Nonetheless, important exceptions exist within specific countries. Notably in India, younger women saw a decline in prevalence of IPV that was not observed among older women; but more educated women experienced a greater increase IPV compared to less educated women. This is consistent with a growing prevalence of attitudes supporting gender equality among young women in India(15). However, there are also considerably more young women attending higher education in India than in the past(16), alongside recent high-profile cases of young women attending university who have suffered extreme gender-based violence and femicide(17). Our findings clearly point to the importance of context and the importance of understanding country-specific changes. Stakeholders cannot assume common trends across settings and must monitor both progress and equity of progress at a national level.

Further research is needed to understand what factors are driving trends in IPV prevalence at the national level. The only multi-level cross-national analysis of factors associated with country level IPV prevalence(18) finds both gender attitudes and country income level are associated with IPV prevalence, but our findings suggest that reductions in IPV are not automatic and do not follow directly from changes in attitudes and national income levels. Over the period studied, seven of the countries moved up in national income classifications (Cambodia, Cameroon, Colombia, Dominican Republic, India, Peru and Nepal) while only one moved down (Jordan)(19). It is possible that national changes in gender attitudes and income levels are concentrated among sub-groups of the population less affected by IPV, but overall, the relationship between broad development indicators such as gender attitudes and country income is not straightforward. Our results suggest that in many settings, countervailing factors may be maintaining or worsening levels of IPV. Further analysis is needed to understand what these factors may be.

Our results on frequency and severity of IPV are novel. They highlight the importance of understanding changes in prevalence of severe IPV, as a proportion of all IPV. Our results suggest that in some countries these women are being left behind when there are overall population level reductions in IPV. Further research is needed to understand these dynamics in population prevalence, but it could be that interventions in these settings are reaching men inclined to perpetrate less severe IPV or who already held more progressive attitudes. Men who perpetrate more severe IPV may differ from those who perpetrate less severe IPV, by exhibiting more antisocial personality characteristics, and having more conservative and entrenched attitudes towards women(20). Women and girls who experience more severe IPV may also be more isolated, be more likely to suffer from poor mental health and therefore less able to access any existing intervention support.

Our findings differ from other efforts to examine trends in IPV prevalence, as we address several important methodological limitations of previous work. Another analysis of DHS datasets concludes that IPV is decreasing in low- and middle-income countries at an annual negative change rate of 0.2 percent(5). However, this analysis includes countries with only two survey time points, which limits the ability to identify trends. In addition, the analysis pools data from different countries at different time points, potentially identifying decreases as a function of which countries have available surveys in different years. Further, the analysis used DHS-provided derived variables on country level prevalence(5), which include different items and question framings over time(5, 6), making it unclear whether observed changes in prevalence are the result of a real decline or a methodological artifact. We posit that a global statistic of change is misleading, given the wide variation in patterns across countries, as well as the small number of settings with sufficient surveys to confidently analyse trends. Our findings illustrate that situations vary at country level, and thus, our results should not be taken as generalisable to other countries and time periods.

Our findings indicate that the global community is not on track to meet SDG 5.2, reducing physical and/or sexual IPV against women and girls, despite apparent global changes in attitudes more supportive of gender equality. Further, none of the four countries showing consistent downward trends are close to achieving elimination of IPV. This underscores the need for sustained efforts to reduce IPV, including to maintain and/or increase investments in strategies identified as effective in reducing IPV and their scale-up. To support countries, RESPECT guidance developed by WHO and other agencies assists in prevention and response efforts. RESPECT outlines a range of different strategies to prevent IPV at the individual and couple level(21), as well as proven interventions for communities. Cross-cutting ingredients for effective programming include addressing multiple risk factors, supporting survivors, engaging with both women and men, and addressing gender and social empowerment, as well as critical reflection and communication skills(22).

Despite progress testing interventions to prevent and respond to IPV, few interventions have been implemented at scale(23). Notable exceptions are programming which has been integrated to sectors or systems, including education, development or health sectors. For example, cash transfer programs operate at-scale across numerous countries and address a structural driver of violence, poverty, as part of countries’ social protection programming. Cash transfers, with and without complementary programming have been shown to reduce IPV across regions(24, 25). However, few other innovations and interventions are developed and tested with scalability in mind, considering the cost and required human resource needs to reach broad populations. Our findings suggest an urgent need to identify, finance and implement interventions that can change population prevalence of IPV at the country level.

Our findings also strongly point to the need to actively monitor equity in progress, to ‘leave no one behind.’ For women experiencing more severe IPV, tailored interventions may be required which respond to underlying dynamics linked to injury, disability and femicide. Encouragingly, some existing IPV prevention interventions seem to be more effective among the most violent men(26), but further research is needed to understand how our broader evidence base of interventions may be effective or not effective with different types of perpetrators. Our findings also suggest that different subgroups of women and girls according to wealth, education and age may experience changes in prevalence in varying directions across-countries, and therefore that context specific monitoring is needed to ensure equity in IPV reductions.

Finally, our findings have implications for the architecture of data collection and availability of quality data on IPV. Typical gaps between DHS rounds across included countries, with some exceptions, are around five years. We consider these minimal requirements to conduct a trend analysis, as longer periods may obscure increases or decreases. With DHS on hold due to the United States Agency for International Development (USAID) shutdown in 2025(27), there is no other widely available data source to monitor progress towards SDG target 5.2.1. This leaves the global community unaccountable to meet targets and undermines research and learning at the national level. A solution is urgently needed to restart funding for collection of this data, both to enable continued monitoring and to avoid the loss of technical capacity on how to rigorously and ethically collect IPV data at scale. Progress in reducing IPV against women cannot be assumed. Ending violence against women will require sustained investment, tailored interventions, and rigorous evidence generation across country contexts.

## Acknowledgments

We thank Sarah LaPointe for initial analysis to assess feasibility of this paper while a Ph.D. student at the University of Buffalo.

## Funding

This study was funded by an anonymous donor to the London School of Hygiene and Tropical Medicine. AP, CC and TP received no funding for work on this study.

## Author Contributions

Conceptualization: AP, TP, KD

Data curation: MM

Methodology: MM, AP, TP

Formal analysis: MM

Funding acquisition: KD

Project administration: KD, AP

Visualization: MM

Writing – original draft writing: MM, KD

Writing – review & editing: AP, TP, CC, KD, MM

## Ethics

Ethical approval was obtained from the London School of Hygiene and Tropical Medicine ethics committee (#32116). Each DHS obtained the required and appropriate country-specific ethical approval for primary data collection.

## Declaration of Interests

AP works for UNICEF, a global organization that works on violence against women, girls and IPV programming, however the study work pre-dates her employment; As such, UNICEF had no role in this study and no influence over results or the decision to publish; Results do not reflect the views or opinions of UNICEF; All other authors declare that they have no competing interests.

## Data Availability Statement

Individual country datasets can be requested through the Demographic and Health Survey Program website (https://dhsprogram.com/). Due to the dismantling of the United States Agency for International Development (USAID), access has been paused during periods of 2025. During these periods of pause, data may be accessed through the individual countries or through repositories like the Integrated Public Use Microdata Series (IPUMS: https://www.ipums.org/). Code generated for this analysis is available upon reasonable request from the corresponding author.

