## Supplementary Materials for "Is intimate partner violence decreasing? Analysis of population-level trends in 21 countries"

**Tables:**

**Table S1: Definitions of intimate partner violence outcomes, including harmonisation notes**

| | Outcome | Numerator | denominator | Notes |
|---|---|---|---|---|
| 1 | Recent experience of Physical IPV (current/most recent partner). | For most recent partner in the last 12 months: yes to any of the following indicators:<br>d105a ever been pushed, shook or had something thrown by husband/partner<br>d105b ever been slapped by husband/partner<br>d105c ever been punched with fist or hit by something harmful by husband/partner<br>d105d ever been kicked or dragged by husband/partner<br>d105e ever been strangled or burnt by husband/partner<br>d105f ever been threatened with knife/gun or other weapon by husband/partner<br>d105g ever been attacked with knife/gun or other weapon by husband/partner<br>d105j ever had arm twisted or hair pulled by husband/partner. See notes for variations. | All women ever in a union aged 15-49 unless otherwise stated | Note for Cambodia 2005, Cameroon 2002, Dominican Republic 2002 and Malawi 2004: questions d105b and d105j were together as d105b "spouse ever slapped or twisted arm" d105j – did not exist. This means that hair pulled is an additional item included in the later surveys which could potentially bias changes in prevalence upwards (see fig. S10). In the Colombian surveys some questions are different to the other surveys for example Colombia 2010 - d105k asks "spouse ever bitten."<br><br>Note that category values for datasets in recode phase 4 (Cambodia 2005, Cameroon 2002, Dominican Republic 2002 and Malawi 2004) are not the same as those in phase 5 onwards. To create a variable that allows for whether the IPV occurred in the last 12 months or not the individual item variables are used in conjunction with an additional variable with the suffix "n" d105an, d105bn etc that gives the frequency in last 12months. |
| 2 | Recent experience of Physical IPV, country specific, ensuring consistent items used across surveys (current/most recent partner). | For most recent partner in the last 12 months:<br>As above (1), but items used for recent of Physical IPV are restricted to allow for harmonisation across surveys; the restricted subset of items are as follows: Kenya, Tanzania, Zambia to d105e and 105j ; Cambodia, Cameroon, Dominican Republic, Haiti, Jordan (not 2007), Malawi, Mozambique, Nepal, Nigeria, Philippines, Senegal, Uganda, Zimbabwe d105a- d105f and d105j ; Peru to d105g ; Colombia d105a, d105c- d105g ; Rwanda d105a- d105c, d105f and d105j. | All women ever in a union aged 15-49 unless otherwise stated | See Note (1) and also note on Rwanda in supplementary materials page 9 and fig. S1. |
| 3 | Recent experience of sexual IPV (current/most recent partner). | For most recent partner in the last 12 months:<br>For most recent partner Yes to any - d105h ever been physically forced into unwanted sex by husband/partner<br>d105i ever been forced into other unwanted sexual acts by husband/partner<br>d105k ever been physically forced to perform sexual acts respondent didn't want to by husband/partner | All women ever in a union aged 15-49 unless otherwise stated | Note for Cambodia2005, Cameroon2002, Dominican Republic2002 and Malawi 2004 - d105i spouse ever forced other sexual acts when not wanted so incorporates d105i and d105k in one. There are additional questions on experience of sexual IPV asked in some DHS, but they are not standard across surveys within a country so have not been listed here. Colombia 2010 - |

| | | | | |
|---|---|---|---|---|
| | | | | d105k is different and asks: "spouse ever bitten" and not "ever been physically forced to perform sexual acts respondent didn't want to by husband/partner" so is not included. |
| 4 | Recent experience of sexual IPV, country specific, ensuring consistent items used across surveys (current/most recent partner). | For most recent partner in the last 12 months:<br>As above (3) but items used for lifetime experience of Sexual IPV are restricted to allow for harmonisation across surveys for all countries d105i, d105i and d105k are used except Cameroon, Rwanda, Colombia, Jordan where only d105h is used. | All women ever in a union aged 15-49 unless otherwise stated | See note (3) |
| 5 | Recent experience of physical and/or sexual IPV | Combining (1) and (3) | All women ever in a union aged 15-49 unless otherwise stated | None |
| 6 | Recent experience of Physical or Sexual IPV, country specific, ensuring consistent items used across surveys | Combining (2) and (4) | All women ever in a union aged 15-49 unless otherwise stated | None |
| 7 | Frequency of Physical and/or Sexual IPV | Using the same items as (6), score 1 for sometimes 2 for often. | All women ever in a union aged 15-49 unless otherwise stated, who have experienced recent physical IPV | For frequency of IPV Cameroon 2004, Malawi 2004, Mali 2006, Rwanda 2005 collected frequency differently, so these surveys are excluded from the frequency score analysis. |
| 8 | Severity of Physical IPV | For most recent partner in the last 12 months for women who have experience recent physical IPV (2): yes to any of the following indicators:<br>d105d ever been kicked or dragged by husband/partner<br>d105e ever been strangled or burnt by husband/partner<br>d105f ever been threatened with knife/gun or other weapon by husband/partner | All women ever in a union aged 15-49 unless otherwise stated who have experienced recent physical IPV | None |

**Table S2: List of all DHS with a domestic violence module, study inclusion and comparison to those used in the trend analysis by Ma et al 2023[a]**

| Region and country | Survey | Recode phase | Included | Used in Ma et al 2023[a] for trend analyses | Reason for exclusion from this study |
|---|---|---|---|---|---|
| **African Region** | | | | | |
| **Angola** | | | | | |
| | 2015-16 | 7 | No | No | Only one survey in country |
| **Benin** | | | | | |
| | 2017-18 | 7 | No | No | Only one survey in country |
| **Burkina Faso** | | | | | |
| | 2010 | 6 | No | No | Only two surveys in country |
| | 2021 | 8 | No | No | Only two surveys in country |
| **Burundi** | | | | | |
| | 2016-17 | 7 | No | No | Only one survey in country |
| **Cameroon** | | | | | |
| | 2004 | 4 | Yes | No | |
| | 2011 | 6 | Yes | Yes | |
| | 2018 | 7 | Yes | Yes | |
| **Chad** | | | | | |
| | 2014-15 | 7 | No | No | Only one survey in country |
| **Comoros** | | | | | |
| | 2012 | 6 | No | No | Only one survey in country |
| **Cote d'Ivoire** | | | | | |
| | 2011-12 | 7 | No | No | Only two surveys in country |
| | 2021 | 8 | No | No | Only two surveys in country |
| **DR Congo** | | | | | |
| | 2007 | 5 | No | Yes | Only two surveys in country |
| | 2013-14 | 6 | No | Yes | Only two surveys in country |
| **Ethiopia** | | | | | |
| | 2016 | 7 | No | | Only one survey in country |
| **Gabon** | | | | | |
| | 2012 | 6 | No | No | Only two surveys in country |
| | 2019-22 | 7 | No | No | Only two surveys in country |
| **Gambia** | | | | | |
| | 2013 | 6 | No | Yes | Only two surveys in country |
| | 2019-2020 | 7 | No | Yes | Only two surveys in country |
| **Ghana** | | | | | |
| | 2008 | 5 | No | No | Only two surveys in country |
| | 2022 | 8 | No | No | Only two surveys in country |
| **Kenya** | | | | | |
| | 2003 | 4 | No | Yes | Incompatible with later surveys* |
| | 2008-09 | 5 | Yes | No | |
| | 2014 | 6 | Yes | Yes | |
| | 2022 | 8 | Yes | No | |
| **Liberia** | | | | | |
| | 2007 | 5 | No | Yes | Only two surveys in country |
| | 2019-2020 | 7 | No | Yes | Only two surveys in country |
| **Madagascar** | | | | | |
| | 2021 | 7 | No | No | Only one survey in country |
| **Malawi** | | | | | |
| | 2004 | 4 | Yes | No | |
| | 2010 | 5 | Yes | No | |
| | 2015-16 | 7 | Yes | No | |
| **Mali** | | | | | |
| | 2006 | 5 | Yes | No | |
| | 2012-13 | 6 | Yes | Yes | |
| | 2018 | 7 | Yes | Yes | |

*Table S2 continued…*

| Region and country | Survey | Recode phase | Included | Used in Ma et al 2023 [a] for trend analyses | Reason for exclusion from this study |
|---|---|---|---|---|---|
| **African Region continued** | | | | | |
| **Mozambique** | | | | | |
| | 2011 | 6 | Yes | No | |
| | 2015 | 6 | Yes | No | |
| | 2022-23 | 8 | Yes | No | |
| **Namibia** | | | | | |
| | 2013 | 6 | No | No | Only one survey in country |
| **Nigeria** | | | | | |
| | 2008 | 5 | Yes | Yes | |
| | 2013 | 6 | Yes | No | |
| | 2018 | 7 | Yes | Yes | |
| **Rwanda** | | | | | |
| | 2005 | 4 | No | Yes | Incompatible with later surveys[b] |
| | 2010 | 6 | Yes | No | |
| | 2014-15 | 6 | Yes | No | |
| | 2019-20 | 7 | Yes | Yes | |
| **Sao Tome and Principe** | | | | | |
| | 2008-09 | 5 | No | No | Only one survey in country |
| **Senegal** | | | | | |
| | 2017 | 7 | Yes | No | |
| | 2018 | 7 | Yes | No | |
| | 2019 | 7 | Yes | No | |
| **Sierra Leone** | | | | | |
| | 2013 | 6 | No | Yes | Only two surveys in country |
| | 2019 | 7 | No | Yes | Only two surveys in country |
| **South Africa** | | | | | |
| | 2016 | 7 | No | No | Only one survey in country |
| **Tanzania** | | | | | |
| | 2010 | 5 | Yes | Yes | |
| | 2015-16 | 7 | Yes | Yes | |
| | 2022 | 8 | Yes | No | |
| **Togo** | | | | | |
| | 2013-14 | 6 | No | No | Only one survey in country |
| **Uganda** | | | | | |
| | 2006 | 5 | Yes | Yes | |
| | 2011 | 6 | Yes | No | |
| | 2016 | 7 | Yes | Yes | |
| **Zambia** | | | | | |
| | 2001-02 | 4 | No | No | Incompatible with later surveys[b] |
| | 2007 | 5 | Yes | Yes | |
| | 2013-14 | 6 | Yes | No | |
| | 2018 | 7 | Yes | Yes | |
| **Zimbabwe** | | | | | |
| | 2005-05 | 5 | Yes | Yes | |
| | 2010-11 | 6 | Yes | No | |
| | 2015 | 7 | Yes | Yes | |
| **Region of the Americas** | | | | | |
| **Bolivia** | | | | | |
| | 2003 | 4 | No | No | Only two surveys (Domestic violence variables are |
| | 2008 | 5 | No | No | Only two surveys (Domestic violence variables not |
| **Colombia** | | | | | |
| | 2000 | 4 | No | No | Incompatible with later surveys[b] |
| | 2005 | 4 | Yes | No | |
| | 2010 | 5 | Yes | No | |
| | 2015 | 7 | Yes | No | |

*Table S2 continued…*

| Region and country | Survey | Recode phase | Included | Used in Ma et al 2023 [a] for trend analyses | Reason for exclusion from this study |
|---|---|---|---|---|---|
| **Region of the Americas Region continued** | | | | | |
| **Dominican Republic** | | | | | |
| | 2002 | 4 | Yes | Yes | |
| | 2007 | 6 | Yes | No | |
| | 2013 | 6 | Yes | Yes | |
| **Guatemala** | | | | | |
| | 2014-15 | 7 | No | No | Only one survey in country |
| **Haiti** | | | | | |
| | 2000 | 4 | No | Yes | Incompatible with later surveys[b] |
| | 2005-06 | 5 | Yes | No | |
| | 2012 | 6 | Yes | No | |
| | 2016-17 | 7 | Yes | Yes | |
| **Honduras** | | | | | |
| | 2005 | | No | No | Only two surveys in country |
| | 2011-12 | | No | No | Only two surveys in country |
| **Peru** | | | | | |
| | 2000 | 4 | No | No | Incompatible with later surveys[b] |
| | 2004-08 | 5 | Yes | No | |
| | 2009 | 5 | Yes | No | |
| | 2010 | 5 | Yes | No | |
| | 2011 | 5 | Yes | No | |
| | 2012 | 5 | Yes | No | |
| **South-East Asian Region** | | | | | |
| **Cambodia** | | | | | |
| | 2000 | 4 | No | Yes | Incompatible with later surveys[b] |
| | 2005 | 5 | Yes | No | |
| | 2014 | 6 | Yes | Yes | |
| | 2021-22 | 8 | Yes | No | |
| **India** | | | | | |
| | 2005-06 | 5 | Yes | Yes | |
| | 2015-16 | 6 | Yes | No | |
| | 2019-21 | 7 | Yes | Yes | |
| **Maldives** | | | | | |
| | 2016-17 | 7 | No | No | Only one survey in country |
| **Myanmar** | | | | | |
| | 2015-16 | 7 | No | No | Only one survey in country |
| **Nepal** | | | | | |
| | 2011 | 6 | Yes | No | |
| | 2016 | 7 | Yes | No | |
| | 2022 | 8 | Yes | No | |
| **Timor Leste** | | | | | |
| | 2009-10 | 5 | No | Yes | Only two surveys in country |
| | 2016 | 7 | No | Yes | Only two surveys in country |
| **European Region** | | | | | |
| **Armenia** | | | | | |
| | 2015-16 | 7 | No | No | Only one survey in country |
| **Azerbaijan** | | | | | |
| | 2006 | 5 | No | No | Only one survey in country |
| **Kyrgyz Republic** | | | | | |
| | 2012 | 6 | No | No | Only one survey in country |
| **Moldova** | | | | | |
| | 2005 | 4 | No | No | Only one survey in country |
| **Tajikistan** | | | | | |
| | 2012 | 6 | No | Yes | Only two surveys in country |
| | 2017 | 7 | No | Yes | Only two surveys in country |
| **Ukraine** | | | | | |
| | 2007 | 5 | No | No | Only one survey in country |

*Table S2 continued…*

| Region and country | | Survey | Recode phase | Included | Used in Ma et al 2023 [a] for trend analyses | Reason for exclusion from this study |
|---|---|---|---|---|---|---|
| **Eastern Mediterranean Region** | | | | | | |
| | **Afghanistan** | | | | | |
| | | 2015 | 7 | No | No | Only one survey in country |
| | **Egypt** | | | | | |
| | | 2005 | 4 | No | No | Only two surveys in country (this is also |
| | | 2014 | 6 | No | No | Only two surveys in country |
| | **Jordan** | | | | | |
| | | 2007 | 5 | Yes | Yes | |
| | | 2012 | 6 | Yes | No | |
| | | 2017-18 | 7 | Yes | Yes | |
| | | 2023 | 8 | Yes | No | |
| | **Mauritania** | | | | | |
| | | 2019-21 | 7 | No | No | Only one survey in country |
| | **Pakistan** | | | | | |
| | | 2012-13 | 6 | No | No | Only two surveys in country |
| | | 2017-18 | 7 | No | No | Only two surveys in country |
| | **Western Pacific Region** | | | | | |
| | **Philippines** | | | | | |
| | | 2008 | 5 | Yes | Yes | |
| | | 2013 | 6 | Yes | No | |
| | | 2017 | 7 | Yes | Yes | |
| | | 2022 | 8 | Yes | No | |

[a] Ma et al, Prevalence and changes of intimate partner violence against women aged 15-49 years in 53 low-income and middle-income countries from 2000 to 2021: a secondary analysis of population-based data. Lancet Glob Health 2023; 11: e1863-73

[b]In order to include these surveys we would have had to restrict all surveys within the country to a very small set of items to standardise, therefore limiting the usability of later surveys.

### A note on Rwanda

For Rwanda 2010 it was clear that there was an anomaly causing an unrealistic spike in the experience of physical and/or sexual IPV (figure s1). The questions were comparable when looking at the individual items across the surveys, however there was a huge change in the proportion in the 2010 DHS for d105d ever been kicked or dragged by husband/partner; where the prevalence was below 10% for the other three surveys but 40% in 2010 and for d105e, ever been strangled or burnt by husband/partner, the prevalence was well below 5% for the other three surveys but around 25% for 2010. As it is infeasible that an "ever" measure should change so drastically across cohorts, we therefore assumed that there had been issues with these questions and excluded them from the harmonised measures of physical and/or sexual IPV.

**Figure S1: Proportion of ever in a union (excluding widowed women) answering yes to each item on their experience of physical and/or sexual IPV by survey in Rwanda.**

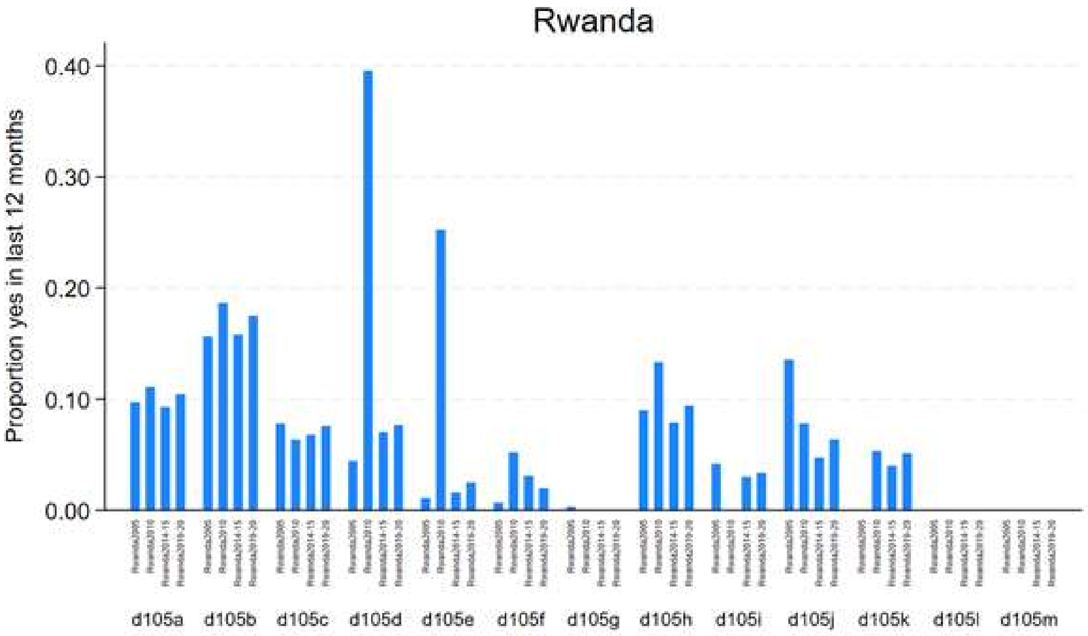

**Table S3: Summary of surveys and samples used across countries**

| Country | Survey | Domestic violence module sample | Unweighted sample 15-49, ever in a union[1] | Sample used for all analyses[1] |
|---|---|---|---|---|
| **African Region** | | | | |
| **Cameroon** | | | | |
| | 2004 | Women 15-49, Widows excluded | 2673 | 15-49, ever in a union, widows excluded |
| | 2011 | Women 15-49 Widows excluded from questions on recent ipv | 4006 | 15-49, ever in a union, widows excluded |
| | 2018 | Women 15-49 | 4690 | 15-49, ever in a union, widows excluded |
| **Kenya** | | | | |
| | 2008-09 | Women 15-49 | 4906 | 15-49, ever in a union |
| | 2014 | Women 15-49 | 4519 | 15-49, ever in a union |
| | 2022 | Women 15-49 | 12888 | 15-49, ever in a union |
| **Malawi** | | | | |
| | 2004 | Women 15-49, widows excluded | 8631 | 15-49, ever in a union, widows excluded |
| | 2010 | Women 15-49 | 5380 | 15-49, ever in a union, widows excluded |
| | 2015-16 | Women 15-49 | 5406 | 15-49, ever in a union, widows excluded |
| **Mali** | | | | |
| | 2006 | Widows 15-49 excluded | 9075 | 15-49, ever in a union, widows excluded |
| | 2012-13 | Women 15-49 | 3120 | 15-49, ever in a union, widows excluded |
| | 2018 | Women 15-49 | 3356 | 15-49, ever in a union, widows excluded |
| **Mozambique** | | | | |
| | 2011 | Women 15-49 | 5824 | 15-49, ever in a union |
| | 2015 | Women 15-59 | 2881 | 15-49, ever in a union |
| | 2022-23 | Women 15-49 | 3992 | 15-49, ever in a union |
| **Nigeria** | | | | |
| | 2008 | Women 15-49 Widows excluded from questions on recent IPV | 19389 | 15-49, ever in a union |
| | 2013 | Women 15-49 | 22305 | 15-49, ever in a union |
| | 2018 | Women 15-49 | 8910 | 15-49, ever in a union |
| **Rwanda** | | | | |

| | | | | |
|---|---|---|---|---|
| | 2010 | Women 15-49 | 3476 | 15-49, ever in a union |
| | 2014-15 | Women 15-49 | 1908 | 15-49, ever in a union |
| | 2019-20 | Women 15-49 | 1947 | 15-49, ever in a union |
| **Senegal** | | | | |
| | 2017 | Women 15-49 | 2660 | 15-49, ever in a union |
| | 2018 | Women 15-49 | 1506 | 15-49, ever in a union |
| | 2019 | Women 15-49 | 1468 | 15-49, ever in a union |
| **Tanzania** | | | | |
| | 2010 | Women 15-49 | 5689 | 15-49, ever in a union |
| | 2015-16 | Women 15-49 | 7597 | 15-49, ever in a union |
| | 2022 | Women 15-49 | 4367 | 15-49, ever in a union |
| **Uganda** | | | | |
| | 2006 | Women 15-49 Widows excluded from questions on recent IPV | 1749 | 15-49, ever in a union, widows excluded |
| | 2011 | Women 15-49 Widows excluded from questions on recent IPV | 1705 | 15-49, ever in a union, widows excluded |
| | 2016 | Women 15-49 | 7536 | 15-49, ever in a union, widows excluded |
| **Zambia** | | | | |
| | 2007 | Women 15-49 | 4246 | 15-49, ever in a union |
| | 2013-14 | Women 15-49 | 9416 | 15-49, ever in a union |
| | 2018 | Women 15-49 | 7358 | 15-49, ever in a union |
| **Zimbabwe** | | | | |
| | 2005-06 | Women 15-49 | 4994 | 15-49, ever in a union |
| | 2010-11 | Women 15-49 | 5282 | 15-49, ever in a union |
| | 2015 | Women 15-49 | 5800 | 15-49, ever in a union |
| **Region of the Americas** | | | | |
| **Colombia** | | | | |
| | 2005 | Women 13-49 | 25620 | 15-49, ever in a union |
| | 2010 | Women 13-49 | 34624 | 15-49, ever in a union |
| | 2015 | Women 13-49 | 24862 | 15-49, ever in a union |
| **Dominican Republic** | | | | |

| | | | |
|---|---|---|---|
| 2002 | Women 15-49 | 7435 | 15-49, ever in a union |
| 2007 | Women 15-49 | 9003 | 15-49, ever in a union |
| 2013 | Women 15-49 | 5803 | 15-49, ever in a union |
| **Haiti** | | | |
| 2005-06 | Women 15-49 Widows excluded from questions on recent IPV | 2680 | 15-49, ever in a union, widows excluded |
| 2012 | Women 15-49 | 6650 | 15-49, ever in a union, widows excluded |
| 2016-17 | Women 15-49 | 4322 | 15-49, ever in a union, widows excluded |
| **Peru** | | | |
| 2004-06 | Women 15-49 | 10354 | 15-49, ever in a union |
| 2007-08 | Women 15-49 | 12572 | 15-49, ever in a union |
| 2009 | Women 15-49 | 13781 | 15-49, ever in a union |
| 2010 | Women 15-49 | 12880 | 15-49, ever in a union |
| 2011 | Women 15-49 | 12898 | 15-49, ever in a union |
| 2012 | Women 15-49 | 13483 | 15-49, ever in a union |
| **South-East Asian Region** | | | |
| **Cambodia** | | | |
| 2005 | Women 15-49 | 2294 | 15-49, ever in a union |
| 2014 | Women 15-49 | 3499 | 15-49, ever in a union |
| 2021-22 | Women 15-49 | 6062 | 15-49, ever in a union |
| **India** | | | |
| 2005-06 | Women 15-49 Widows excluded from questions on recent IPV | 69484 | 18-49, ever in a union, widows excluded |
| 2015-16 | Women 15-49 | 66013 | 18-49, ever in a union, widows excluded |
| 2019-21 | Women 18-49 | 63851 | 18-49, ever in a union, widows excluded |
| **Nepal** | | | |
| 2011 | Women 15-49 | 3505 | 15-49, ever in a union |
| 2016 | Women 15-49 | 3826 | 15-49, ever in a union |
| 2022 | Women 15-49 | 4377 | 15-49, ever in a union |
| **Eastern Mediterranean Region** | | | |
| **Jordan** | | | |
| 2007 | Ever married women 15-49 | 3444 | 15-49, ever married |

| | | | |
|---|---|---|---|
| 2012 | Ever married women 15-49 | 7027 | 15-49, ever married |
| 2017-18 | Ever married women 15-49 | 6852 | 15-49, ever married |
| 2023 | Ever married women 15-49 | 5495 | 15-49, ever married |
| **Western Pacific Region** | | | |
| **Philippines** | | | |
| 2008 | Women 15-49 | 7157 | 15-49, ever in a union |
| 2013 | Women 15-49 | 8160 | 15-49, ever in a union |
| 2017 | Women 15-49 | 13215 | 15-49, ever in a union |
| 2022 | Women 15-49 | 12906 | 15-49, ever in a union |

[1]Ever in a union is defined as ever cohabited or ever married.

**Table S4: Distribution of domestic violence sample by marital status and reported number of unions for women aged 15-49.**

| Country | Survey | Marital status | | | | | | Number of unions for ever married women | | | | | |
|---|---|---|---|---|---|---|---|---|---|---|---|---|---|
| | | Never married | | Married/cohabitating | | Divorced/separated/ widowed | | Once | | More than once | | Unknown | |
| | | % | 95%CI | % | 95%CI | % | 95%CI | % | 95%CI | % | 95%CI | % | 95%CI |
| African Region | | | | | | | | | | | | | |
| Cameroon | | | | | | | | | | | | | |
| | 2004 | 25.4 | (23.3-27.8) | 65.6 | (63.3-67.9) | 8.9 | (7.7-10.2) | 77.7 | (75.6-79.7) | 22.3 | (20.3-24.4) | . | ( .- .) |
| | 2011 | 26.8 | (24.9-28.8) | 64.3 | (62.1-66.4) | 8.9 | (7.8-10.2) | 79.5 | (77.5-81.4) | 20.5 | (18.6-22.5) | . | ( .- .) |
| | 2018 | 35.6 | (33.7-37.5) | 54.9 | (53.0-56.8) | 9.5 | (8.6-10.6) | 88.5 | (87.2-89.7) | 11.5 | (10.3-12.8) | . | ( .- .) |
| Kenya | | | | | | | | | | | | | |
| | 2008-09 | 31.4 | (29.2-33.6) | 58.4 | (56.3-60.4) | 10.3 | (9.0-11.7) | 94.0 | (92.9-94.8) | 5.8 | (5.0-6.9) | 0.2 | (0.1-0.7) |
| | 2014 | 28.9 | (26.9-30.9) | 59.3 | (57.2-61.3) | 11.9 | (10.6-13.2) | 92.8 | (91.9-93.7) | 7.2 | (6.3-8.1) | . | ( .- .) |
| | 2022 | 32.3 | (31.1-33.5) | 56.1 | (54.9-57.3) | 11.6 | (11.0-12.3) | 92.7 | (92.1-93.4) | 7.3 | (6.6-7.9) | . | ( .- .) |
| Malawi | | | | | | | | | | | | | |
| | 2004 | 17.0 | (15.7-18.3) | 70.7 | (69.0-72.3) | 12.3 | (11.4-13.3) | 77.5 | (76.2-78.7) | 22.5 | (21.3-23.8) | . | ( .- .) |
| | 2010 | 18.9 | (17.3-20.5) | 68.0 | (66.3-69.7) | 13.1 | (12.1-14.3) | 75.8 | (74.3-77.3) | 23.9 | (22.4-25.5) | 0.3 | (0.1-0.6) |
| | 2015-16 | 21.9 | (20.1-23.7) | 65.4 | (63.4-67.3) | 12.7 | (11.5-14.1) | 77.8 | (76.1-79.5) | 22.2 | (20.5-23.9) | . | ( .- .) |
| Mali | | | | | | | | | | | | | |
| | 2006 | 12.2 | (10.8-13.7) | 84.4 | (82.8-85.9) | 3.4 | (2.9-4.0) | 83.9 | (82.3-85.3) | 16.1 | (14.7-17.7) | . | ( .- .) |
| | 2012-13 | 14.7 | (12.8-16.8) | 83.4 | (81.3-85.3) | 1.9 | (1.4-2.7) | 93.4 | (92.0-94.6) | 6.6 | (5.4-8.0) | . | ( .- .) |
| | 2018 | 14.7 | (12.9-16.7) | 82.7 | (80.7-84.6) | 2.6 | (2.0-3.4) | 92.7 | (91.5-93.8) | 7.3 | (6.2-8.5) | . | ( .- .) |
| Mozambique | | | | | | | | | | | | | |
| | 2011 | 17.9 | (16.5-19.4) | 68.2 | (66.5-69.8) | 13.9 | (12.8-15.1) | 79.7 | (78.1-81.2) | 20.3 | (18.8-21.9) | . | ( .- .) |
| | 2015 | 9.8 | (8.5-11.2) | 70.9 | (68.5-73.2) | 19.3 | (17.6-21.2) | 78.9 | (76.7-81.0) | 21.1 | (19.0-23.3) | . | ( .- .) |
| | 2022-23 | 20.6 | (18.9-22.4) | 65.3 | (63.3-67.3) | 14.1 | (12.8-15.5) | 79.1 | (77.2-80.9) | 20.9 | (19.1-22.8) | . | ( .- .) |
| Nigeria | | | | | | | | | | | | | |
| | 2008 | 21.9 | (20.9-23.0) | 73.8 | (72.7-75.0) | 4.2 | (3.9-4.6) | 86.8 | (86.1-87.5) | 13.2 | (12.5-13.9) | . | ( .- .) |
| | 2013 | 23.3 | (22.2-24.4) | 72.1 | (70.9-73.2) | 4.6 | (4.3-5.0) | 88.5 | (87.8-89.2) | 10.5 | (9.8-11.1) | 1.0 | (0.8-1.2) |
| | 2018 | 21.7 | (20.4-23.1) | 73.5 | (72.1-74.8) | 4.8 | (4.3-5.3) | 89.1 | (88.2-90.0) | 10.9 | (10.0-11.8) | . | ( .- .) |

***Table S4 continued…:***

| Country | Survey | Never married % | Never married 95%CI | Married/cohabitating % | Married/cohabitating 95%CI | Divorced/separated/ widowed % | Divorced/separated/ widowed 95%CI | Once % | Once 95%CI | More than once % | More than once 95%CI | Unknown % | Unknown 95%CI |
|---|---|---|---|---|---|---|---|---|---|---|---|---|---|
| | | Marital status | | | | | | Number of unions for ever married women | | | | | |
| Rwanda | | | | | | | | | | | | | |
| | 2010 | 39.3 | (37.6-40.9) | 49.9 | (48.3-51.5) | 10.8 | (9.8-11.9) | 86.3 | (85.0-87.5) | 13.6 | (12.4-14.9) | 0.1 | (0.0-0.4) |
| | 2014-15 | 36.9 | (34.6-39.3) | 52.8 | (50.4-55.2) | 10.3 | (8.9-11.9) | 88.0 | (86.3-89.6) | 12.0 | (10.4-13.7) | . | ( .- .) |
| | 2019-20 | 38.9 | (36.6-41.3) | 51.3 | (49.0-53.6) | 9.8 | (8.5-11.2) | 89.8 | (88.1-91.3) | 10.2 | (8.7-11.9) | . | ( .- .) |
| Senegal | | | | | | | | | | | | | |
| | 2017 | 30.8 | (28.3-33.3) | 63.6 | (61.0-66.1) | 5.7 | (4.4-7.3) | 86.8 | (85.0-88.4) | 13.2 | (11.6-15.0) | . | ( .- .) |
| | 2018 | 28.4 | (25.8-31.1) | 65.8 | (62.5-68.9) | 5.8 | (4.0-8.3) | 90.3 | (88.0-92.2) | 9.7 | (7.8-12.0) | . | ( .- .) |
| | 2019 | 26.6 | (23.4-30.1) | 69.0 | (65.5-72.2) | 4.4 | (3.2-6.2) | 88.8 | (86.5-90.8) | 11.2 | (9.2-13.5) | . | ( .- .) |
| Tanzania | | | | | | | | | | | | | |
| | 2010 | 25.0 | (23.4-26.6) | 63.4 | (61.7-65.1) | 11.6 | (10.6-12.7) | 78.6 | (77.0-80.1) | 21.0 | (19.5-22.6) | 0.4 | (0.2-0.8) |
| | 2015-16 | 23.8 | (22.4-25.3) | 63.0 | (61.4-64.6) | 13.2 | (12.2-14.3) | 81.8 | (80.3-83.2) | 18.2 | (16.8-19.7) | . | ( .- .) |
| | 2022 | 26.8 | (24.9-28.8) | 60.1 | (57.9-62.2) | 13.1 | (11.9-14.4) | 86.8 | (85.4-88.1) | 13.2 | (11.9-14.6) | . | ( .- .) |
| Uganda | | | | | | | | | | | | | |
| | 2006 | 23.4 | (20.7-26.3) | 62.5 | (59.4-65.5) | 14.1 | (12.1-16.3) | 76.9 | (74.4-79.2) | 23.1 | (20.8-25.6) | 0.0 | ( .- .) |
| | 2011 | 22.8 | (20.2-25.5) | 63.6 | (60.7-66.4) | 13.6 | (11.9-15.6) | 79.7 | (77.3-82.0) | 20.3 | (18.0-22.7) | 0.0 | ( .- .) |
| | 2016 | 25.5 | (24.2-26.8) | 61.1 | (59.6-62.6) | 13.4 | (12.5-14.4) | 80.9 | (79.6-82.1) | 19.1 | (17.9-20.4) | . | ( .- .) |
| Zambia | | | | | | | | | | | | | |
| | 2007 | 25.3 | (23.2-27.6) | 62.1 | (59.9-64.3) | 12.6 | (11.3-13.9) | 81.1 | (79.5-82.6) | 18.8 | (17.3-20.4) | 0.1 | (0.0-0.2) |
| | 2013-14 | 27.2 | (26.0-28.5) | 60.7 | (59.2-62.1) | 12.1 | (11.2-13.1) | 81.7 | (80.5-82.8) | 18.3 | (17.2-19.5) | . | ( .- .) |
| | 2018 | 30.6 | (28.8-32.4) | 56.7 | (55.0-58.3) | 12.8 | (11.8-13.9) | 82.9 | (81.4-84.3) | 17.1 | (15.7-18.6) | . | ( .- .) |
| Zimbabwe | | | | | | | | | | | | | |
| | 2005-05 | 26.0 | (24.5-27.5) | 58.7 | (57.1-60.3) | 15.3 | (14.2-16.5) | 85.5 | (84.1-86.8) | 14.5 | (13.2-15.9) | . | ( .- .) |
| | 2010-11 | 23.3 | (21.8-24.9) | 62.6 | (60.8-64.4) | 14.1 | (13.1-15.1) | 85.1 | (83.7-86.4) | 14.9 | (13.6-16.3) | . | ( .- .) |
| | 2015 | 23.9 | (22.3-25.7) | 63.6 | (61.8-65.3) | 12.5 | (11.5-13.5) | 83.5 | (82.2-84.7) | 16.5 | (15.3-17.8) | . | ( .- .) |

***Table S4 continued…:***

| Country | Survey | Marital status | | | | | | Number of unions for ever married women | | | | | |
|---|---|---|---|---|---|---|---|---|---|---|---|---|---|
| | | Never married | | Married/cohabitating | | Divorced/separated/ widowed | | Once | | More than once | | Unknown | |
| | | % | 95%CI | % | 95%CI | % | 95%CI | % | 95%CI | % | 95%CI | % | 95%CI |
| Region of the Americas | | | | | | | | | | | | | |
| Colombia | | | | | | | | | | | | | |
| | 2005 | 33.1 | (32.4-33.9) | 51.2 | (50.4-51.9) | 15.7 | (15.2-16.2) | 81.6 | (80.9-82.3) | 18.4 | (17.7-19.1) | . | ( .- .) |
| | 2010 | 31.9 | (31.3-32.5) | 52.4 | (51.8-53.0) | 15.7 | (15.3-16.2) | 78.1 | (77.5-78.7) | 21.9 | (21.3-22.5) | . | ( .- .) |
| | 2015 | 31.6 | (30.6-32.6) | 52.7 | (51.8-53.7) | 15.7 | (15.0-16.5) | 75.6 | (74.7-76.5) | 24.4 | (23.5-25.3) | . | ( .- .) |
| Dominican Republic | | | | | | | | | | | | | |
| | 2002 | 22.2 | (20.6-23.8) | 60.2 | (58.5-62.0) | 17.6 | (16.4-18.9) | 63.6 | (61.7-65.4) | 36.4 | (34.6-38.3) | . | ( .- .) |
| | 2007 | 23.4 | (21.7-25.1) | 57.8 | (55.8-59.8) | 18.8 | (17.5-20.3) | 58.9 | (57.0-60.7) | 38.2 | (36.4-40.0) | 2.9 | (2.2-3.8) |
| | 2013 | 24.1 | (22.5-25.9) | 54.2 | (52.1-56.2) | 21.7 | (20.1-23.4) | 60.8 | (59.2-62.3) | 39.2 | (37.7-40.8) | . | ( .- .) |
| Haiti | | | | | | | | | | | | | |
| | 2005-06 | 32.1 | (29.9-34.4) | 58.4 | (55.8-60.8) | 9.5 | (8.3-10.8) | 61.0 | (58.1-63.8) | 38.8 | (36.0-41.7) | 0.2 | (0.1-0.5) |
| | 2012 | 37.5 | (35.8-39.2) | 54.2 | (52.6-55.8) | 8.3 | (7.5-9.3) | 73.0 | (71.0-74.8) | 26.9 | (25.1-28.9) | 0.1 | (0.0-0.3) |
| | 2016-17 | 39.6 | (37.7-41.6) | 52.2 | (50.3-54.1) | 8.1 | (7.1-9.3) | 74.5 | (72.5-76.5) | 25.5 | (23.5-27.5) | . | ( .- .) |
| Peru | | | | | | | | | | | | | |
| | 2004-06 | 28.1 | (26.9-29.3) | 62.3 | (61.1-63.6) | 9.5 | (8.8-10.3) | 90.3 | (89.5-91.1) | 9.7 | (8.9-10.5) | . | ( .- .) |
| | 2007-08 | 27.7 | (26.5-29.0) | 62.8 | (61.5-64.2) | 9.4 | (8.8-10.2) | 88.0 | (87.1-88.8) | 12.0 | (11.2-12.9) | . | ( .- .) |
| | 2009 | 26.5 | (25.5-27.5) | 64.4 | (63.3-65.5) | 9.1 | (8.5-9.7) | 78.3 | (77.3-79.3) | 10.3 | (9.6-11.0) | 11.4 | (10.6-12.3) |
| | 2010 | 26.5 | (25.4-27.5) | 63.2 | (62.1-64.4) | 10.3 | (9.7-11.0) | 88.0 | (87.1-88.8) | 12.0 | (11.2-12.9) | . | ( .- .) |
| | 2011 | 25.8 | (24.8-26.8) | 63.5 | (62.4-64.5) | 10.7 | (10.1-11.3) | 88.4 | (87.7-89.1) | 11.6 | (10.9-12.3) | . | ( .- .) |
| | 2012 | 25.8 | (24.8-26.7) | 63.9 | (62.8-65.0) | 10.3 | (9.7-11.0) | 87.7 | (86.9-88.5) | 12.3 | (11.5-13.1) | . | ( .- .) |

***Table S4 continued…:***

| Country | Survey | Marital status | | | | | | Number of unions for ever married women | | | | | |
|---|---|---|---|---|---|---|---|---|---|---|---|---|---|
| | | Never married | | Married/cohabitating | | Divorced/separated/ widowed | | Once | | More than once | | Unknown | |
| | | % | 95%CI | % | 95%CI | % | 95%CI | % | 95%CI | % | 95%CI | % | 95%CI |
| South-East Asian Region | | | | | | | | | | | | | |
| Cambodia | | | | | | | | | | | | | |
| | 2005 | 29.8 | (27.2-32.5) | 62.0 | (59.3-64.6) | 8.2 | (6.9-9.7) | 91.5 | (89.7-93.0) | 8.0 | (6.6-9.8) | 0.4 | (0.2-0.8) |
| | 2014 | 24.7 | (22.6-26.8) | 69.1 | (67.0-71.2) | 6.2 | (5.4-7.2) | 92.1 | (90.9-93.2) | 7.9 | (6.8-9.1) | . | ( .- .) |
| | 2021-22 | 24.9 | (23.3-26.7) | 68.7 | (67.0-70.3) | 6.4 | (5.5-7.5) | 90.8 | (89.8-91.8) | 9.2 | (8.2-10.2) | . | ( .- .) |
| India | | | | | | | | | | | | | |
| | 2005-06 | 20.4 | (19.9-20.8) | 74.9 | (74.3-75.3) | 4.8 | (4.6-5.0) | 98.0 | (97.8-98.1) | 2.0 | (1.9-2.2) | 0.0 | (0.0-0.1) |
| | 2015-16 | 22.4 | (21.8-22.9) | 73.3 | (72.8-73.9) | 4.3 | (4.1-4.5) | 98.3 | (98.1-98.4) | 1.7 | (1.6-1.9) | . | ( .- .) |
| | 2019-21 | 13.6 | (13.1-14.2) | 81.2 | (80.5-81.8) | 5.2 | (4.9-5.6) | 98.5 | (98.3-98.7) | 1.5 | (1.3-1.7) | . | ( .- .) |
| Nepal | | | | | | | | | | | | | |
| | 2011 | 23.2 | (21.1-25.3) | 73.5 | (71.3-75.6) | 3.3 | (2.7-4.1) | 94.7 | (93.6-95.7) | 5.3 | (4.3-6.4) | . | ( .- .) |
| | 2016 | 19.8 | (18.2-21.6) | 77.6 | (75.8-79.2) | 2.6 | (2.1-3.3) | 96.4 | (95.5-97.1) | 3.6 | (2.9-4.5) | . | ( .- .) |
| | 2022 | 22.1 | (20.6-23.8) | 74.4 | (72.7-76.1) | 3.4 | (2.8-4.2) | 96.4 | (95.7-97.1) | 3.6 | (2.9-4.3) | . | ( .- .) |
| Eastern Mediterranean Region | | | | | | | | | | | | | |
| Jordan | | | | | | | | | | | | | |
| | 2007 | | | 4.6 | (3.5-6.0) | . | ( .- .) | 97.4 | (96.2-98.2) | 2.6 | (1.8-3.8) | 0.0 | (0.0-0.1) |
| | 2012 | | | 4.5 | (3.6-5.4) | . | ( .- .) | 96.5 | (95.5-97.3) | 3.5 | (2.7-4.5) | . | ( .- .) |
| | 2017-18 | | | 6.7 | (5.6-8.0) | . | ( .- .) | 98.1 | (97.5-98.5) | 1.9 | (1.5-2.5) | . | ( .- .) |
| | 2023 | | | 8.6 | (7.3-10.0) | . | ( .- .) | 98.3 | (97.7-98.7) | 1.7 | (1.3-2.3) | . | ( .- .) |
| Western Pacific Region | | | | | | | | | | | | | |
| Philippines | | | | | | | | | | | | | |
| | 2008 | 25.6 | (24.4-26.8) | 70.0 | (68.8-71.2) | 4.4 | (4.0-4.9) | 89.8 | (89.0-90.6) | 8.4 | (7.7-9.1) | 1.8 | (1.4-2.3) |
| | 2013 | 34.5 | (33.3-35.7) | 60.2 | (59.0-61.4) | 5.3 | (4.8-5.9) | 90.4 | (89.6-91.2) | 8.7 | (8.0-9.4) | 0.9 | (0.7-1.2) |
| | 2017 | 35.7 | (34.2-37.2) | 60.0 | (58.5-61.5) | 4.3 | (3.8-4.9) | 89.0 | (88.1-89.8) | 11.0 | (10.2-11.9) | . | ( .- .) |
| | 2022 | 40.9 | (39.7-42.2) | 55.8 | (54.6-57.1) | 3.3 | (2.9-3.7) | 88.5 | (87.6-89.4) | 11.5 | (10.6-12.4) | . | ( .- .) |

**Table S5: Distribution of socio-economic variables by country and survey**

| | Cambodia 2005 | | Cambodia 2014 | | Cambodia 2021-22 | |
|---|---|---|---|---|---|---|
| | % | 95%CI | % | 95%CI | % | 95%CI |
| *Five-year age group* | | | | | | |
| 15-19 | 3.3 | (2.4-4.4) | 3.3 | (2.6-4.3) | 2.4 | (2.0-3.0) |
| 20-24 | 14.9 | (13.1-16.9) | 14.2 | (12.5-16.1) | 10.7 | (9.5-12.0) |
| 25-29 | 15.3 | (13.5-17.3) | 17.8 | (16.1-19.6) | 17.5 | (16.3-18.7) |
| 30-34 | 15.6 | (14.0-17.4) | 22.4 | (20.7-24.2) | 20.8 | (19.5-22.2) |
| 35-39 | 18.3 | (16.3-20.4) | 13.4 | (12.0-14.9) | 20.7 | (19.3-22.2) |
| 40-44 | 18.9 | (16.7-21.4) | 15.7 | (14.0-17.5) | 17.1 | (15.7-18.5) |
| 45-49 | 13.7 | (11.6-16.1) | 13.2 | (11.6-15.0) | 10.9 | (9.7-12.1) |
| *Place of residence* | | | | | | |
| urban | 14.8 | (13.2-16.6) | 15.1 | (13.4-16.9) | 39.6 | (37.4-41.9) |
| rural | 85.2 | (83.4-86.8) | 84.9 | (83.1-86.6) | 60.4 | (58.1-62.6) |
| *Education* | | | | | | |
| None | 25.2 | (22.7-27.9) | 15.3 | (13.4-17.3) | 14.2 | (12.8-15.6) |
| Primary | 58.6 | (55.7-61.4) | 53.2 | (50.8-55.5) | 45.2 | (43.2-47.1) |
| Secondary+ | 16.2 | (14.1-18.6) | 31.6 | (29.2-34.0) | 40.7 | (38.5-42.9) |
| *Wealth* | | | | | | |
| poor/poorest | 40.3 | (37.0-43.7) | 38.2 | (35.2-41.4) | 36.7 | (34.5-39.0) |
| middle | 20.0 | (17.5-22.8) | 20.8 | (18.7-23.2) | 19.8 | (18.3-21.4) |
| rich/richest | 39.7 | (36.5-42.9) | 40.9 | (37.8-44.1) | 43.5 | (41.0-46.0) |

| | Cameroon 2004 | | Cameroon 2011 | | Cameroon 2018 | |
|---|---|---|---|---|---|---|
| | % | 95%CI | % | 95%CI | % | 95%CI |
| *Five-year age group* | | | | | | |
| 15-19 | 10.3 | (8.9-12.0) | 8.1 | (7.0-9.4) | 7.5 | (6.6-8.6) |
| 20-24 | 20.5 | (18.8-22.4) | 19.7 | (18.2-21.2) | 15.1 | (13.7-16.5) |
| 25-29 | 18.8 | (17.2-20.6) | 21.7 | (20.1-23.4) | 21.9 | (20.5-23.4) |
| 30-34 | 15.7 | (14.1-17.4) | 16 | (14.7-17.5) | 19.8 | (18.4-21.4) |
| 35-39 | 14.4 | (12.8-16.2) | 14 | (12.8-15.3) | 15.0 | (13.8-16.3) |
| 40-44 | 10.7 | (9.2-12.5) | 9.6 | (8.5-10.9) | 10.7 | (9.5-11.9) |
| 45-49 | 9.4 | (8.0-11.1) | 10.9 | (9.6-12.3) | 10.0 | (8.9-11.3) |
| *Place of residence* | | | | | | |
| urban | 49.9 | (47.2-52.6) | 50.4 | (47.9-52.8) | 48.3 | (44.8-51.8) |
| rural | 50.1 | (47.4-52.8) | 49.6 | (47.2-52.1) | 51.7 | (48.2-55.2) |
| *Education* | | | | | | |
| None | 29.2 | (26.7-31.8) | 25.1 | (22.7-27.7) | 27.3 | (24.4-30.4) |
| Primary | 39.5 | (37.2-41.9) | 37.9 | (35.5-40.3) | 32.6 | (30.3-35.0) |
| Secondary+ | 31.2 | (28.9-33.7) | 37 | (34.6-39.5) | 40.1 | (37.5-42.7) |
| *Wealth* | | | | | | |
| poor/poorest | 40.6 | (37.5-43.8) | 37.2 | (34.6-39.8) | 41.4 | (38.3-44.5) |
| middle | 19.4 | (17.4-21.6) | 19.2 | (17.3-21.3) | 20.8 | (19.0-22.8) |
| rich/richest | 40.0 | (37.0-43.0) | 43.6 | (41.0-46.2) | 37.8 | (34.7-41.0) |

***Table S5: Continued…***

| | Colombia 2005 | | Colombia 2010 | | Colombia 2015 | |
|---|---|---|---|---|---|---|
| | % | 95%CI | % | 95%CI | % | 95%CI |
| *Five-year age group* | | | | | | |
| 15-19 | 4.6 | (4.3-5.0) | 4.6 | (4.3-4.9) | 4.0 | (3.7-4.4) |
| 20-24 | 13.1 | (12.5-13.7) | 11.9 | (11.5-12.3) | 12.9 | (12.2-13.6) |
| 25-29 | 15.8 | (15.2-16.5) | 16.2 | (15.7-16.7) | 17.4 | (16.5-18.3) |
| 30-34 | 16.7 | (16.1-17.4) | 17.1 | (16.6-17.6) | 17.4 | (16.5-18.3) |
| 35-39 | 17.7 | (17.0-18.3) | 16.8 | (16.3-17.3) | 16.6 | (15.7-17.5) |
| 40-44 | 16.5 | (15.9-17.2) | 17.4 | (16.9-17.9) | 15.7 | (14.9-16.6) |
| 45-49 | 15.5 | (14.9-16.2) | 16.1 | (15.6-16.6) | 16.0 | (15.2-16.7) |
| *Place of residence* | | | | | | |
| urban | 75.7 | (74.5-76.9) | 76.6 | (75.8-77.3) | 77.8 | (76.6-79.0) |
| rural | 24.3 | (23.1-25.5) | 23.4 | (22.7-24.2) | 22.2 | (21.0-23.4) |
| *Education* | | | | | | |
| None | 3.6 | (3.3-3.9) | 2.3 | (2.1-2.5) | 1.8 | (1.6-2.1) |
| Primary | 33.6 | (32.7-34.6) | 28.9 | (28.2-29.7) | 20.6 | (19.7-21.6) |
| Secondary+ | 62.8 | (61.7-63.8) | 68.8 | (68.0-69.5) | 77.5 | (76.5-78.6) |
| *Wealth* | | | | | | |
| poor/poorest | 37.2 | (35.9-38.5) | 39.3 | (38.3-40.4) | 40.4 | (38.7-42.1) |
| middle | 21.6 | (20.7-22.5) | 21.9 | (21.1-22.8) | 21.7 | (20.6-22.8) |
| rich/richest | 41.2 | (39.9-42.6) | 38.7 | (37.6-39.8) | 37.9 | (35.8-40.1) |

| | Dominican Republic 2002 | | Dominican Republic 2007 | | Dominican Republic 2013 | |
|---|---|---|---|---|---|---|
| | % | 95%CI | % | 95%CI | % | 95%CI |
| *Five-year age group* | | | | | | |
| 15-19 | 7.9 | (7.0-9.0) | 7.7 | (6.7-8.8) | 7.4 | (6.5-8.5) |
| 20-24 | 14.3 | (13.1-15.6) | 13.7 | (12.6-14.9) | 15.6 | (14.3-16.9) |
| 25-29 | 17.6 | (16.4-18.9) | 16.6 | (15.2-18.1) | 15.8 | (14.7-17.1) |
| 30-34 | 17.2 | (16.1-18.4) | 16.5 | (15.4-17.8) | 17.5 | (16.1-18.9) |
| 35-39 | 17.8 | (16.5-19.2) | 18.1 | (16.9-19.4) | 15.7 | (14.3-17.2) |
| 40-44 | 13.1 | (11.9-14.4) | 14.2 | (12.8-15.6) | 14.5 | (13.2-15.9) |
| 45-49 | 12 | (11.0-13.2) | 13.1 | (11.7-14.8) | 13.5 | (12.1-14.9) |
| *Place of residence* | | | | | | |
| urban | 67.2 | (65.7-68.6) | 67.1 | (65.2-68.8) | 74.6 | (71.7-77.3) |
| rural | 32.8 | (31.4-34.3) | 32.9 | (31.2-34.8) | 25.4 | (22.7-28.3) |
| *Education* | | | | | | |
| None | 5.1 | (4.4-5.9) | 5.1 | (4.4-5.9) | 2.4 | (2.0-2.9) |
| Primary | 50.8 | (48.5-53.1) | 44.6 | (42.4-46.8) | 36.0 | (33.2-38.8) |
| Secondary+ | 44.1 | (41.7-46.5) | 50.3 | (48.0-52.6) | 61.6 | (58.8-64.4) |
| *Wealth* | | | | | | |
| poor/poorest | | | 39.6 | (37.3-41.9) | 39.8 | (36.5-43.2) |
| middle | | | 21.4 | (19.8-23.1) | 21.3 | (19.8-23.0) |
| rich/richest | | | 39.0 | (36.2-41.9) | 38.9 | (35.2-42.8) |

***Table S5: Continued…***

| | Haiti 2005-06 | | Haiti 2012 | | Haiti 2016-17 | |
|---|---|---|---|---|---|---|
| | % | 95%CI | % | 95%CI | % | 95%CI |
| *Five-year age group* | | | | | | |
| 15-19 | 6.2 | (5.1-7.6) | 5.1 | (4.4-5.9) | 2.9 | (2.3-3.6) |
| 20-24 | 16.0 | (13.9-18.3) | 15.7 | (14.6-16.8) | 12.6 | (11.3-14.0) |
| 25-29 | 21.5 | (19.3-23.9) | 20.3 | (19.1-21.5) | 18.4 | (16.9-19.9) |
| 30-34 | 16.2 | (14.0-18.6) | 18.5 | (17.3-19.7) | 20.6 | (19.1-22.2) |
| 35-39 | 15.0 | (13.4-16.9) | 15.5 | (14.4-16.6) | 18.9 | (17.4-20.5) |
| 40-44 | 13.3 | (11.3-15.5) | 12.4 | (11.3-13.7) | 13.2 | (11.9-14.5) |
| 45-49 | 11.8 | (10.1-13.7) | 12.6 | (11.5-13.8) | 13.5 | (12.1-15.1) |
| *Place of residence* | | | | | | |
| urban | 45.2 | (41.6-48.8) | 44.7 | (42.0-47.5) | 42.3 | (39.3-45.4) |
| rural | 54.8 | (51.2-58.4) | 55.3 | (52.5-58.0) | 57.7 | (54.6-60.7) |
| *Education* | | | | | | |
| None | 32.1 | (29.1-35.2) | 22.1 | (20.3-24.0) | 20.7 | (18.8-22.8) |
| Primary | 39.1 | (36.1-42.1) | 38.7 | (36.6-40.7) | 36.4 | (34.4-38.6) |
| Secondary+ | 28.8 | (25.5-32.4) | 39.2 | (36.7-41.8) | 42.8 | (40.2-45.6) |
| *Wealth* | | | | | | |
| poor/poorest | 36.0 | (32.0-40.2) | 33.9 | (30.6-37.3) | 35.9 | (32.7-39.2) |
| middle | 17.8 | (15.3-20.6) | 21.4 | (18.8-24.2) | 19.3 | (17.2-21.7) |
| rich/richest | 46.2 | (42.0-50.4) | 44.8 | (41.3-48.2) | 44.8 | (41.2-48.5) |

| | India 2005-06 | | India 2015-16 | | India 2019-21 | |
|---|---|---|---|---|---|---|
| | % | 95%CI | % | 95%CI | % | 95%CI |
| *Five-year age group* | | | | | | |
| 15-19 | 7.0 | (6.6-7.3) | 3.5 | (3.2-3.7) | 2.5 | (2.2-2.8) |
| 20-24 | 17.5 | (17.0-17.9) | 14.9 | (14.4-15.4) | 12.7 | (12.1-13.2) |
| 25-29 | 19.5 | (19.1-19.9) | 19.4 | (19.0-19.9) | 17.2 | (16.7-17.7) |
| 30-34 | 17.6 | (17.2-18.0) | 17.7 | (17.2-18.1) | 17.6 | (17.0-18.1) |
| 35-39 | 15.7 | (15.3-16.1) | 16.6 | (16.2-17.1) | 19.4 | (18.9-20.0) |
| 40-44 | 13.1 | (12.7-13.5) | 14.3 | (13.9-14.8) | 15.4 | (14.8-16.0) |
| 45-49 | 9.7 | (9.4-10.1) | 13.6 | (13.1-14.1) | 15.2 | (14.6-15.7) |
| *Place of residence* | | | | | | |
| urban | 30.7 | (29.6-31.8) | 34.7 | (33.6-35.7) | 30.7 | (29.8-31.7) |
| rural | 69.3 | (68.2-70.4) | 65.3 | (64.3-66.4) | 69.3 | (68.3-70.2) |
| *Education* | | | | | | |
| None | 48.0 | (47.1-49.0) | 32.5 | (31.8-33.2) | 28.5 | (27.8-29.3) |
| Primary | 15.4 | (14.9-15.8) | 14.3 | (13.8-14.8) | 13.9 | (13.4-14.5) |
| Secondary+ | 36.6 | (35.7-37.5) | 53.2 | (52.4-54.0) | 57.6 | (56.7-58.4) |
| *Wealth* | | | | | | |
| poor/poorest | 39.3 | (38.3-40.3) | 36.4 | (35.5-37.3) | 40.6 | (39.8-41.5) |
| middle | 20.1 | (19.5-20.7) | 20.7 | (20.1-21.3) | 21.3 | (20.6-21.9) |
| rich/richest | 40.6 | (39.6-41.6) | 43 | (42.0-44.0) | 38.1 | (37.2-39.0) |

***Table S5: Continued…***

| | Jordan 2007 | | Jordan 2012 | | Jordan 2017-18 | | Jordan 2023 | |
|---|---|---|---|---|---|---|---|---|
| | % | 95%CI | % | 95%CI | % | 95%CI | % | 95%CI |
| *Five-year age group* | | | | | | | | |
| 15-19 | 1.5 | (1.0-2.4) | 2.4 | (1.8-3.1) | 2.8 | (2.2-3.6) | 0.8 | (0.5-1.3) |
| 20-24 | 13.5 | (11.5-15.9) | 10.6 | (9.5-11.8) | 10.4 | (9.3-11.6) | 4.8 | (4.0-5.7) |
| 25-29 | 18.4 | (16.5-20.4) | 17.3 | (15.7-19.0) | 17.5 | (16.1-18.9) | 9.5 | (8.3-10.8) |
| 30-34 | 20.8 | (19.1-22.5) | 19.4 | (17.8-21.1) | 18.3 | (17.1-19.6) | 12.3 | (11.0-13.7) |
| 35-39 | 18.7 | (17.0-20.6) | 18.8 | (17.4-20.2) | 18 | (16.7-19.4) | 16.7 | (14.9-18.6) |
| 40-44 | 16.8 | (15.0-18.8) | 18.4 | (16.8-20.2) | 16.3 | (15.0-17.8) | 22.9 | (20.7-25.1) |
| 45-49 | 10.2 | (8.7-12.0) | 13.1 | (11.9-14.6) | 16.7 | (15.2-18.2) | 33.1 | (30.5-35.9) |
| *Place of residence* | | | | | | | | |
| urban | 85.3 | (84.2-86.3) | 82.7 | (81.7-83.8) | 90.1 | (89.5-90.7) | 91.8 | (90.7-92.8) |
| rural | 14.7 | (13.7-15.8) | 17.3 | (16.2-18.3) | 9.9 | (9.3-10.5) | 8.2 | (7.2-9.3) |
| *Education* | | | | | | | | |
| None | 3.8 | (2.9-4.9) | 2.2 | (1.7-2.8) | 2.1 | (1.7-2.7) | 2.1 | (1.1-3.9) |
| Primary | 6.3 | (5.2-7.6) | 7.6 | (6.6-8.8) | 8.7 | (7.5-9.9) | 4.6 | (3.8-5.6) |
| Secondary+ | 89.9 | (88.2-91.5) | 90.2 | (88.8-91.4) | 89.2 | (87.9-90.5) | 93.3 | (91.2-94.9) |
| *Wealth* | | | | | | | | |
| poor/poorest | 40.8 | (36.5-45.4) | 40.4 | (37.6-43.3) | 40.3 | (37.7-42.9) | 35.6 | (32.4-39.0) |
| middle | 20.5 | (17.8-23.5) | 22.1 | (20.3-24.0) | 20.9 | (19.3-22.5) | 20.9 | (18.9-23.1) |
| rich/richest | 38.6 | (33.6-44.0) | 37.5 | (34.4-40.7) | 38.9 | (36.0-41.8) | 43.5 | (39.4-47.6) |

| | Kenya 2008-09 | | Kenya 2014 | | Kenya 2022 | |
|---|---|---|---|---|---|---|
| | % | 95%CI | % | 95%CI | % | 95%CI |
| *Five-year age group* | | | | | | |
| 15-19 | 3.9 | (3.1-4.9) | 2.8 | (2.2-3.4) | 2.3 | (1.9-2.6) |
| 20-24 | 18.7 | (17.1-20.5) | 16.1 | (14.7-17.6) | 13.9 | (13.0-14.9) |
| 25-29 | 21.1 | (19.5-22.9) | 24.9 | (23.0-26.9) | 22.6 | (21.4-23.7) |
| 30-34 | 19.1 | (17.5-20.7) | 19.0 | (17.5-20.5) | 19.1 | (18.2-20.1) |
| 35-39 | 14.4 | (13.0-15.8) | 15.7 | (14.4-17.2) | 18.0 | (17.0-19.0) |
| 40-44 | 12.0 | (10.7-13.4) | 12.4 | (11.1-13.8) | 13.3 | (12.3-14.4) |
| 45-49 | 10.8 | (9.4-12.4) | 9.2 | (8.1-10.4) | 10.8 | (10.0-11.8) |
| *Place of residence* | | | | | | |
| urban | 23.6 | (18.7-29.2) | 39.5 | (37.5-41.5) | 39.2 | (37.6-40.9) |
| rural | 76.4 | (70.8-81.3) | 60.5 | (58.5-62.5) | 60.8 | (59.1-62.4) |
| *Education* | | | | | | |
| None | 11.8 | (9.6-14.6) | 9.3 | (8.3-10.5) | 7.3 | (6.7-7.9) |
| Primary | 59.8 | (56.7-62.8) | 56.4 | (54.3-58.5) | 42.9 | (41.3-44.5) |
| Secondary+ | 28.4 | (25.4-31.6) | 34.3 | (32.2-36.4) | 49.8 | (48.2-51.5) |
| *Wealth* | | | | | | |
| poor/poorest | 36.4 | (32.4-40.6) | 37 | (35.0-39.0) | 34.8 | (33.3-36.5) |
| middle | 19.5 | (17.2-22.0) | 18.9 | (17.4-20.5) | 19.1 | (18.1-20.3) |
| rich/richest | 44.1 | (39.0-49.4) | 44.1 | (42.0-46.3) | 46.0 | (44.2-47.9) |

***Table S5: Continued…***

| | Malawi 2004 | | Malawi 2010 | | Malawi 2015-16 | |
|---|---|---|---|---|---|---|
| | % | 95%CI | % | 95%CI | % | 95%CI |
| *Five-year age group* | | | | | | |
| 15-19 | 8.8 | (8.0-9.6) | 6.9 | (6.1-7.9) | 7.2 | (6.2-8.2) |
| 20-24 | 25.8 | (24.6-27.0) | 20.7 | (19.3-22.1) | 21 | (19.5-22.6) |
| 25-29 | 21.8 | (20.7-23.0) | 24.3 | (22.9-25.8) | 20.2 | (18.8-21.6) |
| 30-34 | 15.0 | (14.0-15.9) | 16.0 | (14.8-17.2) | 19.6 | (17.9-21.5) |
| 35-39 | 11.2 | (10.4-12.1) | 14.4 | (13.1-15.8) | 14.0 | (12.8-15.3) |
| 40-44 | 9.6 | (8.8-10.5) | 9.3 | (8.3-10.4) | 10.7 | (9.6-11.9) |
| 45-49 | 7.8 | (7.1-8.7) | 8.4 | (7.4-9.6) | 7.3 | (6.4-8.4) |
| *Place of residence* | | | | | | |
| urban | 15.9 | (13.4-18.7) | 18.0 | (16.2-19.9) | 16.7 | (15.4-18.1) |
| rural | 84.1 | (81.3-86.6) | 82.0 | (80.1-83.8) | 83.3 | (81.9-84.6) |
| *Education* | | | | | | |
| None | 26.6 | (25.0-28.3) | 18.9 | (17.4-20.4) | 14.4 | (13.1-15.7) |
| Primary | 61.9 | (60.3-63.5) | 65.4 | (63.5-67.3) | 63.8 | (61.7-65.8) |
| Secondary+ | 11.5 | (10.1-13.1) | 15.7 | (14.1-17.4) | 21.8 | (20.0-23.8) |
| *Wealth* | | | | | | |
| poor/poorest | 39.4 | (37.2-41.6) | 38.7 | (36.7-40.9) | 41.4 | (39.4-43.4) |
| middle | 21.2 | (19.9-22.6) | 20.2 | (18.8-21.6) | 19.5 | (18.0-21.0) |
| rich/richest | 39.4 | (37.0-42.0) | 41.1 | (38.7-43.5) | 39.2 | (36.9-41.5) |

| | Mali 2006 | | Mali 2012-13 | | Mali 2018 | |
|---|---|---|---|---|---|---|
| | % | 95%CI | % | 95%CI | % | 95%CI |
| *Five-year age group* | | | | | | |
| 15-19 | 12.5 | (11.6-13.5) | 9 | (7.7-10.4) | 10.3 | (9.0-11.9) |
| 20-24 | 19.5 | (18.3-20.8) | 18.4 | (16.7-20.2) | 16.7 | (15.2-18.4) |
| 25-29 | 20.4 | (19.3-21.7) | 22.1 | (20.3-23.9) | 21.5 | (19.6-23.4) |
| 30-34 | 14.9 | (13.9-16.0) | 18.0 | (16.5-19.7) | 19.0 | (17.4-20.7) |
| 35-39 | 13.6 | (12.6-14.7) | 13.6 | (12.1-15.3) | 14.9 | (13.3-16.6) |
| 40-44 | 10.0 | (9.0-11.2) | 10.7 | (9.4-12.2) | 10.3 | (9.0-11.9) |
| 45-49 | 9.0 | (7.9-10.3) | 8.2 | (6.9-9.8) | 7.3 | (6.2-8.5) |
| *Place of residence* | | | | | | |
| urban | 31.2 | (26.4-36.4) | 21.0 | (19.0-23.1) | 22.5 | (20.2-24.9) |
| rural | 68.8 | (63.6-73.6) | 79.0 | (76.9-81.0) | 77.5 | (75.1-79.8) |
| *Education* | | | | | | |
| None | 81.5 | (79.5-83.4) | 80.1 | (77.7-82.4) | 71.9 | (69.4-74.4) |
| Primary | 10.9 | (9.5-12.4) | 10.2 | (8.8-11.8) | 13.1 | (11.6-14.8) |
| Secondary+ | 7.6 | (6.6-8.7) | 9.7 | (8.0-11.6) | 14.9 | (13.0-17.1) |
| *Wealth* | | | | | | |
| poor/poorest | 38.7 | (35.1-42.4) | 40.4 | (36.8-44.1) | 39.0 | (35.6-42.5) |
| middle | 19.7 | (18.0-21.6) | 19.6 | (17.8-21.5) | 20.6 | (18.7-22.7) |
| rich/richest | 41.6 | (37.3-46.0) | 40.1 | (36.4-43.9) | 40.4 | (36.8-44.1) |

***Table S5: Continued…***

| | Mozambique 2011 | | Mozambique 2015 | | Mozambique 2022-23 | |
|---|---|---|---|---|---|---|
| | % | 95%CI | % | 95%CI | % | 95%CI |
| *Five-year age group* | | | | | | |
| 15-19 | 10.9 | (9.9-12.0) | 8.6 | (7.3-10.1) | 9.2 | (8.0-10.5) |
| 20-24 | 19.2 | (17.9-20.6) | 20.6 | (18.7-22.6) | 20.5 | (18.9-22.3) |
| 25-29 | 18.5 | (17.2-19.8) | 19.0 | (17.2-20.9) | 20.0 | (18.4-21.7) |
| 30-34 | 17.1 | (15.9-18.4) | 15.4 | (13.7-17.1) | 13.9 | (12.6-15.4) |
| 35-39 | 15.0 | (13.8-16.1) | 15.1 | (13.5-16.8) | 14.6 | (13.2-16.1) |
| 40-44 | 9.7 | (8.8-10.7) | 11.8 | (10.5-13.3) | 11.3 | (10.0-12.7) |
| 45-49 | 9.7 | (8.7-10.7) | 9.6 | (8.2-11.1) | 10.5 | (9.3-11.8) |
| *Place of residence* | | | | | | |
| urban | 30.7 | (28.5-32.9) | 30.7 | (27.7-33.9) | 33.8 | (31.5-36.1) |
| rural | 69.3 | (67.1-71.5) | 69.3 | (66.1-72.3) | 66.2 | (63.9-68.5) |
| *Education* | | | | | | |
| None | 36.2 | (33.9-38.6) | 32.4 | (29.6-35.4) | 32.2 | (29.7-34.8) |
| Primary | 50.8 | (48.6-53.0) | 51.5 | (48.6-54.3) | 45.0 | (42.5-47.5) |
| Secondary+ | 13.0 | (11.4-14.7) | 16.1 | (14.1-18.4) | 22.8 | (20.8-25.0) |
| *Wealth* | | | | | | |
| poor/poorest | 40.3 | (37.7-42.9) | 41.8 | (38.3-45.5) | 40.5 | (37.6-43.4) |
| middle | 21.2 | (19.4-23.1) | 21.1 | (18.6-23.8) | 20.3 | (18.4-22.4) |
| rich/richest | 38.5 | (35.9-41.2) | 37.1 | (33.8-40.5) | 39.2 | (36.3-42.2) |

| | Nepal 2011 | | Nepal 2016 | | Nepal 2022 | |
|---|---|---|---|---|---|---|
| | % | 95%CI | % | 95%CI | % | 95%CI |
| *Five-year age group* | | | | | | |
| 15-19 | 8.1 | (6.6-9.8) | 6.5 | (5.6-7.6) | 5 | (4.1-6.0) |
| 20-24 | 19.6 | (17.7-21.7) | 16.9 | (15.4-18.4) | 15.7 | (14.4-17.1) |
| 25-29 | 18.7 | (17.0-20.5) | 18.8 | (17.3-20.5) | 19.4 | (18.1-20.9) |
| 30-34 | 15.7 | (14.4-17.2) | 18.2 | (16.6-19.8) | 17.5 | (16.1-19.0) |
| 35-39 | 14.4 | (12.9-16.2) | 16.6 | (15.1-18.3) | 16.6 | (15.3-17.9) |
| 40-44 | 12.2 | (10.5-14.2) | 13.5 | (12.2-15.0) | 14.1 | (12.7-15.6) |
| 45-49 | 11.1 | (9.7-12.7) | 9.5 | (8.3-10.9) | 11.7 | (10.5-13.1) |
| *Place of residence* | | | | | | |
| urban | 24.1 | (21.9-26.5) | 59.9 | (55.1-64.5) | 66.9 | (65.3-68.6) |
| rural | 75.9 | (73.5-78.1) | 40.1 | (35.5-44.9) | 33.1 | (31.4-34.7) |
| *Education* | | | | | | |
| None | 48.8 | (45.4-52.1) | 41.9 | (39.2-44.5) | 32.5 | (30.3-34.7) |
| Primary | 18.2 | (16.3-20.4) | 18.7 | (16.7-20.9) | 33.4 | (31.5-35.3) |
| Secondary+ | 33.0 | (30.0-36.2) | 39.4 | (36.6-42.3) | 34.2 | (31.9-36.5) |
| *Wealth* | | | | | | |
| poor/poorest | 38.0 | (33.7-42.4) | 37.6 | (34.4-41.0) | 38.5 | (35.5-41.6) |
| middle | 19.4 | (16.6-22.4) | 21.2 | (19.4-23.2) | 20.9 | (19.1-22.8) |
| rich/richest | 42.7 | (38.0-47.5) | 41.1 | (37.7-44.6) | 40.6 | (37.4-43.9) |

***Table S5: Continued…***

| | Nigeria 2008 | | Nigeria 2013 | | Nigeria 2018 | |
|---|---|---|---|---|---|---|
| | % | 95%CI | % | 95%CI | % | 95%CI |
| *Five-year age group* | | | | | | |
| 15-19 | 7.9 | (7.4-8.5) | 7.7 | (7.1-8.4) | 5.4 | (4.9-6.0) |
| 20-24 | 15.5 | (14.9-16.2) | 15.5 | (14.8-16.1) | 13.6 | (12.7-14.6) |
| 25-29 | 21.5 | (20.8-22.2) | 20.4 | (19.7-21.1) | 20.6 | (19.5-21.7) |
| 30-34 | 17.6 | (17.0-18.3) | 17.1 | (16.5-17.8) | 19.2 | (18.2-20.3) |
| 35-39 | 15.0 | (14.4-15.7) | 15.3 | (14.7-16.0) | 18.4 | (17.4-19.5) |
| 40-44 | 11.6 | (11.0-12.2) | 12.3 | (11.7-12.9) | 11.7 | (10.8-12.7) |
| 45-49 | 10.8 | (10.2-11.5) | 11.7 | (11.0-12.3) | 11.0 | (10.1-12.0) |
| *Place of residence* | | | | | | |
| urban | 31.6 | (30.3-32.9) | 37.2 | (35.4-39.1) | 43.3 | (41.5-45.3) |
| rural | 68.4 | (67.1-69.7) | 62.8 | (60.9-64.6) | 56.7 | (54.7-58.5) |
| *Education* | | | | | | |
| None | 46.7 | (44.9-48.6) | 47.1 | (44.9-49.3) | 41.1 | (39.2-43.1) |
| Primary | 22.5 | (21.4-23.7) | 19.7 | (18.6-20.9) | 16.6 | (15.5-17.7) |
| Secondary+ | 30.8 | (29.2-32.4) | 33.2 | (31.4-35.1) | 42.3 | (40.5-44.1) |
| *Wealth* | | | | | | |
| poor/poorest | 44.3 | (42.2-46.4) | 43.4 | (41.0-45.7) | 38.7 | (36.5-40.9) |
| middle | 18.6 | (17.2-20.0) | 18.5 | (17.2-19.9) | 20.6 | (19.2-22.2) |
| rich/richest | 37.1 | (35.1-39.2) | 38.1 | (36.0-40.2) | 40.7 | (38.6-42.8) |

***Table S5: Continued…***

| | Peru 2004-06 | | Peru 2007-08 | | Peru 2009 | | Peru 2010 | | Peru 2011 | | Peru 2012 | |
|---|---|---|---|---|---|---|---|---|---|---|---|---|
| | % | 95%CI | % | 95%CI | % | 95%CI | % | CIs | % | CIs | % | CIs |
| *Five-year age group* | | | | | | | | | | | | |
| 15-19 | 2.8 | (2.4-3.3) | 3.0 | (2.7-3.5) | 3.1 | (2.8-3.5) | 2.9 | (2.6-3.3) | 2.9 | (2.6-3.4) | 3.2 | (2.9-3.6) |
| 20-24 | 11.0 | (10.2-11.9) | 10.7 | (9.9-11.5) | 10.8 | (10.1-11.5) | 11.1 | (10.3-11.9) | 10.6 | (9.9-11.3) | 10.9 | (10.1-11.7) |
| 25-29 | 16.7 | (15.7-17.8) | 17.9 | (17.0-18.8) | 17.7 | (16.8-18.6) | 17.6 | (16.8-18.5) | 17.6 | (16.6-18.7) | 16.3 | (15.5-17.1) |
| 30-34 | 20.9 | (19.9-22.0) | 20.4 | (19.5-21.4) | 20 | (19.1-20.9) | 20.7 | (19.8-21.7) | 19.9 | (19.0-20.8) | 20.3 | (19.3-21.2) |
| 35-39 | 19.5 | (18.5-20.6) | 17.8 | (16.9-18.8) | 19.5 | (18.5-20.5) | 18.9 | (18.0-19.8) | 19.3 | (18.4-20.3) | 18.9 | (18.0-19.8) |
| 40-44 | 15.7 | (14.8-16.7) | 16.2 | (15.2-17.2) | 15.8 | (15.0-16.7) | 15.4 | (14.5-16.3) | 16.3 | (15.4-17.2) | 16.6 | (15.8-17.5) |
| 45-49 | 13.2 | (12.4-14.1) | 14.1 | (13.1-15.0) | 13.2 | (12.3-14.0) | 13.4 | (12.6-14.3) | 13.4 | (12.6-14.3) | 13.8 | (13.1-14.7) |
| *Place of residence* | | | | | | | | | | | | |
| urban | 62.2 | (60.5-63.9) | 67.0 | (65.3-68.6) | 68.8 | (67.6-70.0) | 69.3 | (68.1-70.5) | 69.8 | (68.5-71.1) | 70.0 | (68.7-71.3) |
| rural | 37.8 | (36.1-39.5) | 33.0 | (31.4-34.7) | 31.2 | (30.0-32.4) | 30.7 | (29.5-31.9) | 30.2 | (28.9-31.5) | 30.0 | (28.7-31.3) |
| *Education* | | | | | | | | | | | | |
| None | 5.1 | (4.3-5.9) | 4.3 | (3.6-5.0) | 3.9 | (3.4-4.4) | 3.1 | (2.7-3.6) | 3.6 | (3.1-4.1) | 3.3 | (2.9-3.8) |
| Primary | 34.0 | (32.3-35.6) | 33.5 | (32.0-35.0) | 31.0 | (29.8-32.2) | 30.8 | (29.6-32.1) | 29.3 | (28.1-30.6) | 29.9 | (28.5-31.3) |
| Secondary+ | 61.0 | (59.0-62.8) | 62.3 | (60.6-64.0) | 65.2 | (63.8-66.5) | 66.0 | (64.6-67.4) | 67.1 | (65.7-68.5) | 66.8 | (65.3-68.3) |
| *Wealth* | | | | | | | | | | | | |
| poor/poorest | 38.1 | (36.0-40.3) | 29.6 | (27.8-31.4) | 40.5 | (38.9-42.1) | 40.0 | (38.4-41.7) | 40.2 | (38.6-41.8) | 40.4 | (38.8-42.0) |
| middle | 21.8 | (20.0-23.7) | 23.3 | (21.6-25.2) | 22.6 | (21.0-24.2) | 22.4 | (21.0-24.0) | 22.8 | (21.3-24.4) | 22.4 | (21.0-23.8) |
| rich/richest | 40.1 | (37.9-42.3) | 47.1 | (44.9-49.2) | 37.0 | (35.0-38.9) | 37.6 | (35.6-39.6) | 37.0 | (35.0-39.1) | 37.2 | (35.4-39.1) |

***Table S5: Continued…***

| | Philippines 2008 | | Philippines 2013 | | Philippines 2017 | | Philippines 2022 | |
|---|---|---|---|---|---|---|---|---|
| | % | 95% CI | % | 95% CI | % | 95% CI | % | CIs |
| *Five-year age group* | | | | | | | | |
| 15-19 | 2.9 | (2.6-3.4) | 3.1 | (2.6-3.6) | 2.8 | (2.3-3.6) | 2.0 | (1.6-2.4) |
| 20-24 | 11.2 | (10.4-12.0) | 12.2 | (11.4-13.1) | 10.9 | (10.1-11.7) | 8.2 | (7.4-9.0) |
| 25-29 | 19.1 | (18.0-20.1) | 15.5 | (14.6-16.3) | 17.3 | (16.1-18.5) | 15.0 | (14.0-16.1) |
| 30-34 | 20.4 | (19.4-21.4) | 19.1 | (18.2-20.1) | 18.1 | (16.8-19.4) | 19.0 | (17.9-20.0) |
| 35-39 | 19.1 | (18.2-20.1) | 17.2 | (16.3-18.1) | 18.8 | (17.8-19.8) | 18.7 | (17.7-19.7) |
| 40-44 | 14.7 | (13.8-15.6) | 17.3 | (16.4-18.3) | 15.9 | (14.9-17.0) | 19.5 | (18.3-20.8) |
| 45-49 | 12.7 | (11.9-13.5) | 15.6 | (14.6-16.7) | 16.3 | (15.2-17.4) | 17.7 | (16.5-18.9) |
| *Place of residence* | | | | | | | | |
| urban | 53.3 | (49.8-56.7) | 49.9 | (48.7-51.1) | 45.5 | (41.9-49.2) | 54.5 | (50.8-58.2) |
| rural | 46.7 | (43.3-50.2) | 50.1 | (48.9-51.3) | 54.5 | (50.8-58.1) | 45.5 | (41.8-49.2) |
| *Education* | | | | | | | | |
| None | 1.5 | (1.2-1.9) | 1.6 | (1.2-2.1) | 0.9 | (0.7-1.2) | 0.9 | (0.6-1.2) |
| Primary | 23.3 | (22.0-24.7) | 20.7 | (19.4-22.0) | 17.6 | (16.3-19.0) | 13.6 | (12.6-14.7) |
| Secondary+ | 75.2 | (73.7-76.6) | 77.8 | (76.4-79.1) | 81.5 | (80.0-82.9) | 85.5 | (84.4-86.6) |
| *Wealth* | | | | | | | | |
| poor/poorest | 40.3 | (38.4-42.2) | 38.7 | (37.0-40.3) | 39.8 | (37.6-42.0) | 39.8 | (37.8-41.9) |
| middle | 20.6 | (19.3-21.9) | 20.2 | (19.1-21.4) | 20.1 | (18.6-21.7) | 21.1 | (19.7-22.6) |
| rich/richest | 39.1 | (37.3-41.1) | 41.1 | (39.4-42.9) | 40.0 | (37.1-43.1) | 39.1 | (36.9-41.3) |

***Table S5: Continued…***

| | Rwanda 2010 | | Rwanda 2014-15 | | Rwanda 2019-20 | |
|---|---|---|---|---|---|---|
| | % | 95% CI | % | 95% CI | % | 95% CI |
| *Five-year age group* | | | | | | |
| 15-19 | 1.4 | (1.1-1.9) | 1.1 | (0.7-1.8) | 1.3 | (0.8-2.1) |
| 20-24 | 13.1 | (11.9-14.4) | 13.2 | (11.5-15.1) | 9.7 | (8.4-11.2) |
| 25-29 | 23.1 | (21.8-24.6) | 19.2 | (17.4-21.2) | 15.4 | (13.8-17.1) |
| 30-34 | 18.8 | (17.5-20.2) | 24.3 | (22.1-26.5) | 22.9 | (20.9-25.1) |
| 35-39 | 16.2 | (14.9-17.6) | 15.8 | (14.1-17.6) | 21.4 | (19.3-23.7) |
| 40-44 | 14.1 | (12.7-15.7) | 14.9 | (13.0-16.9) | 17.3 | (15.3-19.6) |
| 45-49 | 13.1 | (11.8-14.6) | 11.5 | (9.7-13.6) | 11.9 | (10.3-13.8) |
| *Place of residence* | | | | | | |
| urban | 13.4 | (11.1-16.1) | 17.5 | (15.7-19.4) | 16.5 | (14.7-18.4) |
| rural | 86.6 | (83.9-88.9) | 82.5 | (80.6-84.3) | 83.5 | (81.6-85.3) |
| *Education* | | | | | | |
| None | 21.6 | (19.9-23.4) | 18.1 | (16.1-20.2) | 14.0 | (12.0-16.2) |
| Primary | 68.6 | (66.7-70.5) | 70.2 | (67.7-72.6) | 63.6 | (61.1-66.1) |
| Secondary+ | 9.8 | (8.5-11.3) | 11.7 | (10.1-13.6) | 22.4 | (20.2-24.8) |
| *Wealth* | | | | | | |
| poor/poorest | 41.9 | (39.9-44.0) | 41.9 | (39.2-44.7) | 40.1 | (37.1-43.0) |
| middle | 20.1 | (18.6-21.7) | 19.7 | (17.7-21.9) | 19.1 | (17.1-21.3) |
| rich/richest | 38.0 | (35.8-40.2) | 38.4 | (35.6-41.3) | 40.8 | (37.5-44.2) |

| | Senegal 2017 | | Senegal 2018 | | Senegal 2019 | |
|---|---|---|---|---|---|---|
| | % | 95% CI | % | 95% CI | % | 95% CI |
| *Five-year age group* | | | | | | |
| 15-19 | 7.8 | (6.5-9.4) | 6.7 | (4.9-9.0) | 7.4 | (5.7-9.5) |
| 20-24 | 15.1 | (13.4-16.9) | 15.2 | (12.2-18.9) | 15.7 | (13.1-18.8) |
| 25-29 | 21.1 | (19.0-23.3) | 20.3 | (17.1-24.0) | 17.1 | (14.5-20.1) |
| 30-34 | 19.4 | (17.6-21.4) | 17.7 | (15.0-20.9) | 20.1 | (16.7-23.9) |
| 35-39 | 13.7 | (12.0-15.7) | 16.4 | (13.5-19.7) | 18.6 | (15.9-21.8) |
| 40-44 | 13.7 | (11.9-15.6) | 13.5 | (11.1-16.4) | 10.4 | (8.3-12.9) |
| 45-49 | 9.2 | (7.5-11.1) | 10.1 | (7.6-13.3) | 10.7 | (8.4-13.5) |
| *Place of residence* | | | | | | |
| urban | 40.6 | (38.0-43.4) | 42.8 | (38.9-46.8) | 43.0 | (38.6-47.5) |
| rural | 59.4 | (56.6-62.0) | 57.2 | (53.2-61.1) | 57.0 | (52.5-61.4) |
| *Education* | | | | | | |
| None | 61.2 | (58.4-64.0) | 56.0 | (51.6-60.4) | 59.5 | (55.1-63.7) |
| Primary | 22.9 | (20.7-25.4) | 23.2 | (19.8-26.9) | 23.7 | (20.4-27.5) |
| Secondary+ | 15.8 | (13.9-17.9) | 20.8 | (17.2-24.9) | 16.8 | (14.0-20.0) |
| *Wealth* | | | | | | |
| poor/poorest | 42.2 | (38.5-45.9) | 39.4 | (33.8-45.3) | 38.3 | (33.3-43.5) |
| middle | 20.9 | (18.2-23.8) | 18.5 | (15.0-22.4) | 19.0 | (15.7-22.7) |
| rich/richest | 36.9 | (33.6-40.4) | 42.2 | (37.3-47.1) | 42.8 | (38.0-47.7) |

***Table S5: Continued…***

| | Tanzania 2010 | | Tanzania 2015-16 | | Tanzania 2022 | |
|---|---|---|---|---|---|---|
| | % | 95% CI | % | 95% CI | % | 95% CI |
| *Five-year age group* | | | | | | |
| 15-19 | 5.4 | (4.5-6.5) | 6.6 | (5.9-7.5) | 5.6 | (4.8-6.7) |
| 20-24 | 18.4 | (17.1-19.9) | 18.0 | (16.8-19.2) | 16.9 | (15.4-18.4) |
| 25-29 | 20.5 | (19.2-21.8) | 19.0 | (17.9-20.2) | 19.4 | (17.9-21.0) |
| 30-34 | 17.7 | (16.4-19.0) | 16.2 | (15.0-17.4) | 16.9 | (15.5-18.4) |
| 35-39 | 16.4 | (15.1-17.7) | 16.5 | (15.4-17.7) | 15.4 | (14.0-17.0) |
| 40-44 | 12 | (11.0-13.1) | 13.7 | (12.6-15.0) | 13.9 | (12.2-15.7) |
| 45-49 | 9.6 | (8.6-10.8) | 9.9 | (9.0-11.0) | 11.9 | (10.6-13.4) |
| *Place of residence* | | | | | | |
| urban | 25.4 | (23.1-28.0) | 32.4 | (30.2-34.6) | 32.4 | (27.9-37.2) |
| rural | 74.6 | (72.0-76.9) | 67.6 | (65.4-69.8) | 67.6 | (62.8-72.1) |
| *Education* | | | | | | |
| None | 23.0 | (20.9-25.2) | 17.8 | (16.3-19.4) | 19.9 | (17.6-22.4) |
| Primary | 69.9 | (67.7-71.9) | 66.8 | (64.7-68.8) | 59.8 | (57.5-61.9) |
| Secondary+ | 7.2 | (6.1-8.4) | 15.4 | (13.6-17.5) | 20.4 | (18.6-22.3) |
| *Wealth* | | | | | | |
| poor/poorest | 39.5 | (36.9-42.1) | 38.2 | (35.4-41.0) | 37.1 | (33.6-40.8) |
| middle | 20.6 | (19.0-22.4) | 18.9 | (17.4-20.4) | 19.8 | (18.0-21.8) |
| rich/richest | 39.9 | (37.1-42.9) | 42.9 | (40.3-45.6) | 43.1 | (39.1-47.2) |

| | Uganda 2006 | | Uganda 2011 | | Uganda 2016 | |
|---|---|---|---|---|---|---|
| | % | 95% CI | % | 95% CI | % | 95% CI |
| *Five-year age group* | | | | | | |
| 15-19 | 6.3 | (4.9-7.9) | 7.7 | (6.1-9.7) | 7.3 | (6.6-8.2) |
| 20-24 | 20.5 | (18.2-23.0) | 19.8 | (17.4-22.4) | 21.0 | (19.8-22.3) |
| 25-29 | 19.6 | (17.5-21.8) | 23.0 | (20.3-25.9) | 19.2 | (18.2-20.2) |
| 30-34 | 18.6 | (16.5-21.0) | 15.6 | (13.6-17.9) | 17.8 | (16.8-18.9) |
| 35-39 | 15.0 | (13.0-17.2) | 14.4 | (12.4-16.7) | 14.6 | (13.6-15.6) |
| 40-44 | 11.0 | (9.1-13.1) | 10.2 | (8.5-12.2) | 10.9 | (10.0-11.8) |
| 45-49 | 9.1 | (7.4-11.1) | 9.3 | (7.6-11.4) | 9.2 | (8.2-10.2) |
| *Place of residence* | | | | | | |
| urban | 14.2 | (11.9-16.8) | 17.1 | (14.6-19.9) | 23.5 | (21.8-25.3) |
| rural | 85.8 | (83.2-88.1) | 82.9 | (80.1-85.4) | 76.5 | (74.7-78.2) |
| *Education* | | | | | | |
| None | 23.3 | (20.8-26.1) | 16.9 | (14.3-19.7) | 12.9 | (11.9-14.1) |
| Primary | 62.8 | (60.1-65.5) | 60.8 | (56.8-64.6) | 60.0 | (58.2-61.7) |
| Secondary+ | 13.9 | (11.9-16.0) | 22.4 | (19.6-25.4) | 27.1 | (25.4-28.8) |
| *Wealth* | | | | | | |
| poor/poorest | 38.6 | (35.2-42.0) | 38.6 | (35.4-41.8) | 39.7 | (37.8-41.7) |
| middle | 20.9 | (18.1-24.0) | 19.6 | (17.1-22.3) | 19.6 | (18.3-21.0) |
| rich/richest | 40.5 | (37.1-44.0) | 41.9 | (38.4-45.5) | 40.6 | (38.3-43.1) |

***Table S5: Continued…***

| | Zambia 2007 | | Zambia 2013-14 | | Zambia 2018 | |
|---|---|---|---|---|---|---|
| | % | 95% CI | % | 95% CI | % | 95% CI |
| *Five-year age group* | | | | | | |
| 15-19 | 5.8 | (5.0-6.7) | 5.5 | (4.8-6.2) | 4.8 | (4.2-5.5) |
| 20-24 | 19.5 | (18.0-21.1) | 16.7 | (15.8-17.8) | 17.6 | (16.4-18.8) |
| 25-29 | 23.0 | (21.6-24.6) | 21.0 | (20.0-22.1) | 19.8 | (18.7-21.0) |
| 30-34 | 18.7 | (17.4-20.2) | 19.9 | (18.9-20.9) | 18.3 | (17.0-19.8) |
| 35-39 | 13.5 | (12.2-14.9) | 16.6 | (15.4-17.8) | 17.3 | (16.0-18.7) |
| 40-44 | 10.3 | (9.1-11.7) | 11.6 | (10.7-12.6) | 12.7 | (11.6-13.8) |
| 45-49 | 9.1 | (7.9-10.4) | 8.6 | (7.9-9.5) | 9.5 | (8.5-10.6) |
| *Place of residence* | | | | | | |
| urban | 37.1 | (34.4-39.8) | 42.0 | (40.2-43.8) | 41.5 | (38.3-44.8) |
| rural | 62.9 | (60.2-65.6) | 58.0 | (56.2-59.8) | 58.5 | (55.2-61.7) |
| *Education* | | | | | | |
| None | 12.7 | (11.2-14.4) | 10.9 | (9.9-12.0) | 10.2 | (9.0-11.6) |
| Primary | 60.0 | (57.5-62.4) | 55.0 | (53.3-56.6) | 50.5 | (48.6-52.5) |
| Secondary+ | 27.3 | (25.1-29.7) | 34.1 | (32.4-35.9) | 39.3 | (37.1-41.4) |
| *Wealth* | | | | | | |
| poor/poorest | 39.1 | (36.3-41.9) | 39.6 | (38.0-41.3) | 40.0 | (37.4-42.7) |
| middle | 19.6 | (17.6-21.6) | 19.5 | (18.2-20.9) | 19.2 | (17.6-20.9) |
| rich/richest | 41.4 | (38.5-44.3) | 40.9 | (38.9-42.9) | 40.7 | (38.0-43.5) |

| | Zimbabwe 2005-05 | | Zimbabwe 2010-11 | | Zimbabwe 2015 | |
|---|---|---|---|---|---|---|
| | % | 95%CI | % | 95%CI | % | 95%CI |
| *Five-year age group* | | | | | | |
| 15-19 | 6.8 | (6.0-7.7) | 7.1 | (6.2-8.2) | 6.4 | (5.5-7.4) |
| 20-24 | 22.6 | (21.2-24.1) | 20.1 | (18.9-21.5) | 16.2 | (15.0-17.5) |
| 25-29 | 19.9 | (18.8-21.2) | 22.1 | (20.8-23.5) | 19.7 | (18.5-20.9) |
| 30-34 | 17.6 | (16.3-19.0) | 17.6 | (16.4-18.8) | 20.9 | (19.6-22.3) |
| 35-39 | 13.2 | (12.1-14.4) | 14.8 | (13.7-16.0) | 16.1 | (14.9-17.3) |
| 40-44 | 11 | (9.8-12.4) | 10.2 | (9.2-11.2) | 12.9 | (11.8-14.0) |
| 45-49 | 8.8 | (7.8-9.9) | 8.0 | (7.1-9.0) | 7.9 | (7.0-8.8) |
| *Place of residence* | | | | | | |
| urban | 35.2 | (32.9-37.5) | 33.8 | (31.6-36.1) | 35.6 | (33.3-38.0) |
| rural | 64.8 | (62.5-67.1) | 66.2 | (63.9-68.4) | 64.4 | (62.0-66.7) |
| *Education* | | | | | | |
| None | 5.2 | (4.2-6.4) | 3.0 | (2.4-3.7) | 1.6 | (1.2-2.3) |
| Primary | 36.3 | (34.4-38.3) | 32.5 | (30.6-34.4) | 29.9 | (27.9-32.0) |
| Secondary+ | 58.4 | (56.0-60.9) | 64.5 | (62.4-66.6) | 68.4 | (66.2-70.6) |
| *Wealth* | | | | | | |
| poor/poorest | 36.3 | (33.5-39.2) | 38.0 | (35.4-40.6) | 37.2 | (34.4-40.0) |
| middle | 17.8 | (16.1-19.6) | 19.6 | (17.4-21.9) | 17.5 | (15.9-19.3) |
| rich/richest | 45.9 | (43.1-48.8) | 42.5 | (39.5-45.5) | 45.3 | (42.7-47.8) |

**Table S6: Prevalence of recent intimate parter violence, using country specific harmonised measures**

| Survey | 12-month physical IPV % | 12-month sexual IPV % | 12-month physical and/or sexual IPV % |
|---|---|---|---|
| Cambodia 2005 | 8.3 (6.9-10.0) | 1.7 (1.2-2.4) | 9.0 (7.6-10.8) |
| Cambodia 2014 | 9.3 (8.1-10.8) | 3.9 (3.0-5.2) | 10.9 (9.5-12.6) |
| Cambodia 2021-22 | 4.6 (4.0-5.4) | 1.9 (1.5-2.5) | 5.6 (4.8-6.4) |
| Cameroon 2004 | 22.8 (20.8-24.9) | 14.0 (12.4-15.8) | 30.5 (28.3-32.7) |
| Cameroon 2011 | 29.2 (27.3-31.2) | 11.2 (10.0-12.6) | 32.6 (30.7-34.6) |
| Cameroon 2018 | 20.3 (18.3-22.4) | 5.8 (5.0-6.8) | 21.9 (20.0-24.0) |
| Colombia 2005 | 19.4 (18.7-20.1) | 6.9 (6.4-7.3) | 20.9 (20.2-21.7) |
| Colombia 2010 | 18.6 (18.0-19.2) | 5.5 (5.2-5.8) | 19.7 (19.1-20.3) |
| Colombia 2015 | 16.5 (15.7-17.3) | 3.8 (3.5-4.1) | 17.5 (16.7-18.3) |
| Dominican Republic 2002 | 18.3 (16.9-19.8) | 6.5 (5.6-7.5) | 19.1 (17.7-20.7) |
| Dominican Republic 2007 | 10.7 (9.7-11.9) | 3.5 (2.9-4.3) | 11.5 (10.4-12.6) |
| Dominican Republic 2013 | 14.7 (13.4-16.2) | 4.2 (3.6-5.0) | 15.6 (14.2-17.1) |
| Haiti 2005-06 | 12.1 (10.1-14.4) | 10.1 (8.5-12.0) | 17.5 (15.2-20.2) |
| Haiti 2012 | 10.6 (9.4-11.8) | 8.7 (7.7-9.9) | 15.2 (13.8-16.7) |
| Haiti 2016-17 | 10.2 (9.0-11.6) | 7.2 (6.1-8.6) | 14.2 (12.6-15.8) |
| India 2005-06 | 21.3 (20.6-22.1) | 7.1 (6.7-7.6) | 23.8 (23.1-24.6) |
| India 2015-16 | 22.8 (22.1-23.5) | 5.7 (5.4-6.1) | 24.1 (23.4-24.9) |
| India 2019-21 | 22.8 (22.0-23.7) | 5.2 (4.8-5.6) | 23.9 (23.0-24.8) |
| Jordan 2007 | 12.2 (10.4-14.3) | 5.6 (4.5-7.0) | 14.6 (12.6-16.9) |
| Jordan 2012 | 11.2 (9.9-12.7) | 6.0 (5.0-7.0) | 14.1 (12.7-15.6) |
| Jordan 2017-18 | 12.7 (11.2-14.3) | 3.3 (2.7-4.2) | 13.8 (12.3-15.5) |
| Jordan 2023 | 7.6 (6.4-9.1) | 1.7 (1.1-2.4) | 8.3 (7.0-9.8) |
| Kenya 2008-09 | 29.1 (27.0-31.4) | 14.1 (12.6-15.7) | 31.7 (29.5-33.9) |
| Kenya 2014 | 22.1 (20.4-23.9) | 9.8 (8.6-11.2) | 25.2 (23.3-27.1) |
| Kenya 2022 | 18.2 (17.2-19.3) | 7.7 (7.0-8.5) | 21.0 (19.8-22.1) |
| Malawi 2004 | 13.1 (12.1-14.1) | 11.4 (10.4-12.6) | 19.5 (18.3-20.8) |
| Malawi 2010 | 15.1 (13.9-16.5) | 13.7 (12.4-15.0) | 22.3 (20.8-23.9) |
| Malawi 2015-16 | 16.5 (15.0-18.1) | 15.8 (14.5-17.1) | 24.6 (23.0-26.3) |
| Mali 2006 | 18.1 (16.6-19.7) | 4.1 (3.3-5.0) | 19.5 (17.9-21.3) |
| Mali 2012-13 | 20.7 (18.4-23.2) | 12.1 (10.3-14.2) | 26.6 (24.1-29.4) |
| Mali 2018 | 18.2 (16.3-20.2) | 7.9 (6.7-9.3) | 21.1 (19.0-23.3) |
| Mozambique 2011 | 25.9 (24.3-27.7) | 6.9 (6.0-7.9) | 27.7 (26.0-29.3) |
| Mozambique 2015 | 14.3 (12.6-16.2) | 2.8 (2.1-3.7) | 14.9 (13.2-16.7) |
| Mozambique 2022-23 | 15.5 (14.0-17.2) | 4.5 (3.8-5.3) | 17.0 (15.4-18.7) |
| Nepal 2011 | 10.7 (9.4-12.1) | 7.7 (6.7-9.0) | 14.3 (12.8-15.9) |
| Nepal 2016 | 10.0 (8.8-11.3) | 4.0 (3.2-4.9) | 11.2 (10.0-12.6) |
| Nepal 2022 | 11.8 (10.5-13.2) | 4.3 (3.6-5.0) | 13.0 (11.7-14.5) |
| Nigeria 2008 | 14.3 (13.5-15.2) | 3.2 (2.8-3.6) | 15.2 (14.4-16.1) |
| Nigeria 2013 | 9.4 (8.7-10.2) | 3.8 (3.4-4.2) | 11.1 (10.3-11.9) |
| Nigeria 2018 | 12.0 (11.0-13.0) | 4.7 (4.2-5.4) | 13.9 (12.9-15.1) |
| Peru 2004-06 | 12.8 (11.9-13.7) | 3.6 (3.1-4.0) | 13.7 (12.9-14.7) |
| Peru 2007-08 | 14.0 (12.9-15.0) | 3.7 (3.2-4.2) | 14.9 (13.9-16.1) |
| Peru 2009 | 13.5 (12.7-14.3) | 3.2 (2.9-3.6) | 14.2 (13.4-15.0) |
| Peru 2010 | 13.0 (12.2-13.8) | 3.4 (3.0-3.8) | 13.9 (13.1-14.7) |
| Peru 2011 | 12.6 (11.9-13.4) | 3.3 (2.9-3.7) | 13.6 (12.9-14.4) |
| Peru 2012 | 12.1 (11.4-13.0) | 3.2 (2.9-3.7) | 12.9 (12.1-13.7) |
| Philippines 2008 | 7.3 (6.6-8.1) | 4.8 (4.3-5.4) | 10.0 (9.2-10.9) |
| Philippines 2013 | 5.3 (4.8-5.9) | 3.3 (2.9-3.8) | 7.1 (6.5-7.8) |
| Philippines 2017 | 4.3 (3.8-4.9) | 2.2 (1.9-2.6) | 5.4 (4.8-6.1) |
| Philippines 2022 | 3.4 (2.9-3.9) | 1.4 (1.1-1.7) | 4.1 (3.6-4.7) |
| Rwanda 2010 | 22.4 (20.9-24.1) | 12.7 (11.5-14.0) | 26.9 (25.2-28.6) |
| Rwanda 2014-15 | 17.4 (15.5-19.4) | 7.6 (6.3-9.2) | 20.1 (18.0-22.3) |
| Rwanda 2019-20 | 19.4 (17.5-21.6) | 9.3 (7.9-11.0) | 23.4 (21.3-25.7) |
| Senegal 2017 | 8.9 (7.4-10.8) | 5.9 (4.5-7.7) | 12.2 (10.3-14.3) |

| | | | |
|---|---|---|---|
| Senegal 2018 | 5.2 (3.9-7.0) | 4.0 (2.6-6.1) | 7.6 (5.8-9.8) |
| Senegal 2019 | 4.6 (3.3-6.5) | 3.0 (1.8-4.7) | 6.1 (4.5-8.1) |
| Tanzania 2010 | 32.2 (30.4-34.1) | 13.2 (12.0-14.5) | 35.3 (33.5-37.1) |
| Tanzania 2015-16 | 26.8 (25.3-28.4) | 10.4 (9.5-11.4) | 29.4 (27.8-31.0) |
| Tanzania 2022 | 23.6 (21.8-25.4) | 8.8 (7.6-10.3) | 26.1 (24.3-27.9) |
| Uganda 2006 | 34.9 (31.6-38.4) | 24.8 (22.3-27.4) | 44.8 (41.5-48.1) |
| Uganda 2011 | 24.9 (21.7-28.5) | 20.9 (18.4-23.8) | 34.6 (31.1-38.2) |
| Uganda 2016 | 22.9 (21.6-24.3) | 16.8 (15.6-18.1) | 30.3 (28.8-31.9) |
| Zambia 2007 | 38.6 (36.4-40.8) | 16.2 (14.8-17.7) | 42.0 (39.8-44.3) |
| Zambia 2013-14 | 21.2 (20.0-22.4) | 13.0 (12.0-14.0) | 26.5 (25.2-27.9) |
| Zambia 2018 | 20.8 (19.5-22.2) | 10.7 (9.5-12.0) | 25.0 (23.3-26.7) |
| Zimbabwe 2005-05 | 22.6 (21.0-24.3) | 11.5 (10.4-12.7) | 27.5 (25.8-29.2) |
| Zimbabwe 2010-11 | 20.7 (19.2-22.2) | 13.3 (12.1-14.5) | 27.2 (25.7-28.7) |
| Zimbabwe 2015 | 15.2 (14.0-16.5) | 9.3 (8.3-10.4) | 19.8 (18.4-21.3) |

**Figure S2: Predicted prevalence of any recent physical IPV, by country**

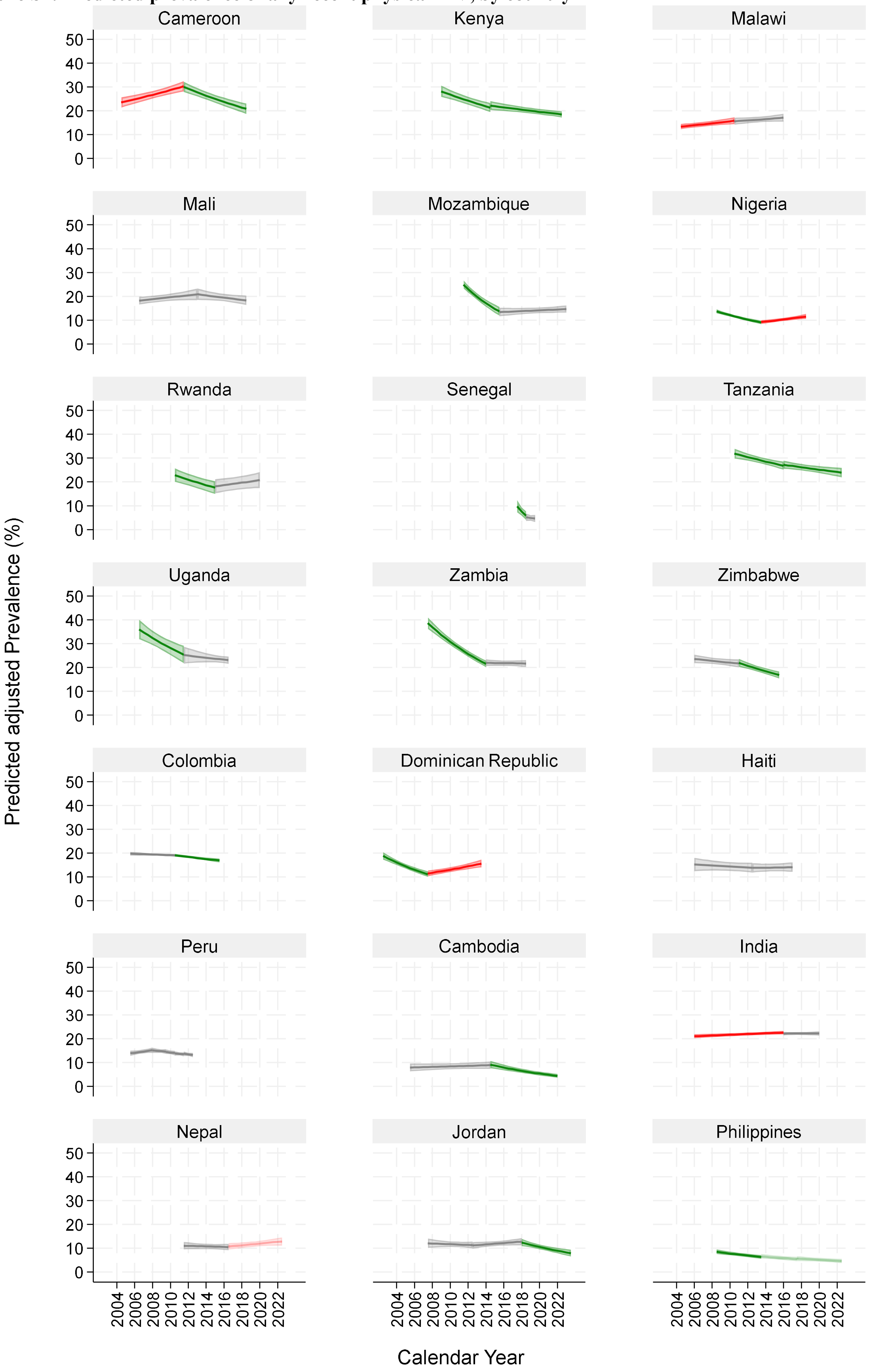

Notes: Countries are ordered by WHO region and alphabetically within region. The prevalence and 95% confidence intervals are for any recent (12-month) physical IPV, from log binomial regression models adjusted for five-year age groups. Green is where there is strong evidence of a decline in the period (p<0.01); lighter green is weak evidence of a decline (p<0.05); red is strong evidence of an increase in the period (p<0.01); lighter red is weak evidence of an increase (p<0.05); grey there is no evidence of a change in prevalence over the period.

**Figure S3: Predicted prevalence of any recent sexual physical IPV, by country**

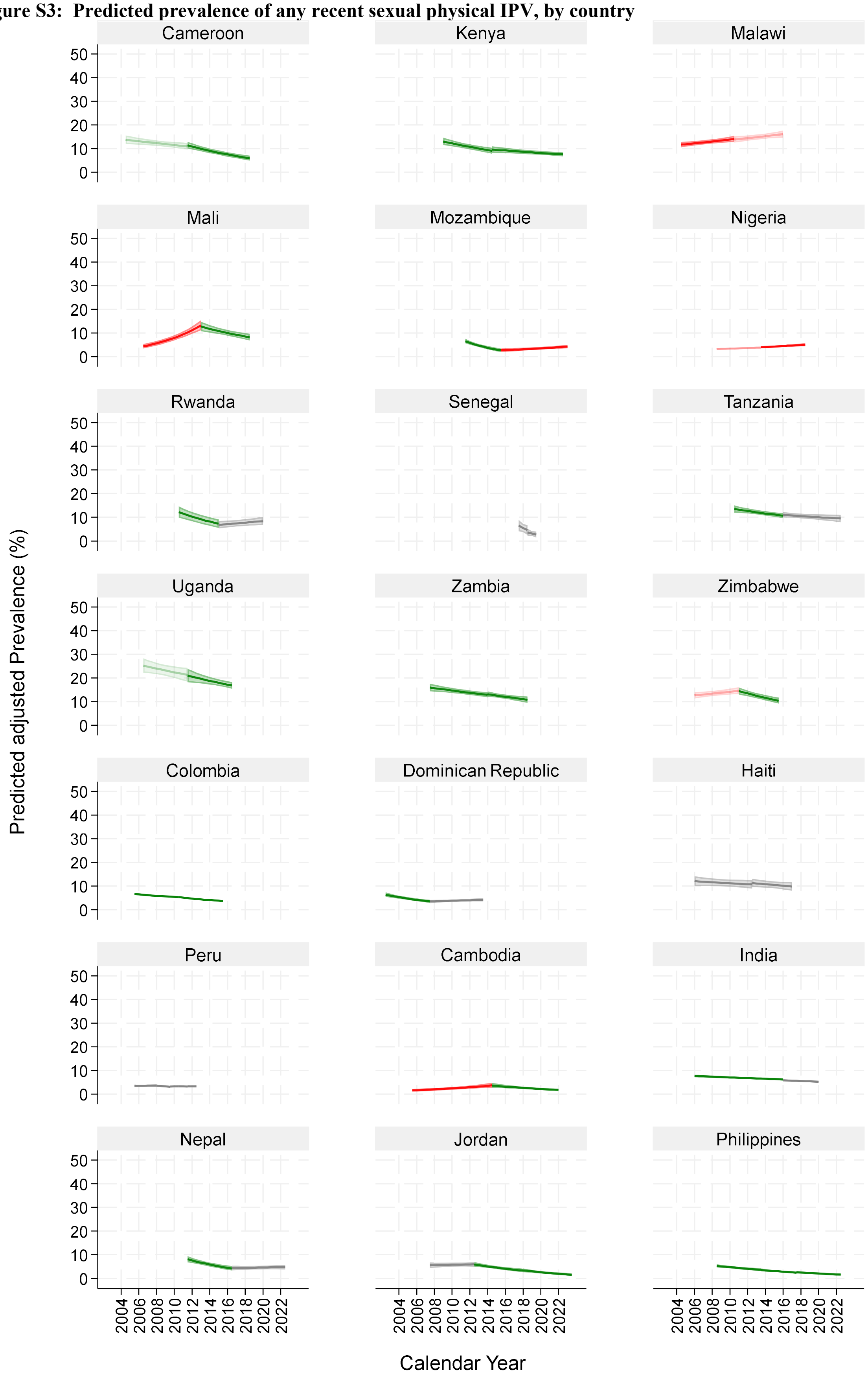

Notes: Countries are ordered by WHO region and alphabetically within region. The prevalence and 95% confidence intervals are for any recent (12-month) sexual IPV, from log binomial regression models adjusted for five-year age groups. Green is where there is strong evidence of a decline in the period (p<0.01); lighter green is weak evidence of a decline (p<0.05); red is strong evidence of an increase in the period (p<0.01); lighter red is weak evidence of an increase (p<0.05); grey there is no evidence of a change in prevalence over the period.

**Table S7: Risk ratios for recent IPV outcomes trends, by country, adjusted for five-year age group.**

| Country | Survey | 12-month physical IPV | | 12-month sexual IPV | | 12-month physical and/or sexual IPV | |
|---|---|---|---|---|---|---|---|
| | | RR | 95% CI | RR | 95% CI | RR | 95% CI |
| *African Region* | | | | | | | |
| Cameroon | Cameroon 2004 | 1 | | 1 | | 1 | |
| | Cameroon 2011 | 1.29 | (1.15-1.44) | 0.8 | (0.67-0.95) | 1.08 | (0.98-1.19) |
| | Cameroon 2018 | 0.89 | (0.78-1.02) | 0.41 | (0.33-0.51) | 0.72 | (0.64-0.82) |
| Kenya | Kenya 2008-09 | 1 | | 1 | | 1 | |
| | Kenya 2014 | 0.76 | (0.68-0.85) | 0.7 | (0.59-0.83) | 0.79 | (0.72-0.88) |
| | Kenya 2022 | 0.63 | (0.57-0.69) | 0.55 | (0.47-0.63) | 0.67 | (0.61-0.73) |
| Malawi | Malawi 2004 | 1 | | 1 | | 1 | |
| | Malawi 2010 | 1.18 | (1.05-1.32) | 1.2 | (1.05-1.37) | 1.16 | (1.05-1.27) |
| | Malawi 2015-16 | 1.28 | (1.14-1.44) | 1.39 | (1.23-1.58) | 1.28 | (1.17-1.41) |
| Mali | Mali 2006 | 1 | | 1 | | 1 | |
| | Mali 2012-13 | 1.14 | (0.99-1.32) | 3.03 | (2.34-3.93) | 1.37 | (1.20-1.56) |
| | Mali 2018 | 1 | (0.87-1.16) | 1.96 | (1.50-2.55) | 1.08 | (0.94-1.24) |
| Mozambique | Mozambique 2011 | 1 | | 1 | | 1 | |
| | Mozambique 2015 | 0.55 | (0.48-0.64) | 0.41 | (0.30-0.57) | 0.54 | (0.47-0.62) |
| | Mozambique 2022-23 | 0.6 | (0.53-0.67) | 0.66 | (0.53-0.82) | 0.62 | (0.55-0.69) |
| Nigeria | Nigeria 2008 | 1 | | 1 | | 1 | |
| | Nigeria 2013 | 0.66 | (0.59-0.73) | 1.19 | (1.01-1.41) | 0.73 | (0.66-0.81) |
| | Nigeria 2018 | 0.83 | (0.74-0.93) | 1.51 | (1.27-1.80) | 0.91 | (0.83-1.01) |
| Rwanda | Rwanda 2010 | 1 | | 1 | | 1 | |
| | Rwanda 2014-15 | 0.77 | (0.67-0.88) | 0.6 | (0.48-0.74) | 0.74 | (0.66-0.84) |
| | Rwanda 2019-20 | 0.88 | (0.77-1.00) | 0.73 | (0.60-0.89) | 0.88 | (0.78-0.99) |
| Senegal | Senegal 2017 | 1 | | 1 | | 1 | |
| | Senegal 2018 | 0.59 | (0.42-0.84) | 0.69 | (0.42-1.16) | 0.63 | (0.46-0.87) |
| | Senegal 2019 | 0.51 | (0.34-0.76) | 0.51 | (0.30-0.86) | 0.5 | (0.36-0.70) |
| Tanzania | Tanzania 2010 | 1 | | 1 | | 1 | |
| | Tanzania 2015-16 | 0.84 | (0.77-0.92) | 0.79 | (0.69-0.91) | 0.84 | (0.77-0.91) |
| | Tanzania 2022 | 0.74 | (0.66-0.81) | 0.68 | (0.56-0.82) | 0.74 | (0.68-0.82) |
| Uganda | Uganda 2006 | 1 | | 1 | | 1 | |
| | Uganda 2011 | 0.71 | (0.60-0.85) | 0.85 | (0.72-1.00) | 0.77 | (0.68-0.88) |
| | Uganda 2016 | 0.65 | (0.58-0.74) | 0.68 | (0.60-0.77) | 0.68 | (0.62-0.74) |
| Zambia | Zambia 2007 | 1 | | 1 | | 1 | |
| | Zambia 2013-14 | 0.56 | (0.51-0.61) | 0.81 | (0.71-0.91) | 0.64 | (0.59-0.69) |
| | Zambia 2018 | 0.55 | (0.50-0.60) | 0.67 | (0.57-0.78) | 0.6 | (0.55-0.66) |
| Zimbabwe | Zimbabwe 2005-05 | 1 | | 1 | | 1 | |
| | Zimbabwe 2010-11 | 0.92 | (0.83-1.02) | 1.15 | (1.01-1.32) | 1 | (0.92-1.08) |
| | Zimbabwe 2015 | 0.7 | (0.63-0.78) | 0.83 | (0.71-0.96) | 0.75 | (0.68-0.83) |

***Table S7 Continued…***

| country | survey | Recent Physical IPV | | Recent Sexual IPV | | Recent Physical and Sexual IPV | |
|---|---|---|---|---|---|---|---|
| | | RR | 95% CI | RR | 95% CI | RR | 95% CI |
| *Region of the Americas* | | | | | | | |
| Colombia | Colombia 2005 | 1 | | 1 | | 1 | |
| | Colombia 2010 | 0.96 | (0.92-1.01) | 0.8 | (0.73-0.87) | 0.95 | (0.90-0.99) |
| | Colombia 2015 | 0.85 | (0.80-0.91) | 0.55 | (0.49-0.61) | 0.84 | (0.79-0.89) |
| Dominican Republic | Dominican Republic 2002 | 1 | | 1 | | 1 | |
| | Dominican Republic 2007 | 0.59 | (0.52-0.68) | 0.55 | (0.43-0.70) | 0.61 | (0.54-0.69) |
| | Dominican Republic 2013 | 0.81 | (0.71-0.92) | 0.66 | (0.53-0.82) | 0.82 | (0.73-0.93) |
| Haiti | Haiti 2005-06 | 1 | | 1 | | 1 | |
| | Haiti 2012 | 0.91 | (0.74-1.12) | 0.89 | (0.72-1.10) | 0.9 | (0.76-1.06) |
| | Haiti 2016-17 | 0.92 | (0.74-1.15) | 0.76 | (0.60-0.97) | 0.87 | (0.73-1.05) |
| Peru | Peru 2004-06 | 1 | | 1 | | 1 | |
| | Peru 2007-08 | 1.09 | (0.98-1.21) | 1.04 | (0.86-1.25) | 1.08 | (0.98-1.20) |
| | Peru 2009 | 1.05 | (0.96-1.16) | 0.91 | (0.77-1.08) | 1.03 | (0.94-1.13) |
| | Peru 2010 | 1.01 | (0.92-1.11) | 0.95 | (0.80-1.13) | 1.01 | (0.92-1.11) |
| | Peru 2011 | 0.99 | (0.90-1.09) | 0.92 | (0.77-1.10) | 0.99 | (0.91-1.09) |
| | Peru 2012 | 0.95 | (0.86-1.05) | 0.91 | (0.76-1.09) | 0.94 | (0.85-1.03) |
| *South-East Asian Region* | | | | | | | |
| Cambodia | Cambodia 2005 | 1 | | 1 | | 1 | |
| | Cambodia 2014 | 1.13 | (0.89-1.45) | 2.38 | (1.49-3.78) | 1.22 | (0.97-1.54) |
| | Cambodia 2021-22 | 0.55 | (0.43-0.70) | 1.16 | (0.74-1.81) | 0.61 | (0.49-0.77) |
| India | India 2005-06 | 1 | | 1 | | 1 | |
| | India 2015-16 | 1.06 | (1.01-1.11) | 0.81 | (0.74-0.88) | 1.01 | (0.96-1.05) |
| | India 2019-21 | 1.07 | (1.01-1.13) | 0.74 | (0.68-0.82) | 1 | (0.95-1.05) |
| Nepal | Nepal 2011 | 1 | | 1 | | 1 | |
| | Nepal 2016 | 0.95 | (0.79-1.13) | 0.52 | (0.40-0.68) | 0.8 | (0.68-0.93) |
| | Nepal 2022 | 1.13 | (0.95-1.35) | 0.57 | (0.45-0.71) | 0.93 | (0.80-1.09) |
| *Eastern Mediterranean Region* | | | | | | | |
| Jordan | Jordan 2007 | 1 | | 1 | | 1 | |
| | Jordan 2012 | 0.93 | (0.76-1.14) | 1.06 | (0.81-1.39) | 0.98 | (0.81-1.17) |
| | Jordan 2017-18 | 1.06 | (0.87-1.30) | 0.6 | (0.44-0.82) | 0.96 | (0.80-1.16) |
| | Jordan 2023 | 0.7 | (0.54-0.89) | 0.31 | (0.20-0.47) | 0.62 | (0.49-0.78) |
| *Western Pacific Region* | | | | | | | |
| Philippines | Philippines 2008 | 1 | | 1 | | 1 | |
| | Philippines 2013 | 0.74 | (0.64-0.85) | 0.69 | (0.58-0.82) | 0.72 | (0.63-0.81) |
| | Philippines 2017 | 0.61 | (0.52-0.72) | 0.47 | (0.39-0.57) | 0.55 | (0.48-0.64) |
| | Philippines 2022 | 0.5 | (0.41-0.59) | 0.3 | (0.23-0.38) | 0.44 | (0.37-0.51) |

**Table S8: Average marginal effects for intersurvey periods showing the change in recent physical and/or sexual IPV**

| | Country | Midpoint of earlier survey | midpoint of later survey | Marginal effect | 95 % CI | p-value |
|---|---|---|---|---|---|---|
| **African Region** | | | | | | |
| | Cameroon | 2004 | 2011 | 0.32 | (-0.12- 0.76) | 0.150 |
| | | 2011 | 2018 | -1.5 | (-1.88--1.11) | 0.000 |
| | Kenya | 2008.5 | 2014 | -1.18 | (-1.71--0.66) | 0.000 |
| | | 2014 | 2022 | -0.49 | (-0.75--0.22) | 0.000 |
| | Malawi | 2004 | 2010 | 0.53 | ( 0.19- 0.86) | 0.002 |
| | | 2010 | 2015.5 | 0.45 | ( 0.04- 0.87) | 0.034 |
| | Mali | 2006 | 2012.5 | 1.06 | ( 0.63- 1.50) | 0.000 |
| | | 2012.5 | 2018 | -1.02 | (-1.64--0.40) | 0.001 |
| | Mozambique | 2011 | 2015 | -3.37 | (-4.08--2.66) | 0.000 |
| | | 2015 | 2022.5 | 0.28 | (-0.05- 0.61) | 0.093 |
| | Nigeria | 2008 | 2013 | -0.79 | (-1.02--0.56) | 0.000 |
| | | 2013 | 2018 | 0.53 | ( 0.28- 0.79) | 0.000 |
| | Rwanda | 2005 | 2010 | 4.08 | ( 3.52- 4.63) | 0.000 |
| | | 2010 | 2014.5 | -5.88 | (-6.58--5.18) | 0.000 |
| | | 2014.5 | 2019.5 | 0.78 | ( 0.11- 1.46) | 0.023 |
| | Senegal | 2017 | 2018 | -4.97 | (-8.45--1.50) | 0.005 |
| | | 2018 | 2019 | -1.36 | (-3.93- 1.22) | 0.301 |
| | Tanzania | 2010 | 2015.5 | -1.03 | (-1.51--0.56) | 0.000 |
| | | 2015.5 | 2022 | -0.49 | (-0.88--0.11) | 0.013 |
| | Uganda | 2006 | 2011 | -2.08 | (-3.05--1.11) | 0.000 |
| | | 2011 | 2016 | -0.83 | (-1.58--0.07) | 0.031 |
| | Zambia | 2007 | 2013.5 | -2.23 | (-2.62--1.83) | 0.000 |
| | | 2013.5 | 2018 | -0.33 | (-0.83- 0.17) | 0.202 |
| | Zimbabwe | 2005.5 | 2010.5 | -0.05 | (-0.51- 0.41) | 0.831 |
| | | 2010.5 | 2015 | -1.51 | (-1.99--1.04) | 0.000 |
| **Region of the Americas** | | | | | | |
| | Colombia | 2005 | 2010 | -0.23 | (-0.43--0.03) | 0.021 |
| | | 2010 | 2015 | -0.48 | (-0.69--0.27) | 0.000 |
| | Dominican Republic | 2002 | 2007 | -1.52 | (-1.89--1.15) | 0.000 |
| | | 2007 | 2013 | 0.71 | ( 0.40- 1.01) | 0.000 |
| | Haiti | 2005.5 | 2012 | -0.36 | (-0.83- 0.12) | 0.139 |
| | | 2012 | 2016.5 | -0.05 | (-0.62- 0.53) | 0.868 |
| | Peru | 2005 | 2007.5 | 0.52 | (-0.10- 1.14) | 0.098 |
| | | 2007.5 | 2009 | -0.56 | (-1.53- 0.41) | 0.256 |
| | | 2009 | 2010 | -0.34 | (-1.52- 0.84) | 0.569 |
| | | 2010 | 2011 | -0.21 | (-1.40- 0.98) | 0.729 |
| | | 2011 | 2012 | -0.73 | (-1.97- 0.51) | 0.250 |
| **South-East Asian Region** | | | | | | |
| | Cambodia | 2005 | 2014 | 0.21 | (-0.05- 0.46) | 0.120 |
| | | 2014 | 2021.5 | -0.68 | (-0.89--0.46) | 0.000 |
| | India | 2005.5 | 2015.5 | 0.05 | (-0.05- 0.16) | 0.334 |
| | | 2015.5 | 2019.5 | -0.07 | (-0.35- 0.21) | 0.614 |

| | | | | |
|---|---|---|---|---|
| Nepal | 2011 | 2016 | -0.61 (-1.03--0.18) | 0.005 |
| | 2016 | 2022 | 0.35 ( 0.00- 0.71) | 0.050 |
| **Eastern Mediterranean Region** | | | | |
| Jordan | 2007 | 2012 | -0.06 (-0.58- 0.46) | 0.817 |
| | 2012 | 2017.5 | -0.03 (-0.43- 0.37) | 0.886 |
| | 2017.5 | 2023 | -0.97 (-1.44--0.50) | 0.000 |
| **Western Pacific Region** | | | | |
| Philippines | 2008 | 2013 | -0.61 (-0.85--0.38) | 0.000 |
| | 2013 | 2017 | -0.46 (-0.72--0.21) | 0.000 |
| | 2017 | 2022 | -0.28 (-0.50--0.07) | 0.009 |

**Figure S4: Trends in age adjusted risk ratios (RR) for recent physical and/or sexual IPV by age group 15-24 (blue) and 25-49 (green)**

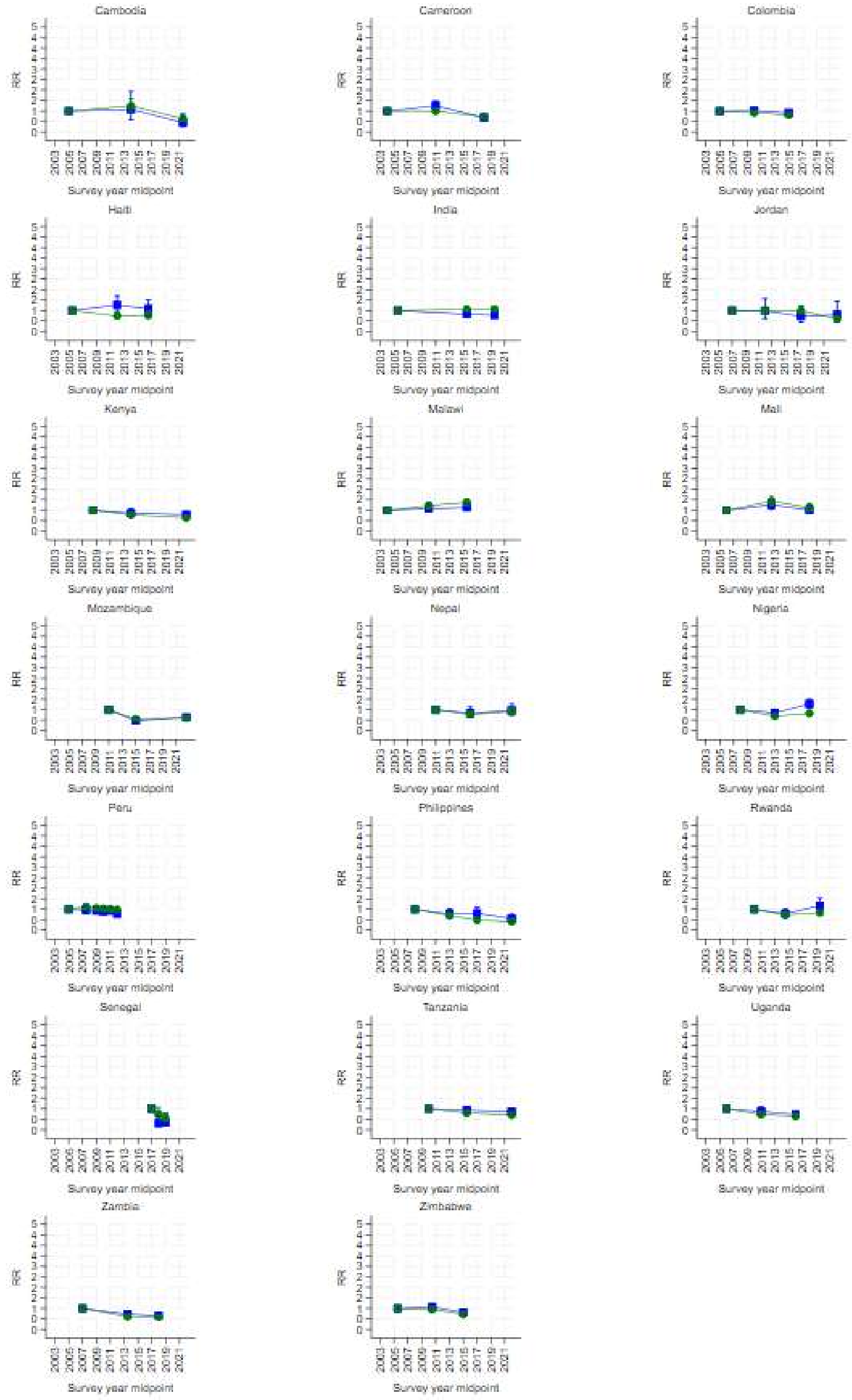

**Figure S5: Trends in age adjusted risk ratios for recent physical and/or sexual IPV, by no education (blue), primary education (green), and secondary or higher (red)**

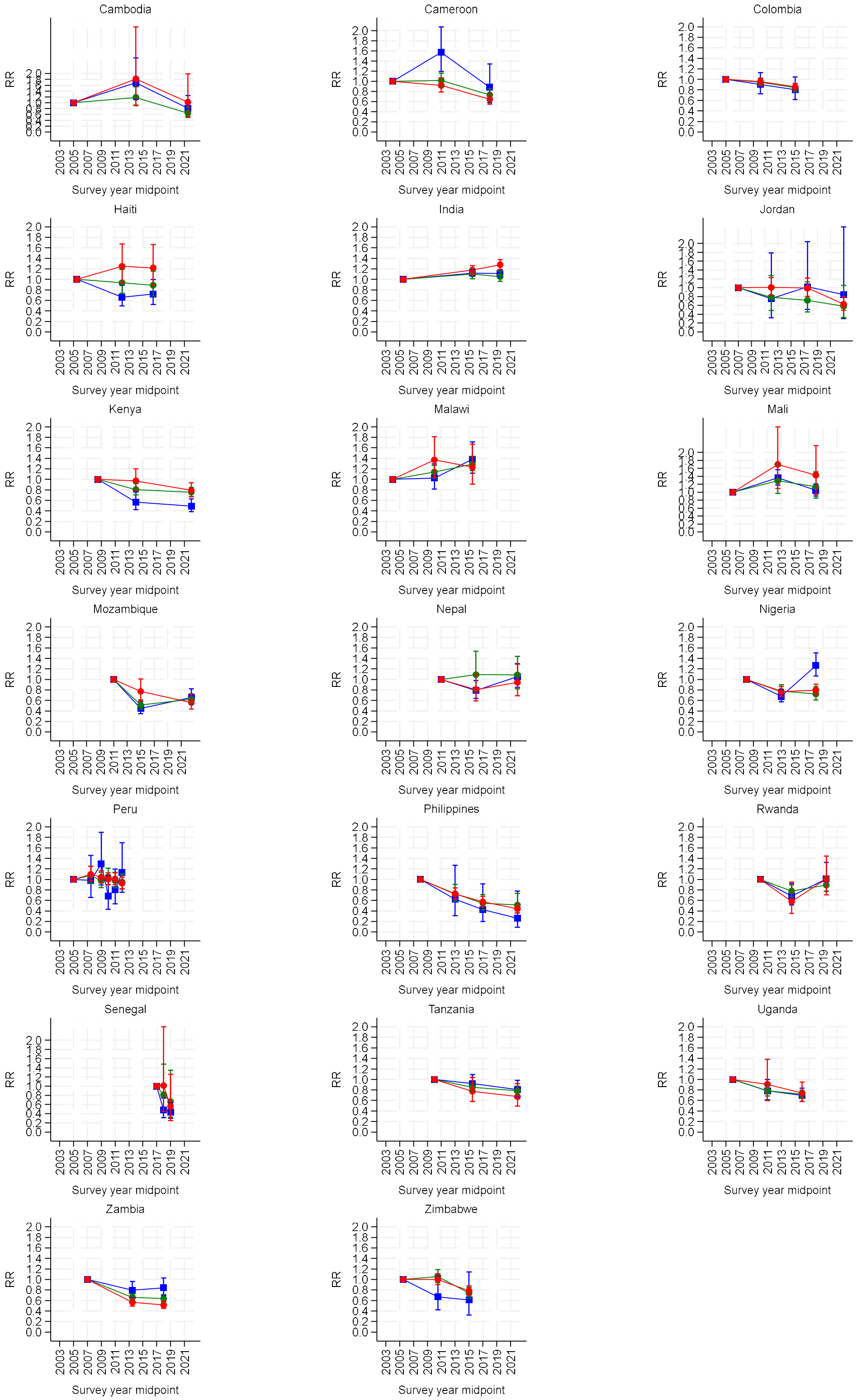

**Figure S6: Trends in age adjusted risk ratios for recent physical and/or sexual IPV, by rural (green) and urban (blue)**

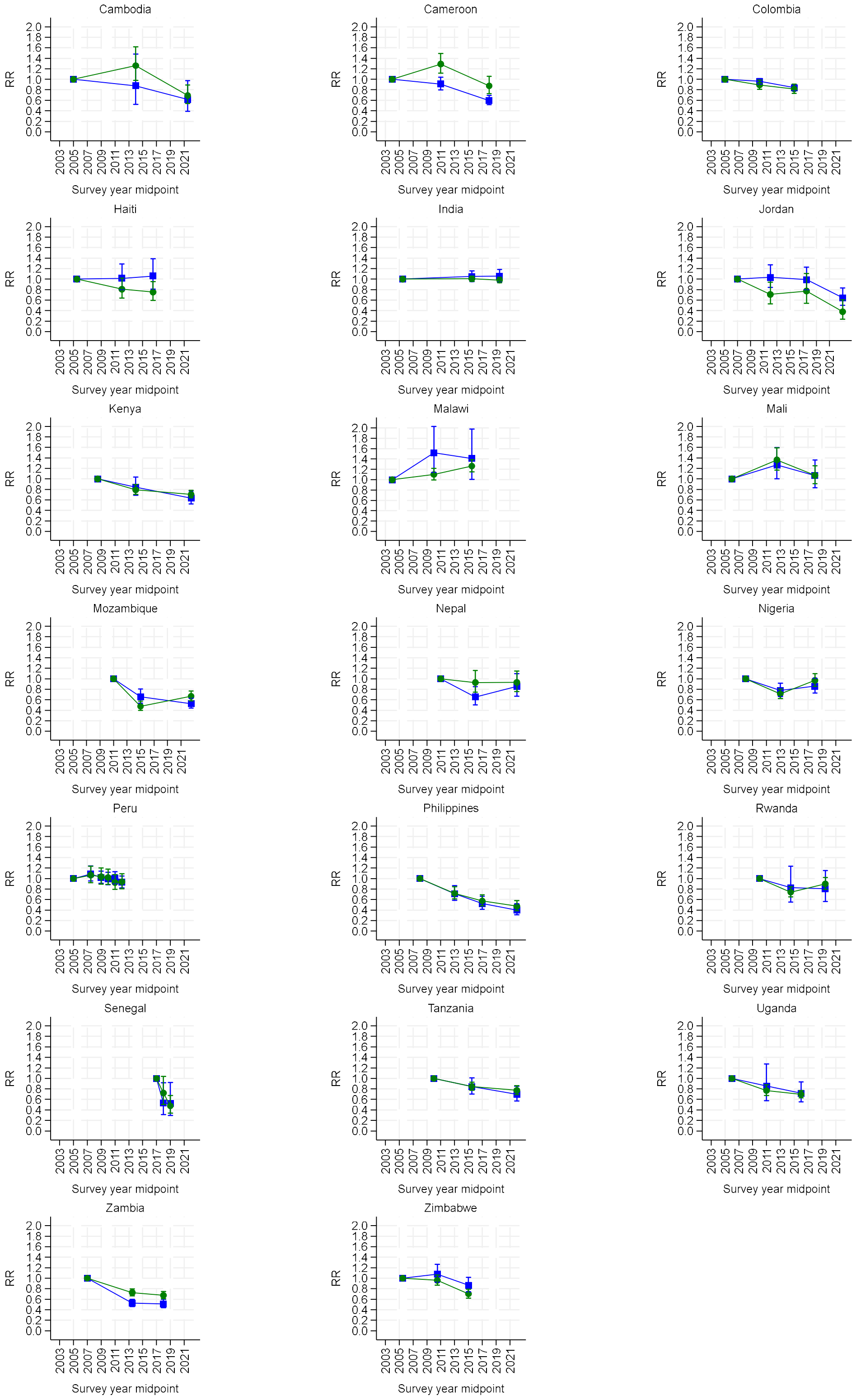

**Figure S7: Trends in age adjusted risk ratios for recent physical and/or sexual IPV, by richest two wealth quintiles – (dark blue), middle wealth quintile (grey), and poorest two wealth quintiles (light blue)**

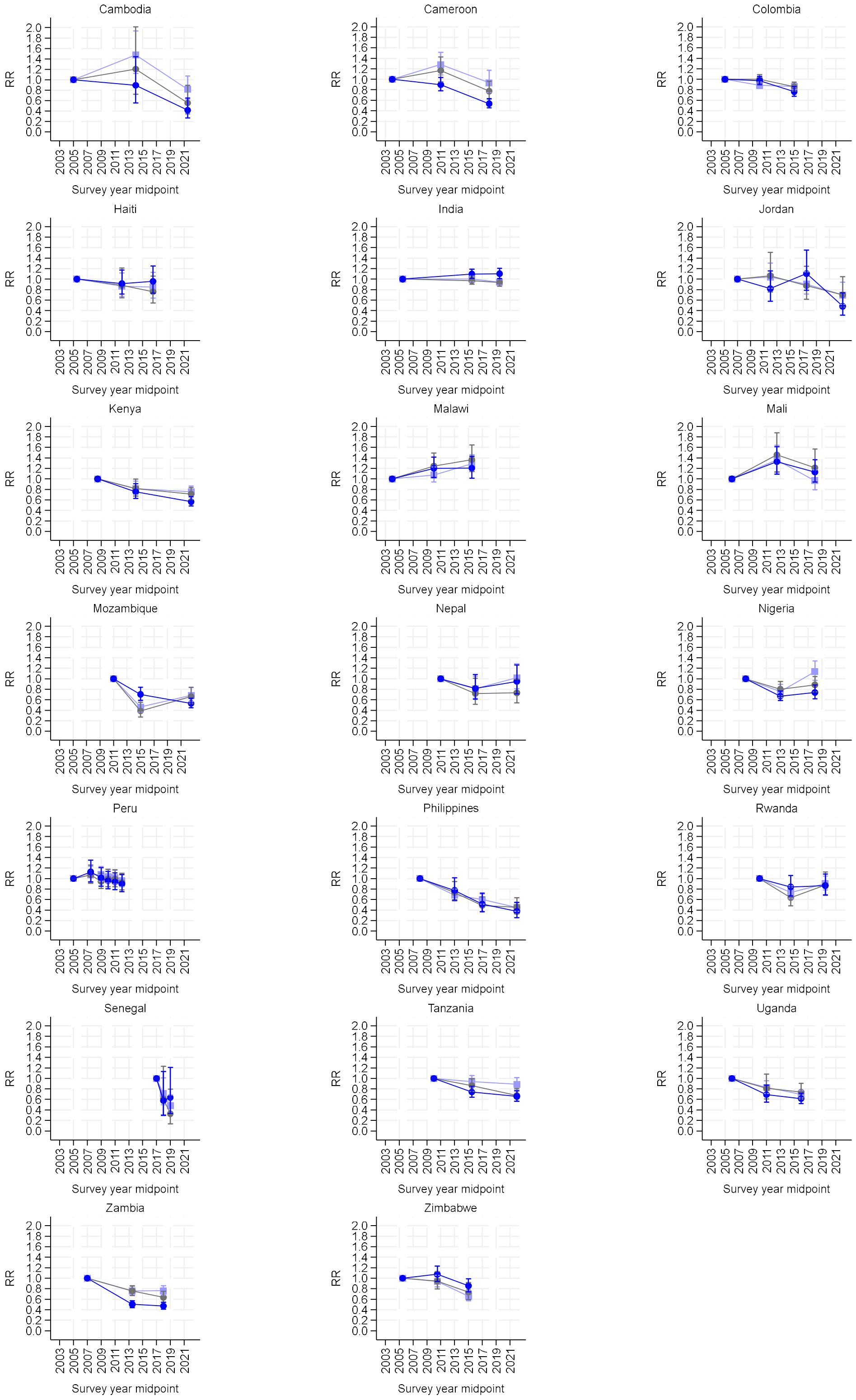

**Table S9: Stratified risk ratio of physical and/or sexual IPV by broad age group and country**

| Country | Survey year | 15-24 RR (95% CI) | | 25-49 RR (95% CI) | | | Wald test for interaction between broad age group and survey Wald F | P-value |
|---|---|---|---|---|---|---|---|---|
| Cambodia | | | | | | | | |
| | 2005 | 1 | | 1 | | | 9.95 | 0.000 |
| | 2014 | 1.08 (0.60-1.93) | | 1.25 | (0.98-1.59) | | | |
| | 2021-22 | 0.46 (0.24-0.86) | * | 0.64 | (0.50-0.81) | *** | | |
| Cameroon | | | | | | | | |
| | 2004 | 1 | | 1 | | | 12.77 | 0.000 |
| | 2011 | 1.26 (1.08-1.47) | ** | 1 | (0.88-1.13) | | | |
| | 2018 | 0.69 (0.57-0.84) | *** | 0.73 | (0.63-0.84) | *** | | |
| Colombia | | | | | | | | |
| | 2005 | 1 | | 1 | | | 13.16 | 0.000 |
| | 2010 | 1.02 (0.93-1.12) | | 0.92 | (0.88-0.97) | ** | | |
| | 2015 | 0.91 (0.82-1.02) | | 0.81 | (0.76-0.87) | *** | | |
| Haiti | | | | | | | | |
| | 2005-06 | 1 | | 1 | | | 3.06 | 0.009 |
| | 2012 | 1.27 (0.95-1.70) | | 0.75 | (0.62-0.92) | ** | | |
| | 2016-17 | 1.09 (0.78-1.51) | | 0.79 | (0.64-0.97) | * | | |
| India | | | | | | | | |
| | 2005-06 | 1 | | 1 | | | 10.82 | 0.000 |
| | 2015-16 | 0.83 (0.77-0.90) | *** | 1.07 | (1.02-1.12) | ** | | |
| | 2019-21 | 0.79 (0.71-0.88) | *** | 1.07 | (1.02-1.12) | * | | |
| Jordan | | | | | | | | |
| | 2007 | 1 | | 1 | | | 3.49 | 0.001 |
| | 2012 | 0.98 (0.62-1.56) | | 0.98 | (0.81-1.18) | | | |
| | 2017-18 | 0.74 (0.45-1.23) | | 1 | (0.82-1.22) | | | |
| | 2023 | 0.81 (0.46-1.43) | | 0.61 | (0.48-0.77) | *** | | |
| Kenya | | | | | | | | |
| | 2008-09 | 1 | | 1 | | | 16.55 | 0.000 |
| | 2014 | 0.86 (0.69-1.07) | | 0.78 | (0.69-0.87) | *** | | |
| | 2022 | 0.79 (0.66-0.94) | ** | 0.64 | (0.58-0.70) | *** | | |
| Malawi | | | | | | | | |
| | 2004 | 1 | | 1 | | | 6.49 | 0.000 |
| | 2010 | 1.08 (0.93-1.25) | | 1.21 | (1.08-1.36) | ** | | |
| | 2015-16 | 1.13 (0.99-1.30) | | 1.38 | (1.23-1.54) | *** | | |
| Mali | | | | | | | | |
| | 2006 | 1 | | 1 | | | 4.83 | 0.000 |
| | 2012-13 | 1.25 (1.04-1.50) | * | 1.43 | (1.23-1.66) | *** | | |
| | 2018 | 1.02 (0.84-1.24) | | 1.12 | (0.95-1.31) | | | |
| Mozambique | | | | | | | | |
| | 2011 | 1 | | 1 | | | 25.45 | 0.000 |
| | 2015 | 0.47 (0.36-0.61) | *** | 0.57 | (0.49-0.66) | *** | | |
| | 2022-23 | 0.64 (0.51-0.79) | *** | 0.61 | (0.54-0.69) | *** | | |
| Nepal | | | | | | | | |
| | 2011 | 1 | | 1 | | | 1.73 | 0.125 |
| | 2016 | 0.84 (0.62-1.13) | | 0.78 | (0.64-0.94) | ** | | |
| | 2022 | 0.97 (0.72-1.29) | | 0.92 | (0.77-1.09) | | | |
| Nigeria | | | | | | | | |
| | 2008 | 1 | | 1 | | | 12.55 | 0.000 |
| | 2013 | 0.85 (0.72-1.00) | | 0.7 | (0.63-0.77) | *** | | |
| | 2018 | 1.27 (1.08-1.51) | ** | 0.83 | (0.74-0.93) | ** | | |
| Peru | | | | | | | | |
| | 2004-06 | 1 | | 1 | | | 2.94 | 0.001 |
| | 2007-08 | 0.97 (0.80-1.18) | | 1.12 | (1.00-1.25) | * | | |

| | | | | | | | | |
|---|---|---|---|---|---|---|---|---|
| | 2009 | 0.93 (0.78-1.12) | | 1.06 | (0.96-1.17) | | | |
| | 2010 | 0.9 (0.75-1.09) | | 1.04 | (0.94-1.15) | | | |
| | 2011 | 0.93 (0.77-1.13) | | 1.01 | (0.91-1.12) | | | |
| | 2012 | 0.77 (0.64-0.95) | * | 0.99 | (0.89-1.09) | | | |
| Philippines | | | | | | | | |
| | 2008 | 1 | | 1 | | | 23.29 | 0.000 |
| | 2013 | 0.78 (0.61-1.01) | | 0.7 | (0.61-0.81) | *** | | |
| | 2017 | 0.81 (0.59-1.10) | | 0.49 | (0.42-0.57) | *** | | |
| | 2022 | 0.56 (0.41-0.76) | *** | 0.41 | (0.34-0.49) | *** | | |
| Rwanda | | | | | | | | |
| | 2010 | 1 | | 1 | | | 5.63 | 0.000 |
| | 2014-15 | 0.79 (0.61-1.03) | | 0.74 | (0.64-0.85) | *** | | |
| | 2019-20 | 1.17 (0.89-1.53) | | 0.84 | (0.74-0.95) | ** | | |
| Senegal | | | | | | | | |
| | 2017 | 1 | | 1 | | | 5.32 | 0.000 |
| | 2018 | 0.31 (0.15-0.64) | ** | 0.75 | (0.54-1.06) | | | |
| | 2019 | 0.36 (0.18-0.75) | ** | 0.56 | (0.39-0.80) | ** | | |
| Tanzania | | | | | | | | |
| | 2010 | 1 | | 1 | | | 9.11 | 0.000 |
| | 2015-16 | 0.94 (0.81-1.08) | | 0.81 | (0.73-0.88) | *** | | |
| | 2022 | 0.87 (0.73-1.03) | | 0.7 | (0.63-0.79) | *** | | |
| Uganda | | | | | | | | |
| | 2006 | 1 | | 1 | | | 15.38 | 0.000 |
| | 2011 | 0.87 (0.70-1.10) | | 0.73 | (0.63-0.84) | *** | | |
| | 2016 | 0.73 (0.62-0.87) | *** | 0.65 | (0.59-0.72) | *** | | |
| Zambia | | | | | | | | |
| | 2007 | 1 | | 1 | | | 32.79 | 0.000 |
| | 2013-14 | 0.73 (0.64-0.82) | *** | 0.61 | (0.55-0.67) | *** | | |
| | 2018 | 0.66 (0.57-0.76) | *** | 0.58 | (0.53-0.64) | *** | | |
| Zimbabwe | | | | | | | | |
| | 2005-05 | 1 | | 1 | | | 9.94 | 0.000 |
| | 2010-11 | 1.06 (0.93-1.22) | | 0.95 | (0.87-1.05) | | | |
| | 2015 | 0.81 (0.70-0.95) | ** | 0.72 | (0.64-0.80) | *** | | |

Notes: Estimates are adjusted for five-year age group, with Wald F-statistic for a test of interaction between survey and broad age group; CI = confidence interval, RR = risk ratio. * P-value<0.05, **p-value<0.01, ***p-value<0.001***

**Table S10: Stratified risk ratio of physical and/or sexual IPV by education and country**

| | Survey year | Education: None RR (95% CI) | | Primary RR (95% CI) | | | Secondary+ RR (95% CI) | | Wald test for interaction between education group and survey Wald F | P-value |
|---|---|---|---|---|---|---|---|---|---|---|
| Cambodia | | | | | | | | | | |
| | 2005 | 1 | | 1 | | | 1 | | 12.75 | 0.000 |
| | 2014 | 1.69(1.12-2.54) | * | 1.18 | (0.90-1.55) | | 1.82(0.92-3.60) | | | |
| | 2021-22 | 0.81(0.52-1.25) | | 0.65 | (0.50-0.84) | ** | 1.02(0.52-2.00) | | | |
| Cameroon | | | | | | | | | | |
| | 2004 | 1 | | 1 | | | 1 | | 11.35 | 0.000 |
| | 2011 | 1.57(1.19-2.08) | ** | 1.02 | (0.89-1.16) | | 0.92(0.79-1.07) | | | |
| | 2018 | 0.88(0.58-1.34) | | 0.73 | (0.63-0.85) | *** | 0.64(0.55-0.76) | *** | | |
| Colombia | | | | | | | | | | |
| | 2005 | 1 | | 1 | | | 1 | | 8.3 | 0.000 |
| | 2010 | 0.91(0.73-1.13) | | 0.95 | (0.88-1.02) | | 0.96(0.90-1.02) | | | |
| | 2015 | 0.8(0.62-1.05) | | 0.84 | (0.76-0.92) | *** | 0.86(0.80-0.92) | *** | | |
| Haiti | | | | | | | | | | |
| | 2005-06 | 1 | | 1 | | | 1 | | 4.55 | 0.000 |
| | 2012 | 0.66(0.49-0.89) | ** | 0.93 | (0.74-1.19) | | 1.25(0.93-1.68) | | | |
| | 2016-17 | 0.72(0.52-0.99) | * | 0.89 | (0.68-1.16) | | 1.22(0.89-1.66) | | | |
| India | | | | | | | | | | |
| | 2005-06 | 1 | | 1 | | | 1 | | 166.56 | 0.000 |
| | 2015-16 | 1.12(1.06-1.18) | *** | 1.1 | (1.01-1.20) | * | 1.18(1.10-1.26) | *** | | |
| | 2019-21 | 1.11(1.05-1.18) | *** | 1.05 | (0.96-1.16) | | 1.28(1.19-1.38) | *** | | |
| Jordan | | | | | | | | | | |
| | 2007 | 1 | | 1 | | | 1 | | 4.12 | 0.000 |
| | 2012 | 0.76(0.32-1.79) | | 0.79 | (0.49-1.28) | | 1.01(0.83-1.23) | | | |
| | 2017-18 | 1.02(0.51-2.04) | | 0.72 | (0.45-1.13) | | 0.99(0.81-1.22) | | | |
| | 2023 | 0.84(0.30-2.38) | | 0.58 | (0.32-1.05) | | 0.63(0.49-0.80) | *** | | |
| Kenya | | | | | | | | | | |
| | 2008-09 | 1 | | 1 | | | 1 | | 19.76 | 0.000 |
| | 2014 | 0.57(0.42-0.76) | *** | 0.81 | (0.71-0.92) | ** | 0.97(0.79-1.20) | | | |
| | 2022 | 0.49(0.38-0.63) | *** | 0.76 | (0.67-0.85) | *** | 0.79(0.67-0.94) | ** | | |
| Malawi | | | | | | | | | | |
| | 2004 | 1 | | 1 | | | 1 | | 8.82 | 0.000 |
| | 2010 | 1.03(0.82-1.29) | | 1.14 | (1.03-1.27) | * | 1.37(1.04-1.81) | * | | |
| | 2015-16 | 1.38(1.12-1.71) | ** | 1.29 | (1.17-1.43) | *** | 1.23(0.91-1.67) | | | |
| Mali | | | | | | | | | | |
| | 2006 | 1 | | 1 | | | 1 | | 3.6 | 0.000 |
| | 2012-13 | 1.36(1.18-1.57) | *** | 1.29 | (0.97-1.71) | | 1.7(1.09-2.65) | * | | |
| | 2018 | 1.05(0.90-1.21) | | 1.14 | (0.86-1.51) | | 1.43(0.94-2.17) | | | |
| Mozambique | | | | | | | | | | |
| | 2011 | 1 | | 1 | | | 1 | | 18.77 | 0.000 |
| | 2015 | 0.45(0.35-0.58) | *** | 0.51 | (0.43-0.61) | *** | 0.77(0.59-1.01) | | | |
| | 2022-23 | 0.66(0.53-0.82) | *** | 0.62 | (0.53-0.73) | *** | 0.56(0.43-0.72) | *** | | |
| Nepal | | | | | | | | | | |
| | 2011 | 1 | | 1 | | | 1 | | 14.28 | 0.000 |
| | 2016 | 0.79(0.64-0.98) | * | 1.09 | (0.77-1.54) | | 0.81(0.59-1.10) | | | |
| | 2022 | 1.05(0.85-1.30) | | 1.09 | (0.82-1.44) | | 0.94(0.69-1.29) | | | |
| Nigeria | | | | | | | | | | |
| | 2008 | 1 | | 1 | | | 1 | | 37.48 | 0.000 |
| | 2013 | 0.68(0.57-0.80) | *** | 0.78 | (0.67-0.90) | ** | 0.77(0.68-0.86) | *** | | |
| | 2018 | 1.26(1.06-1.50) | ** | 0.72 | (0.61-0.86) | *** | 0.8(0.70-0.91) | ** | | |
| Peru | | | | | | | | | | |
| | 2004-06 | 1 | | 1 | | | 1 | | 1.57 | 0.062 |

| | | | | | | | | | |
|---|---|---|---|---|---|---|---|---|---|
| 2007-08 | 0.98(0.66-1.46) | | 1.08 | (0.93-1.25) | | 1.1(0.97-1.24) | | | |
| 2009 | 1.3(0.89-1.89) | | 0.98 | (0.84-1.13) | | 1.04(0.93-1.16) | | | |
| 2010 | 0.68(0.43-1.07) | | 1.05 | (0.91-1.21) | | 1(0.90-1.13) | | | |
| 2011 | 0.8(0.54-1.20) | | 0.98 | (0.85-1.13) | | 1.01(0.90-1.13) | | | |
| 2012 | 1.13(0.76-1.70) | | 0.95 | (0.82-1.10) | | 0.93(0.82-1.04) | | | |
| Philippines | | | | | | | | | |
| 2008 | 1 | | 1 | | | 1 | | 14.85 | 0.000 |
| 2013 | 0.63(0.31-1.27) | | 0.73 | (0.58-0.90) | ** | 0.72(0.62-0.84) | *** | | |
| 2017 | 0.43(0.20-0.91) | * | 0.55 | (0.42-0.71) | *** | 0.57(0.48-0.68) | *** | | |
| 2022 | 0.26(0.09-0.78) | * | 0.51 | (0.36-0.73) | *** | 0.44(0.37-0.53) | *** | | |
| Rwanda | | | | | | | | | |
| 2010 | 1 | | 1 | | | 1 | | 6.98 | 0.000 |
| 2014-15 | 0.69(0.51-0.92) | * | 0.78 | (0.68-0.90) | *** | 0.58(0.35-0.95) | * | | |
| 2019-20 | 1.01(0.77-1.32) | | 0.89 | (0.78-1.03) | | 1.01(0.71-1.45) | | | |
| Senegal | | | | | | | | | |
| 2017 | 1 | | 1 | | | 1 | | 3.45 | 0.001 |
| 2018 | 0.48(0.31-0.74) | ** | 0.81 | (0.45-1.48) | | 1.01(0.45-2.29) | | | |
| 2019 | 0.44(0.30-0.64) | *** | 0.66 | (0.33-1.35) | | 0.56(0.25-1.26) | | | |
| Tanzania | | | | | | | | | |
| 2010 | 1 | | 1 | | | 1 | | 8.67 | 0.000 |
| 2015-16 | 0.92(0.78-1.09) | | 0.85 | (0.78-0.93) | *** | 0.78(0.58-1.04) | | | |
| 2022 | 0.81(0.66-0.98) | * | 0.78 | (0.70-0.87) | *** | 0.67(0.49-0.92) | * | | |
| Uganda | | | | | | | | | |
| 2006 | 1 | | 1 | | | 1 | | 17.09 | 0.000 |
| 2011 | 0.78(0.61-1.00) | | 0.78 | (0.68-0.90) | *** | 0.91(0.60-1.38) | | | |
| 2016 | 0.7(0.58-0.83) | *** | 0.72 | (0.65-0.80) | *** | 0.74(0.58-0.95) | * | | |
| Zambia | | | | | | | | | |
| 2007 | 1 | | 1 | | | 1 | | 30.65 | 0.000 |
| 2013-14 | 0.8(0.66-0.96) | * | 0.66 | (0.60-0.73) | *** | 0.56(0.50-0.64) | *** | | |
| 2018 | 0.84(0.69-1.03) | | 0.64 | (0.57-0.71) | *** | 0.51(0.45-0.59) | *** | | |
| Zimbabwe | | | | | | | | | |
| 2005-05 | 1 | | 1 | | | 1 | | 10.75 | 0.000 |
| 2010-11 | 0.67(0.42-1.07) | | 1.06 | (0.94-1.19) | | 1(0.90-1.11) | | | |
| 2015 | 0.61(0.32-1.14) | | 0.74 | (0.64-0.85) | *** | 0.79(0.70-0.88) | *** | | |

Notes: Estimates are adjusted for five-year age group, with Wald F-statistic for a test of interaction between survey and broad age group. CI = confidence interval, RR = risk ratio. * P-value<0.05, **p-value<0.01, ***p-value<0.001***

**Table S11: Stratified risk ratio of physical and/or sexual IPV by place of residence and country**

| | Survey year | Urban RR (95% CI) | | Rural RR (95% CI) | | | Wald test for interaction between residence and survey Wald F | P-value |
|---|---|---|---|---|---|---|---|---|
| Cambodia | | | | | | | | |
| | 2005 | 1 | | 1 | | | 13.34 | 0.000 |
| | 2014 | 0.88(0.52-1.48) | | 1.26 | (0.98-1.62) | | | |
| | 2021-22 | 0.61(0.39-0.97) | * | 0.69 | (0.54-0.89) | ** | | |
| Cameroon | | | | | | | | |
| | 2004 | 1 | | 1 | | | 16.14 | 0.000 |
| | 2011 | 0.91(0.80-1.04) | | 1.29 | (1.11-1.49) | ** | | |
| | 2018 | 0.6(0.51-0.69) | *** | 0.88 | (0.72-1.06) | | | |
| Colombia | | | | | | | | |
| | 2005 | 1 | | 1 | | | 15.7 | 0.000 |
| | 2010 | 0.96(0.91-1.01) | | 0.89 | (0.81-0.98) | * | | |
| | 2015 | 0.84(0.78-0.90) | *** | 0.82 | (0.73-0.91) | *** | | |
| Haiti | | | | | | | | |
| | 2005-06 | 1 | | 1 | | | 2.57 | 0.025 |
| | 2012 | 1.02(0.80-1.29) | | 0.81 | (0.64-1.02) | | | |
| | 2016-17 | 1.06(0.81-1.39) | | 0.75 | (0.59-0.95) | * | | |
| India | | | | | | | | |
| | 2005-06 | 1 | | 1 | | | 33.74 | 0.000 |
| | 2015-16 | 1.05(0.95-1.16) | | 1.01 | (0.96-1.06) | | | |
| | 2019-21 | 1.05(0.94-1.18) | | 0.98 | (0.93-1.03) | | | |
| Jordan | | | | | | | | |
| | 2007 | 1 | | 1 | | | 5.71 | 0.000 |
| | 2012 | 1.03(0.84-1.27) | | 0.71 | (0.53-0.94) | * | | |
| | 2017-18 | 0.99(0.80-1.23) | | 0.77 | (0.54-1.11) | | | |
| | 2023 | 0.64(0.50-0.83) | ** | 0.38 | (0.24-0.60) | *** | | |
| Kenya | | | | | | | | |
| | 2008-09 | 1 | | 1 | | | 15.59 | 0.000 |
| | 2014 | 0.84(0.69-1.03) | | 0.79 | (0.70-0.89) | *** | | |
| | 2022 | 0.63(0.52-0.77) | *** | 0.71 | (0.64-0.78) | *** | | |
| Malawi | | | | | | | | |
| | 2004 | 1 | | 1 | | | 7.09 | 0.000 |
| | 2010 | 1.52(1.14-2.03) | ** | 1.1 | (1.00-1.22) | | | |
| | 2015-16 | 1.41(1.01-1.98) | * | 1.26 | (1.15-1.39) | *** | | |
| Mali | | | | | | | | |
| | 2006 | 1 | | 1 | | | 6.74 | 0.000 |
| | 2012-13 | 1.27(1.00-1.60) | * | 1.36 | (1.17-1.59) | *** | | |
| | 2018 | 1.07(0.83-1.36) | | 1.07 | (0.91-1.25) | | | |
| Mozambique | | | | | | | | |
| | 2011 | 1 | | 1 | | | 30.8 | 0.000 |
| | 2015 | 0.66(0.53-0.81) | *** | 0.48 | (0.40-0.57) | *** | | |
| | 2022-23 | 0.53(0.44-0.63) | *** | 0.67 | (0.58-0.77) | *** | | |
| Nepal | | | | | | | | |
| | 2011 | 1 | | 1 | | | 2.56 | 0.026 |
| | 2016 | 0.65(0.50-0.85) | ** | 0.93 | (0.75-1.15) | | | |
| | 2022 | 0.86(0.67-1.10) | | 0.93 | (0.75-1.15) | | | |
| Nigeria | | | | | | | | |
| | 2008 | 1 | | 1 | | | 8.98 | 0.000 |
| | 2013 | 0.78(0.66-0.91) | ** | 0.71 | (0.63-0.81) | *** | | |
| | 2018 | 0.86(0.73-1.01) | | 0.97 | (0.85-1.10) | | | |
| Peru | | | | | | | | |
| | 2004-06 | 1 | | 1 | | | 2.25 | 0.010 |

| | | | | | | | |
|---|---|---|---|---|---|---|---|
| | 2007-08 | 1.09(0.95-1.24) | 1.07 | (0.92-1.24) | | | |
| | 2009 | 1.02(0.91-1.14) | 1.04 | (0.89-1.20) | | | |
| | 2010 | 0.99(0.88-1.12) | 1.02 | (0.89-1.18) | | | |
| | 2011 | 1.01(0.90-1.13) | 0.93 | (0.80-1.07) | | | |
| | 2012 | 0.93(0.82-1.05) | 0.94 | (0.81-1.09) | | | |
| Philippines | | | | | | | |
| | 2008 | 1 | 1 | | | 19.04 | 0.000 |
| | 2013 | 0.71(0.58-0.87) ** | 0.72 | (0.61-0.84) | *** | | |
| | 2017 | 0.52(0.41-0.66) *** | 0.58 | (0.48-0.69) | *** | | |
| | 2022 | 0.4(0.31-0.52) *** | 0.47 | (0.39-0.58) | *** | | |
| Rwanda | | | | | | | |
| | 2010 | 1 | 1 | | | 7.18 | 0.000 |
| | 2014-15 | 0.83(0.55-1.24) | 0.74 | (0.65-0.84) | *** | | |
| | 2019-20 | 0.81(0.56-1.15) | 0.9 | (0.79-1.02) | | | |
| Senegal | | | | | | | |
| | 2017 | 1 | 1 | | | 5.86 | 0.000 |
| | 2018 | 0.53(0.31-0.91) * | 0.72 | (0.50-1.04) | | | |
| | 2019 | 0.52(0.30-0.92) * | 0.48 | (0.34-0.67) | *** | | |
| Tanzania | | | | | | | |
| | 2010 | 1 | 1 | | | 9.88 | 0.000 |
| | 2015-16 | 0.84(0.70-1.01) | 0.85 | (0.77-0.93) | *** | | |
| | 2022 | 0.7(0.57-0.85) *** | 0.77 | (0.69-0.86) | *** | | |
| Uganda | | | | | | | |
| | 2006 | 1 | 1 | | | 23.47 | 0.000 |
| | 2011 | 0.85(0.57-1.27) | 0.77 | (0.67-0.88) | *** | | |
| | 2016 | 0.72(0.55-0.93) * | 0.7 | (0.63-0.77) | *** | | |
| Zambia | | | | | | | |
| | 2007 | 1 | 1 | | | 37.86 | 0.000 |
| | 2013-14 | 0.53(0.46-0.60) *** | 0.73 | (0.66-0.80) | *** | | |
| | 2018 | 0.51(0.44-0.60) *** | 0.67 | (0.61-0.75) | *** | | |
| Zimbabwe | | | | | | | |
| | 2005-05 | 1 | 1 | | | 11.1 | 0.000 |
| | 2010-11 | 1.08(0.92-1.26) | 0.96 | (0.87-1.06) | | | |

Notes: Estimates are adjusted for five-year age group, with Wald F-statistic for a test of interaction between survey and broad age group. CI = confidence interval, RR = risk ratio. * P-value<0.05, **p-value<0.01, ***p-value<0.001***

**Table S12: Stratified risk ratio of physical and/or sexual IPV by wealth group and country**

| | | Wealth | | | | | | | | | |
|---|---|---|---|---|---|---|---|---|---|---|---|
| Country | Survey year | Poor/poorest | | | Middle | | | Richer/richest | | | Wald test for interaction between wealth and survey time | |
| | | RR (95% CI) | | | RR (95% CI) | | | RR (95% CI) | | | Wald F | P-value |
| Cambodia | | | | | | | | | | | | |
| | 2005 | 1 | | | 1 | | | 1 | | | 13.68 | 0.000 |
| | 2014 | 1.47 | (1.12-1.93) | ** | 1.2 | (0.72-2.02) | | 0.89 | (0.55-1.44) | | | |
| | 2021-22 | 0.82 | (0.62-1.07) | | 0.55 | (0.34-0.89) | * | 0.41 | (0.26-0.65) | *** | | |
| Cameroon | | | | | | | | | | | | |
| | 2004 | 1 | | | 1 | | | 1 | | | 11.07 | 0.000 |
| | 2011 | 1.28 | (1.08-1.52) | ** | 1.17 | (0.95-1.43) | | 0.9 | (0.78-1.03) | | | |
| | 2018 | 0.94 | (0.75-1.17) | | 0.77 | (0.62-0.96) | * | 0.54 | (0.46-0.63) | *** | | |
| Colombia | | | | | | | | | | | | |
| | 2005 | 1 | | | 1 | | | 1 | | | 15.31 | 0.000 |
| | 2010 | 0.88 | (0.83-0.95) | *** | 1 | (0.92-1.09) | | 0.97 | (0.89-1.06) | | | |
| | 2015 | 0.86 | (0.80-0.93) | *** | 0.86 | (0.78-0.95) | ** | 0.76 | (0.67-0.86) | *** | | |
| Haiti | | | | | | | | | | | | |
| | 2005-06 | 1 | | | 1 | | | 1 | | | 1.48 | 0.158 |
| | 2012 | 0.86 | (0.67-1.11) | | 0.88 | (0.65-1.21) | | 0.92 | (0.72-1.17) | | | |
| | 2016-17 | 0.85 | (0.64-1.13) | | 0.76 | (0.55-1.06) | | 0.96 | (0.73-1.25) | | | |
| India | | | | | | | | | | | | |
| | 2005-06 | 1 | | | 1 | | | 1 | | | 154.66 | 0.000 |
| | 2015-16 | 1 | (0.95-1.05) | | 0.97 | (0.91-1.04) | | 1.1 | (1.01-1.19) | * | | |
| | 2019-21 | 0.94 | (0.89-1.00) | * | 0.94 | (0.86-1.01) | | 1.1 | (1.01-1.21) | * | | |
| Jordan | | | | | | | | | | | | |
| | 2007 | 1 | | | 1 | | | 1 | | | 6.22 | 0.000 |
| | 2012 | 1.04 | (0.83-1.30) | | 1.07 | (0.75-1.51) | | 0.82 | (0.58-1.15) | | | |
| | 2017-18 | 0.91 | (0.72-1.14) | | 0.87 | (0.61-1.25) | | 1.1 | (0.79-1.55) | | | |
| | 2023 | 0.7 | (0.52-0.94) | * | 0.7 | (0.47-1.05) | | 0.48 | (0.32-0.74) | ** | | |
| Kenya | | | | | | | | | | | | |
| | 2008-09 | 1 | | | 1 | | | 1 | | | 14.42 | 0.000 |
| | 2014 | 0.82 | (0.70-0.96) | * | 0.82 | (0.67-1.00) | * | 0.75 | (0.63-0.91) | ** | | |
| | 2022 | 0.75 | (0.66-0.87) | *** | 0.71 | (0.61-0.84) | *** | 0.57 | (0.48-0.66) | *** | | |

| | | | | | | | | | | | |
|---|---|---|---|---|---|---|---|---|---|---|---|
| Malawi | | | | | | | | | | | |
| | 2004 | 1 | | | 1 | | | 1 | | | 6.14 | 0.000 |
| | 2010 | 1.07 | (0.94-1.23) | | 1.24 | (1.04-1.49) | * | 1.2 | (1.02-1.41) | * | | |
| | 2015-16 | 1.29 | (1.14-1.46) | *** | 1.37 | (1.14-1.65) | ** | 1.2 | (1.01-1.43) | * | | |
| Mali | | | | | | | | | | | | |
| | 2006 | 1 | | | 1 | | | 1 | | | 3.57 | 0.000 |
| | 2012-13 | 1.36 | (1.12-1.64) | ** | 1.46 | (1.13-1.88) | ** | 1.33 | (1.09-1.61) | ** | | |
| | 2018 | 0.96 | (0.79-1.18) | | 1.21 | (0.93-1.57) | | 1.13 | (0.94-1.36) | | | |
| Mozambique | | | | | | | | | | | | |
| | 2011 | 1 | | | 1 | | | 1 | | | 18.06 | 0.000 |
| | 2015 | 0.46 | (0.36-0.58) | *** | 0.39 | (0.27-0.55) | *** | 0.7 | (0.59-0.84) | *** | | |
| | 2022-23 | 0.68 | (0.56-0.83) | *** | 0.66 | (0.52-0.84) | ** | 0.53 | (0.45-0.63) | *** | | |
| Nepal | | | | | | | | | | | | |
| | 2011 | 1 | | | 1 | | | 1 | | | 4.99 | 0.000 |
| | 2016 | 0.81 | (0.63-1.05) | | 0.72 | (0.51-1.01) | | 0.81 | (0.61-1.08) | | | |
| | 2022 | 1.02 | (0.81-1.28) | | 0.73 | (0.54-0.99) | * | 0.95 | (0.71-1.26) | | | |
| Nigeria | | | | | | | | | | | | |
| | 2008 | 1 | | | 1 | | | 1 | | | 13.59 | 0.000 |
| | 2013 | 0.76 | (0.64-0.89) | ** | 0.8 | (0.68-0.95) | ** | 0.67 | (0.58-0.77) | *** | | |
| | 2018 | 1.14 | (0.97-1.34) | | 0.88 | (0.75-1.04) | | 0.74 | (0.62-0.88) | ** | | |
| Peru | | | | | | | | | | | | |
| | 2004-06 | 1 | | | 1 | | | 1 | | | 5.48 | 0.000 |
| | 2007-08 | 1.07 | (0.92-1.24) | | 1.07 | (0.91-1.26) | | 1.13 | (0.94-1.35) | | | |
| | 2009 | 1.08 | (0.95-1.23) | | 0.94 | (0.81-1.10) | | 1.01 | (0.85-1.21) | | | |
| | 2010 | 1.04 | (0.92-1.18) | | 1 | (0.85-1.17) | | 0.96 | (0.81-1.14) | | | |
| | 2011 | 1.02 | (0.89-1.16) | | 0.99 | (0.84-1.17) | | 0.94 | (0.79-1.11) | | | |
| | 2012 | 0.96 | (0.85-1.10) | | 0.93 | (0.78-1.10) | | 0.9 | (0.75-1.07) | | | |
| Philippines | | | | | | | | | | | | |
| | 2008 | 1 | | | 1 | | | 1 | | | 18.8 | 0.000 |
| | 2013 | 0.69 | (0.59-0.80) | *** | 0.74 | (0.58-0.94) | * | 0.77 | (0.59-1.01) | | | |
| | 2017 | 0.6 | (0.51-0.71) | *** | 0.49 | (0.36-0.65) | *** | 0.52 | (0.37-0.72) | *** | | |
| | 2022 | 0.45 | (0.37-0.54) | *** | 0.45 | (0.33-0.63) | *** | 0.38 | (0.26-0.55) | *** | | |
| Rwanda | | | | | | | | | | | | |
| | 2010 | 1 | | | 1 | | | 1 | | | 9.23 | 0.000 |
| | 2014-15 | 0.73 | (0.62-0.87) | *** | 0.63 | (0.48-0.83) | ** | 0.84 | (0.67-1.06) | | | |
| | 2019-20 | 0.9 | (0.78-1.05) | | 0.88 | (0.68-1.12) | | 0.87 | (0.69-1.09) | | | |

| | | | | | | | | | | | |
|---|---|---|---|---|---|---|---|---|---|---|---|
| Senegal | | | | | | | | | | | |
| | 2017 | 1 | | | 1 | | | 1 | | | 3.69 | 0.000 |
| | 2018 | 0.71 | (0.50-1.01) | | 0.61 | (0.31-1.23) | | 0.58 | (0.29-1.13) | | | |
| | 2019 | 0.48 | (0.34-0.69) | *** | 0.33 | (0.13-0.79) | * | 0.63 | (0.33-1.21) | | | |
| Tanzania | | | | | | | | | | | | |
| | 2010 | 1 | | | 1 | | | 1 | | | 8.86 | 0.000 |
| | 2015-16 | 0.94 | (0.84-1.05) | | 0.86 | (0.75-0.99) | * | 0.74 | (0.64-0.85) | *** | | |
| | 2022 | 0.89 | (0.78-1.02) | | 0.68 | (0.57-0.80) | *** | 0.66 | (0.56-0.76) | *** | | |
| Uganda | | | | | | | | | | | | |
| | 2006 | 1 | | | 1 | | | 1 | | | 22.71 | 0.000 |
| | 2011 | 0.83 | (0.71-0.96) | * | 0.81 | (0.61-1.08) | | 0.69 | (0.55-0.87) | ** | | |
| | 2016 | 0.7 | (0.63-0.78) | *** | 0.74 | (0.60-0.90) | ** | 0.61 | (0.52-0.72) | *** | | |
| Zambia | | | | | | | | | | | | |
| | 2007 | 1 | | | 1 | | | 1 | | | 26.19 | 0.000 |
| | 2013-14 | 0.76 | (0.68-0.84) | *** | 0.76 | (0.67-0.86) | *** | 0.5 | (0.44-0.57) | *** | | |
| | 2018 | 0.76 | (0.68-0.86) | *** | 0.63 | (0.54-0.75) | *** | 0.47 | (0.41-0.55) | *** | | |
| Zimbabwe | | | | | | | | | | | | |
| | 2005-05 | 1 | | | 1 | | | 1 | | | 8.73 | 0.000 |
| | 2010-11 | 0.94 | (0.83-1.06) | | 0.94 | (0.79-1.13) | | 1.07 | (0.94-1.23) | | | |

Notes: Estimates are adjusted for five-year age group, with Wald F-statistic for a test of interaction between survey and broad age group. CI = confidence interval, RR = risk ratio. * P-value<0.05, **p-value<0.01, ***p-value<0.001***

**Sensitivity Analysis**

Ever having experienced recent physical and/or IPV was defined as saying yes to having experienced any of the items on the standardised list in the last 12 months. In some instances, an individual reports experiencing IPV but the recall period, whether it was in the last 12 months or not, was not known. In the main analysis when the recall period was unknown we recode the last 12 months as no experience of IPV. We also excluded women from the analysis if all the responses to each item were missing. To assess whether this assumption could change the interpretation of the results, we calculated the proportion of women who were coded as not having experienced recent IPV who had at least one item where the recall period of IPV was unknown and the proportion of women who had all information on experience of IPV missing. Only a small proportion of the final domestic violence sample we used had missing data for all items between 0.00 and 0.58% (table S13). For women who were coded as having not experienced recent IPV but had at least one item where they reporting having experienced the specific IPV but the recall period was not recorded was under 1% in all surveys apart from Cameroon 2004 (2.19%), Jordan 2007 (1.49%), Zambia 2007 (1.63%) and Zimbabwe 2005-06 (2.79%). If we assumed the extreme that all these women had actually experienced recent IPV none of the trends seen would change, apart from Zimbabwe where there would have been evidence of a decline in the first period as well as the second period.

**Table S13: Prevalence of non-missing and missing observations for IPV outcomes.** Columns from left to right are: Percentage of individuals who 1) had complete information and reported no 12-month physical and/or sexual IPV, 2) had at least one item of 12-month physical and/or sexual IPV, 3) for some items information was missing, but for those not missing reported no 12-month physical and/or sexual IPV was reported who reported no recent IPV , therefore could potentially be miss-coded as not having experience 12-month physical and/or sexual IPV 4) No information on any items, so excluded from the analysis.

| Survey | No recent IPV (%) | Recent IPV (%) | No recent IPV at least one unknown (%) | All information missing (%) | Total (%) |
|---|---|---|---|---|---|
| Cambodia 2005 | 90.94 | 9.05 | 0.02 | 0.00 | 100 |
| Cambodia 2014 | 89.03 | 10.95 | 0.01 | 0.02 | 100 |
| Cambodia 2021- 22 | 94.43 | 5.57 | 0.00 | 0.00 | 100 |
| Cameroon 2004 | 67.33 | 30.46 | 2.19 | 0.02 | 100 |
| Cameroon 2011 | 66.58 | 32.58 | 0.72 | 0.11 | 100 |
| Cameroon 2018 | 78.08 | 21.92 | 0.00 | 0.00 | 100 |
| Colombia 2005 | 79.09 | 20.91 | 0.00 | 0.00 | 100 |
| Colombia 2010 | 80.31 | 19.69 | 0.00 | 0.00 | 100 |
| Colombia 2015 | 82.52 | 17.48 | 0.00 | 0.00 | 100 |
| Dominican Republic 2002 | 80.78 | 19.12 | 0.00 | 0.10 | 100 |
| Dominican Republic 2007 | 87.26 | 11.48 | 1.26 | 0.00 | 100 |
| Dominican Republic 2013 | 84.20 | 15.63 | 0.15 | 0.02 | 100 |
| Haiti 2005-06 | 82.36 | 17.54 | 0.10 | 0.00 | 100 |
| Haiti 2012 | 84.54 | 15.16 | 0.29 | 0.00 | 100 |
| Haiti 2016-17 | 85.84 | 14.16 | 0.00 | 0.00 | 100 |
| India 2005-06 | 75.85 | 23.95 | 0.20 | 0.00 | 100 |
| India 2015-16 | 75.92 | 24.08 | 0.00 | 0.00 | 100 |
| India 2019-21 | 76.15 | 23.85 | 0.00 | 0.00 | 100 |
| Jordan 2007 | 83.90 | 14.62 | 1.49 | 0.00 | 100 |
| Jordan 2012 | 85.91 | 14.09 | 0.00 | 0.00 | 100 |
| Jordan 2017-18 | 86.18 | 13.82 | 0.00 | 0.00 | 100 |
| Jordan 2023 | 91.72 | 8.28 | 0.00 | 0.00 | 100 |
| Kenya 2008-09 | 67.84 | 31.69 | 0.48 | 0.00 | 100 |
| Kenya 2014 | 74.10 | 25.12 | 0.67 | 0.10 | 100 |
| Kenya 2022 | 79.04 | 20.96 | 0.00 | 0.00 | 100 |
| Malawi 2004 | 80.45 | 19.51 | 0.00 | 0.04 | 100 |
| Malawi 2010 | 77.20 | 22.34 | 0.46 | 0.00 | 100 |
| Malawi 2015-16 | 75.38 | 24.62 | 0.00 | 0.00 | 100 |
| Mali 2006 | 80.15 | 19.51 | 0.21 | 0.13 | 100 |
| Mali 2012-13 | 73.37 | 26.63 | 0.00 | 0.00 | 100 |
| Mali 2018 | 78.91 | 21.09 | 0.00 | 0.00 | 100 |
| Mozambique 2011 | 72.35 | 27.65 | 0.00 | 0.00 | 100 |
| Mozambique 2015 | 84.56 | 14.84 | 0.37 | 0.22 | 100 |
| Mozambique 2022-23 | 83.00 | 17.00 | 0.00 | 0.00 | 100 |
| Nepal 2011 | 85.72 | 14.28 | 0.00 | 0.00 | 100 |
| Nepal 2016 | 88.76 | 11.24 | 0.00 | 0.00 | 100 |
| Nepal 2022 | 86.98 | 13.02 | 0.00 | 0.00 | 100 |
| Nigeria 2008 | 84.06 | 15.16 | 0.34 | 0.43 | 100 |

| | | | | | |
|---|---|---|---|---|---|
| Nigeria 2013 | 88.39 | 11.08 | 0.54 | 0.00 | 100 |
| Nigeria 2018 | 86.05 | 13.95 | 0.00 | 0.00 | 100 |
| Peru 2004-06 | 86.16 | 13.74 | 0.10 | 0.00 | 100 |
| Peru 2007-08 | 84.99 | 14.93 | 0.07 | 0.00 | 100 |
| Peru 2009 | 85.83 | 14.17 | 0.00 | 0.00 | 100 |
| Peru 2010 | 86.12 | 13.88 | 0.00 | 0.00 | 100 |
| Peru 2011 | 86.40 | 13.60 | 0.00 | 0.00 | 100 |
| Peru 2012 | 87.12 | 12.88 | 0.00 | 0.00 | 100 |
| Philippines 2008 | 89.92 | 10.03 | 0.05 | 0.00 | 100 |
| Philippines 2013 | 92.78 | 7.11 | 0.11 | 0.00 | 100 |
| Philippines 2017 | 94.58 | 5.42 | 0.00 | 0.00 | 100 |
| Philippines 2022 | 95.89 | 4.11 | 0.00 | 0.00 | 100 |
| Rwanda 2010 | 72.88 | 26.89 | 0.23 | 0.00 | 100 |
| Rwanda 2014-15 | 79.38 | 20.10 | 0.47 | 0.06 | 100 |
| Rwanda 2019-20 | 76.60 | 23.40 | 0.00 | 0.00 | 100 |
| Senegal 2017 | 87.84 | 12.16 | 0.00 | 0.00 | 100 |
| Senegal 2018 | 92.42 | 7.58 | 0.00 | 0.00 | 100 |
| Senegal 2019 | 93.91 | 6.09 | 0.00 | 0.00 | 100 |
| Tanzania 2010 | 64.22 | 35.27 | 0.51 | 0.00 | 100 |
| Tanzania 2015-16 | 70.64 | 29.36 | 0.00 | 0.00 | 100 |
| Tanzania 2022 | 73.94 | 26.06 | 0.00 | 0.00 | 100 |
| Uganda 2006 | 54.93 | 44.81 | 0.26 | 0.00 | 100 |
| Uganda 2011 | 64.51 | 34.58 | 0.91 | 0.00 | 100 |
| Uganda 2016 | 69.67 | 30.33 | 0.00 | 0.00 | 100 |
| Zambia 2007 | 56.35 | 42.01 | 1.63 | 0.00 | 100 |
| Zambia 2013-14 | 73.00 | 26.51 | 0.43 | 0.06 | 100 |
| Zambia 2018 | 75.03 | 24.97 | 0.00 | 0.00 | 100 |
| Zimbabwe 2005-06 | 69.31 | 27.32 | 2.79 | 0.58 | 100 |
| Zimbabwe 2010-11 | 72.43 | 27.15 | 0.41 | 0.00 | 100 |
| Zimbabwe 2015 | 80.15 | 19.85 | 0.00 | 0.00 | 100 |

**Figure S8: Crude prevalence of recent physical and/or sexual IPV with robustness checks for definition of partner.** Comparing using only the most recent partner (circles) to using any partner in the past 12 months (triangles).

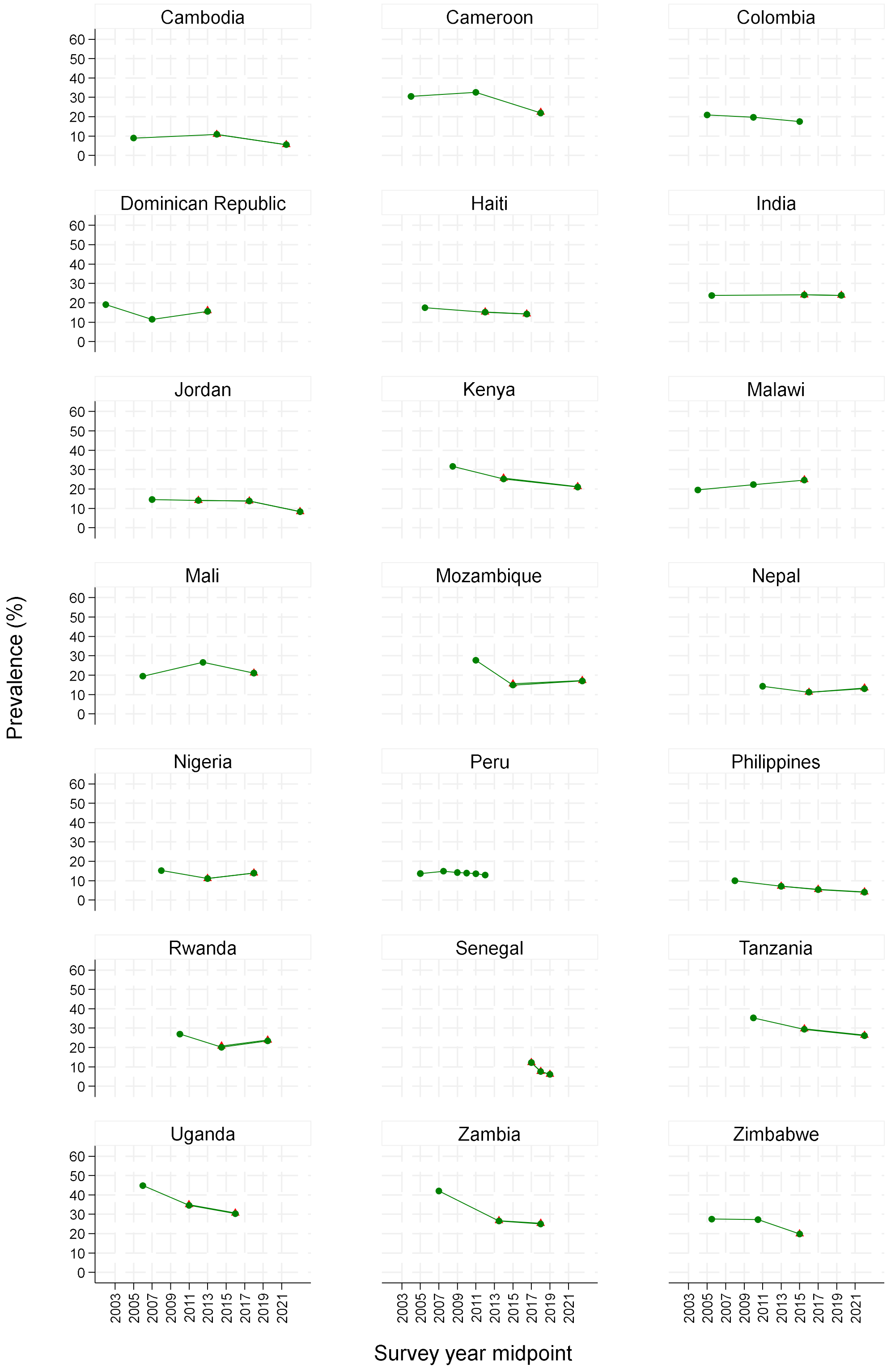

**Figure S9: Trends over time in crude prevalence for recent physical and/or sexual IPV with robustness checks for item wording changes.** Figures assess whether wording changes over time in questions 105b ("ever been slapped by husband/partner") and 105j ("ever had arm twisted or hair pulled by husband/partner"), adding reference to "pulling hair," might change trends in recent IPV. The countries shown (Cambodia, Cameroon, Dominican Republic and Malawi) were countries where wording changed over the surveys. Recent (12-month) physical and/or sexual IPV using all items is shown (blue) compared to all items, excluding question items d105b and d105j (red).

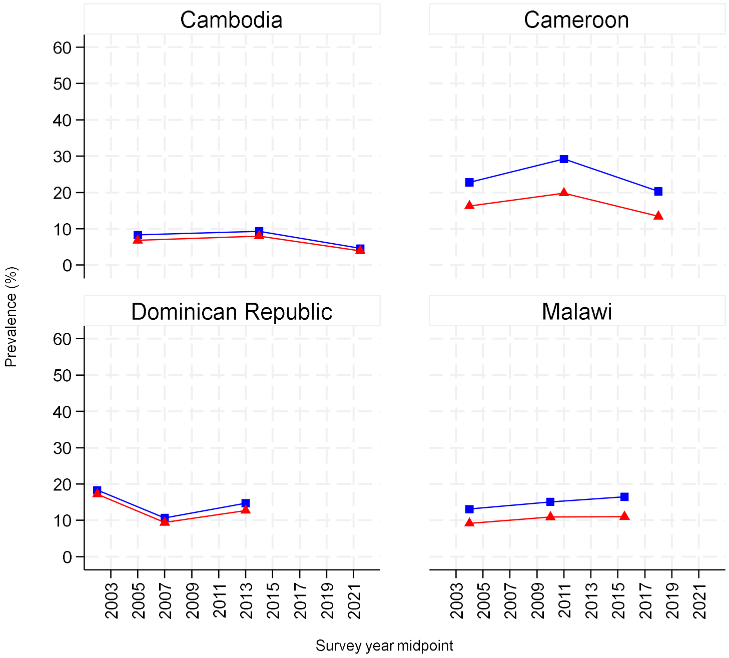
